# Early-life exposure to persistent organic pollutants, gut microbiota diversity and metabolites, and respiratory health in Norwegian children

**DOI:** 10.1101/2022.08.30.22279379

**Authors:** Virissa Lenters, Lützen Portengen, Merete Eggesbø, Roel Vermeulen

## Abstract

**Background:** Evidence suggests that early-life exposure to certain environmental chemicals increases the risk of allergic diseases, while gut microbiota diversity and microbiota-derived short-chain fatty acid (SCFA) metabolites may be protective.

**Objectives:** We assessed associations between persistent organic pollutants (POPs), microbial markers, and subsequent risk of asthma and lower respiratory tract infection (LRTI).

**Methods:** We studied a Norwegian birth cohort (HUMIS). Twenty-six POPs [polychlorinated biphenyls (PCBs), polybrominated diphenyl ethers (PBDEs), perfluoroalkyl substances (PFASs), and organochlorine pesticides (OCPs)] were quantified in maternal breastmilk (n=993). Shannon diversity and SCFAs were assessed at multiple time points before 2 years of age in a subset of children. We evaluated registry-based diagnosis of asthma when children were a median age of 10 years, along with maternal-reported asthma and LRTI by 2 years of age.

**Results:** ∑_14_PCBs was associated with decreased odds and ∑_4_OCPs with increased odds of asthma; associations between β-HCH (OR=2.99 per 2-SD increase; 95% CI: 1.66, 5.43) and PCB-138 (OR=0.43; 95% CI: 0.20, 0.91) and asthma by age 10 years were most robust. PBDEs and PFASs were not consistently associated with asthma and no POPs were associated with LRTI. There were both inverse and positive associations between diversity and respiratory outcomes, and generally imprecise associations for SCFAs. There was limited evidence that POP exposures perturbed diversity or production of SCFAs, except for an association between ∑_14_PCBs and reduced diversity at 2 years, and there was no clear evidence of mediation effects.

**Conclusions:** This study provides support for associations between some POPs and risk of childhood asthma, and indications of a potential independent role of gut microbiota.

---

Asthma is one of the most common chronic conditions in childhood, and together with lower respiratory tract infections (LRTIs), is a leading cause of hospitalization in young children (Asher and Pearce 2014; Nair et al. 2013). Worldwide variability and temporal trends in the prevalence of asthma—namely decades of increasing prevalence which appear to have peaked at the turn of the century in higher-income countries—provide strong support for environmental causal risk factors (Eder et al. 2006; Lai et al. 2009).

Epidemiological evidence suggests that prenatal and early-life exposure to ubiquitous environmental chemicals may disrupt the developing immune system and increase the risk of allergic diseases (Gascon et al. 2013). Several studies have reported both positive and inverse associations between exposure to persistent organic pollutants (POPs) and asthma, wheeze, and related symptoms; including associations with polychlorinated biphenyls (PCBs), organochlorines pesticides (OCPs), brominated flame retardants such as polybrominated diphenyl ethers (PBDEs), and fluoro-surfactants such as poly- and perfluoroalkyl substances (PFASs) (Gascon et al. 2012; Hansen et al. 2014; Humblet et al. 2014; Impinen et al. 2019; Smit et al. 2015). While many of these compounds have been restricted or prohibited, human exposure continues due to their persistence in the environment and the long life cycle of some chemically treated household products.

Gut microbiota play a crucial role in maturation of the immune system (Mazmanian et al. 2005; Olszak et al. 2012), but colonization of the newborn appears altered in westernized countries (Vangay et al. 2018; Yatsunenko et al. 2012). Perturbations of microbiota composition (dysbiosis), and bacterial ‘imprinting’ during sensitive developmental windows, may have long-lasting consequences for health (Gilbert et al. 2018). Short-chain fatty acids (SCFAs), primarily produced by gut microbiota during fermentation of non-digestible fibers, are also immunomodulatory; for example, SCFAs induce regulatory T cells and activate G-protein-coupled receptors (GPR41 and GPR43) (Arpaia et al. 2013; Smith et al. 2013). In mice, a fiber-rich diet and the concomitant increase in circulating SCFAs were protective against allergic airway inflammation (Trompette et al. 2014). Emerging evidence from prospective epidemiological studies support that greater microbial diversity and higher levels of anti-inflammatory SCFAs protect against allergic disease (Abrahamsson et al. 2014; Arrieta et al. 2015; Bisgaard et al. 2011; Patrick et al. 2020; Penders et al. 2007; Ronduit et al. 2019).

Environmental chemicals, including some heavy metals and POPs, have been found to perturb gut microbiota composition and metabolic activity in small number of animal experimental studies and human observational studies (Chiu et al. 2020; Iszatt et al. 2019). We evaluated the risk of asthma and LRTIs in association with early-life chemical profiles and microbiota diversity and metabolic products. Furthermore, we explored potential mediation of chemical–outcome associations by these microbiota markers.

## Methods

### Study population

We used data from the prospective Norwegian HUMIS birth cohort (n=2606), which has been described in detail elsewhere (Dahl et al. 2018; Eggesbo et al. 2011). Briefly, mothers were recruited in 2003–2009 by public health nurses during routine postnatal care home visits in seven counties across Norway. A subset of mothers were recruited by a pediatrician at the maternity ward in Østfold county hospital in Southern Norway in 2002–2005, where for every mother of a preterm infant (<37 weeks gestation) enrolled, two mothers of term infants were consecutively enrolled. These mothers were asked to collect a fecal sample 4 days postpartum and from their infants when they were 4, 10, and 30 days, and 4, 12, and 24 months old. All mothers were asked to collect breast milk on eight consecutive days before the child reached 2 months of age. Minor deviations in this sampling protocol, such as collection by breast pump, were accepted (see Supplemental Methods). The study was approved by the Regional Committees for Medical and Health Research Ethics and the Norwegian Data Inspectorate, and written informed consent was obtained from all participating women prior to enrolment.

We restricted analyses to mother–child pairs with singletons. For analyses of POP exposures and respiratory health outcomes, the sample size with available POP data was 993, of which 157 were oversampled for small for gestational age (SGA) [<10^th^ percentile of sex-specific Norwegian standards (Skjaerven et al. 2000)], rapid growth in the first 6 months [change in weight *z*-score >0.67 (Monteiro and Victora 2005)], or (at enrolment) for preterm birth. For analyses of microbiota markers and outcomes, the sample size available was 438, and for analyses of POPs and microbiota markers it was 298 (see Figure S1 for a flow chart of the study population samples).

### Respiratory health outcomes

The primary outcome of interest was childhood asthma. We linked to the Norwegian Patient Registry to identify children who had ever been diagnosed with asthma by a health specialist [based on the International Classification of Diseases, 10th revision (ICD-10) codes J45 and J46]. The registry collects diagnostic data from all hospitals and outpatient clinics in Norway. Individual records are available from 2008 onwards, when children (n=993 sample) were a median of 3.7 years old [interquartile range (IQR), 2.9–4.5; range, 0–5.1], and were available up until August 2014 at the time of linkage, when children were 10.4 years old (IQR, 9.6–11.2; range, 4.8–11.8). We also evaluated two secondary outcomes that were ascertained based on questionnaires administered to mothers at 6, 12, and 24 months postpartum: current asthma at 2 years of age, and the occurrence of one or more

LRTIs by 2 years, defined as maternal-reported pneumonia, bronchitis, or respiratory syncytial virus infections. Early-life LRTIs predict the development of asthma, although it is unclear whether this reflects causal or common causes (Rantala et al. 2015). Asthma at 2 years of age may represent early-onset transient or persistent wheeze phenotypes (Savenije et al. 2011), clinically diagnosed or interpreted by the mother as asthma, which may resolve later in childhood.

### Chemical exposure assessment

Chemicals were measured using gas or liquid chromatography coupled to mass spectrometry, as previously described (Forns et al. 2015; Polder et al. 2009; Thomsen et al. 2010) (see also Supplemental Methods). We evaluated the 26 chemicals which were measured and detected in ≥70% of samples: seven dioxin-like (DL) PCBs (mono-*ortho* congeners 105, 114, 118, 156, 157, 167, and 189) and seven non-dioxin-like (NDL) PCBs (congeners 74, 99, 138, 153, 170, 180, and 194); four organochlorine pesticides (OCPs), hexachlorobenzene (HCB), β-hexachlorocyclohexane (β-HCH), oxychlordane, and dichlorodiphenyldichloroethylene (DDE); six PBDEs, congeners 28, 47, 99, 100, 153, and 154; and two PFASs, perfluorooctanoate (PFOA) and perfluorooctane sulfonate (PFOS).

Values below the limit of detection (LOD) were imputed (0–2.5% for 24 chemicals, 10% for PFOA, and 18% BDE-154). We used single conditional imputation, including the other exposure variables, maternal age, and parity as predictors, using maximum likelihood estimation assuming a log-normal distribution (Lubin et al. 2004). Molar concentrations of the DL-PCBs, NDL-PCBs, OCPs, PBDEs, and PFASs were summed. Breast milk was collected at a median of 32 days postpartum (IQR, 24–43). There was no significant trend in concentration levels with timing of sampling for nearly all chemicals (the exception was a minor decreasing trend for PFOA), so we did not calibrate by sampling period.

### SCFAs and gut microbiota diversity

Eight SCFAs were measured using gas chromatography with flame ionization detection following previously published analytical protocols (Hoverstad et al. 1984; Midtvedt et al. 1988; Zijlstra et al. 1977) (see Supplemental Methods). We did not assess *n*-caproic or *i*-caproic in the present analysis as the majority (>74%) of values were below the detection limit. We imputed SCFA data below the LOD using the method described above (Lubin et al. 2004). Microbiota composition was characterized using DNA extraction and purification, PCR amplification of the V4 region of the 16S rRNA gene, and sequencing using the Illumina HiSeq instrument, as elaborated in the Supplemental Methods. We calculated the Shannon diversity index, a robust measure of taxonomic within individual (α-)diversity that accounts for both richness and evenness of species (Haegeman et al. 2013).

### Covariates

Information on potential confounders was obtained from the questionnaires completed by the mothers 6, 12, and 24 months postpartum, and supplemented with information from the Medical Birth Registry of Norway (child’s sex, gestational age, birth weight, and maternal pre-pregnancy height and weight, and smoking during pregnancy).

### Statistical analysis

#### Associations between POPs and respiratory outcomes

First, we estimated associations between POPs and the three respiratory outcomes. To account for the complex correlation pattern of the POP exposures and to mitigate potential estimation problems due to collinearity (Lenters et al. 2018), we modelled all POPs simultaneously using a multi-pollutant penalized regression approach for variable selection, elastic net regression (Zou and Hastie 2005). A generalized linear model (GLM) is fitted with an added penalty that maximizes the penalized log-likelihood in logistic regression, shrinking coefficients towards or exactly to zero, thereby achieving selection. The elastic net penalty is a weighted sum of the ridge penalty, proportional to the sum of the squared regression coefficients, and the lasso penalty, proportional to the absolute value of the coefficients, which allows for selection of groups of collinear variables.

We refit the elastic net-selected subset in a multi-pollutant logistic regression (with ordinary maximum likelihood estimation) model to obtain effect estimates which were not biased due to penalization. We also assessed associations with summed molar concentrations of the PCBs, OCPs, PBDEs, and PFASs, under the assumptions that each molecule has a single potent binding site, and that chemicals within a group act via common biological pathways. Given the low correlations across summed chemical groups, we assessed unpenalized multivariable models. For comparison, we also present the single-pollutant effect estimates for individual and summed chemicals from logistic regression models, and applied false discovery rate correction (Benjamini and Hochberg 1995).

Based on *a priori* knowledge, we considered the minimal sufficient adjustment set for the models of POP exposures and asthma and LRTI outcomes to include maternal age (years), parity (nulliparous, multiparous), pre-pregnancy body mass index (continuous), maternal education (≤12, >12 years education, as a proxy of socioeconomic position), diet, and maternal stress (see directed acyclic graph, Figure S2A). We also tested models further adjusted for covariates for which *a priori* evidence of potential confounding is weaker: child birth year (continuous), child sex, delivery mode (C-section vs. vaginal), duration of any breastfeeding (months), parental asthma (maternal or paternal), maternal fish consumption in the year before delivery (≤23, >23 servings), and the number of playmates at age 2 years, as a proxy of daycare attendance (≤4, >4 playmates). We did not incorporate sampling weights (16% were oversampled for preterm, SGA, or rapid growth) in any of the models because this may reduce precision when covariates related to sampling are included in models of exposure–outcome associations (Gelman 2007), and only adjusted for sampling design covariates deemed to be confounders (Wirth and Tchetgen Tchetgen 2014).

We used multiple imputation by chained equations (Buuren and Groothuis-Oudshoorn 2011) to impute missing outcome data (0% for registry-based asthma by 2014, 13% for maternal-reported asthma at 2 years, and 21% for LRTI), chemical exposure data (because the sample was not analyzed for that chemical; 0–4% for 23 chemicals, and 11% for 3 chemicals) and covariate data (0–3% for covariates in the minimally adjusted models, and 0–23% in the further adjusted models). We imputed 100 datasets using predictive mean matching for continuous variables and logistic regression for binary variables, and estimates were combined using Rubin’s rules (see Supplemental Methods for additional details). As a sensitivity analysis, we also ran complete case analyses.

Chemical exposures were natural log (ln)–transformed in all analyses to reduce the influence of extreme values. We input exposures and covariates which were scaled to two standard deviations (2-SD) to impart variables with same prior probability of selection independent of the contrast in levels, and to facilitate comparison of coefficients across variables (Gelman 2008). In elastic net models, we set covariates to be unpenalized. We repeated elastic net modeling using 100 multiply imputed datasets, and averaged regression coefficients for the exposures that were selected in at least half of the models (Lenters et al. 2019; Wood et al. 2008). Selection (i.e., the optimal level of penalization) within imputed datasets was determined using 10-fold cross-validation. We also assessed potential effect modification by child sex and maternal education and smoking.

#### Associations between gut microbiota markers and respiratory outcomes

Second, we assessed associations between Shannon diversity and total SCFA levels and the respiratory outcomes using univariable logistic regression models, and for the six individual SCFAs using penalized multivariable logistic regression models. We assessed the samples from 4, 12, and 24 months; this was because the most samples had been analyzed for these sampling time points, increasing statistical power, and because this represents a time period when colonization of the gut is well underway but highly plastic (Palmer et al. 2007; Yatsunenko et al. 2012) and when the immune system is still rapidly maturing (Simon et al. 2015). We scaled SCFAs to 2-SD prior to modeling. As a sensitivity analysis, we repeated the modeling using the relative abundance of each SCFA, normalized to the total SCFAs, and using isometric log-ratio transformation of SCFAs to account for the compositional data, or dependence between SCFA proportions (Egozcue et al. 2003; Filzmoser et al. 2010). These models were adjusted for maternal education, preterm birth, and maternal smoking during pregnancy and when the child was one year old. As a sensitivity analysis, models were further adjusted for maternal fish consumption during pregnancy, breastfeeding, and delivery by C-section (see Figure S2B).

#### Associations between POPs and gut microbiota markers

Third, we assessed associations between POP exposures and diversity and SCFAs using penalized and unpenalized linear regression models. These models were adjusted for maternal education, breastfeeding, delivery via C-section, and whether the child had recently received antibiotics (see Figure S2C). We also used multiple imputation for analyses based on microbiota data, and present estimates based on complete cases for comparison.

If there was a robust association between a POP and outcome which overlapped with associations with gut microbiota markers, we inputted these in an exploratory causal mediation analysis to assess if microbiota mediated the direct association (Pearl 2014) (as illustrated in Figure S3). We did not pursue a longitudinal approach given the large number of missing blocks across sampling time points.

We used QIIME (v.1.7.0.2) (Caporaso et al. 2010) and R software (v.3.2.3; R Foundation for Statistical Computing, Vienna, Austria) to perform statistical analyses, including the *glmnet* package for elastic net modeling (Friedman et al. 2010), and the *mice* imputation (Buuren and Groothuis-Oudshoorn 2011) and *mediation* (Tingley et al. 2014) packages.

## Results

### Study population

Characteristics of the study population (n=993) are presented in Table 1. 4.4% of children were registered as diagnosed with asthma by 2014. 7.0% were reported by the mother to have asthma at 2 years of age, of which 33.3% of these had a later diagnosis in the registry, and 19.9% had had an LRTI by 2 years of age. More than half of the mothers were older than 30 years when they became pregnant, had given birth before, and were within the normal weight range prior to pregnancy, and 11% smoked at the beginning of pregnancy. Nearly a quarter of children had received antibiotics by one year of age. Characteristics for the subset with fecal samples (n=438) are presented in Table S1, and were similar.

**Table 1.**
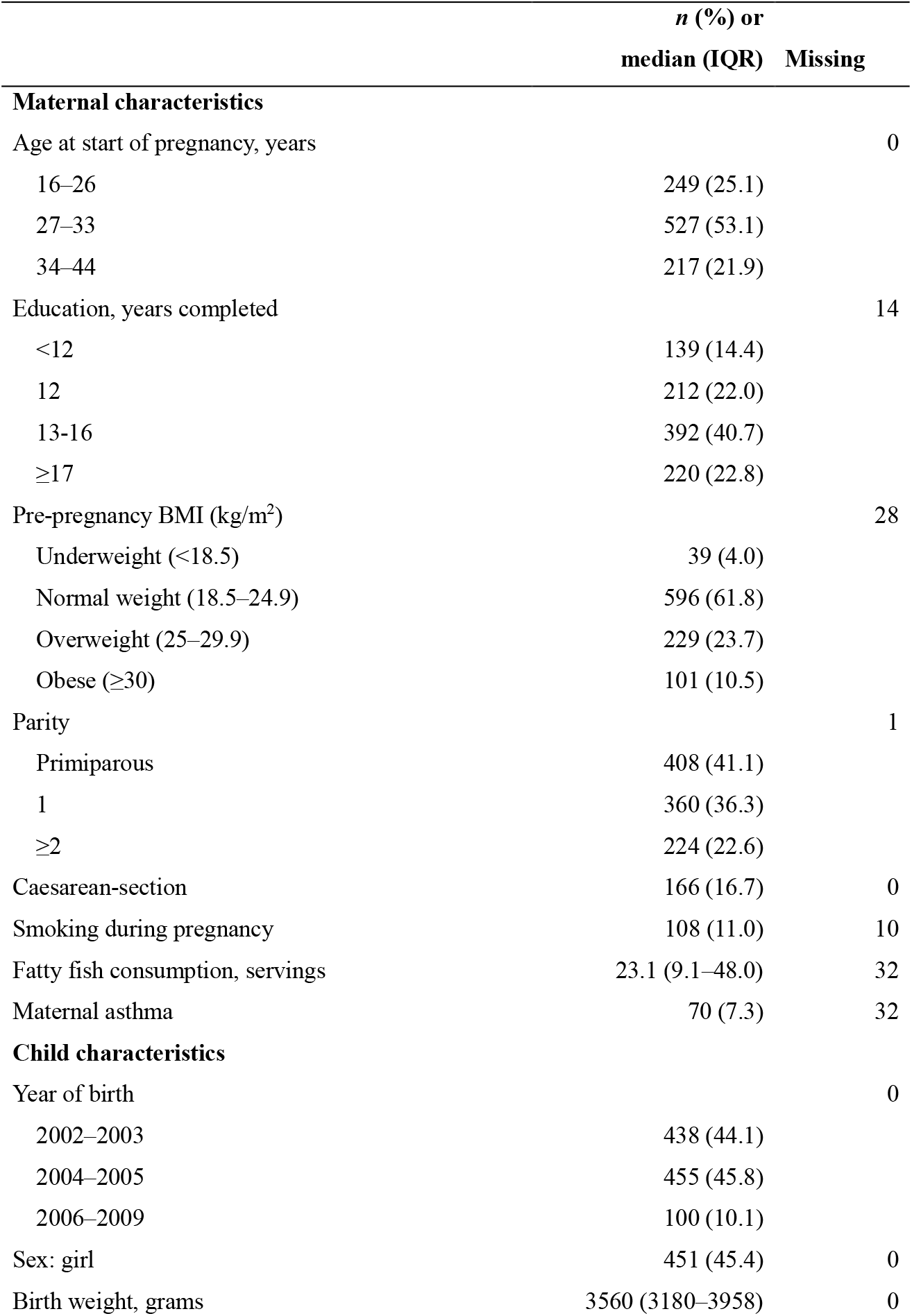
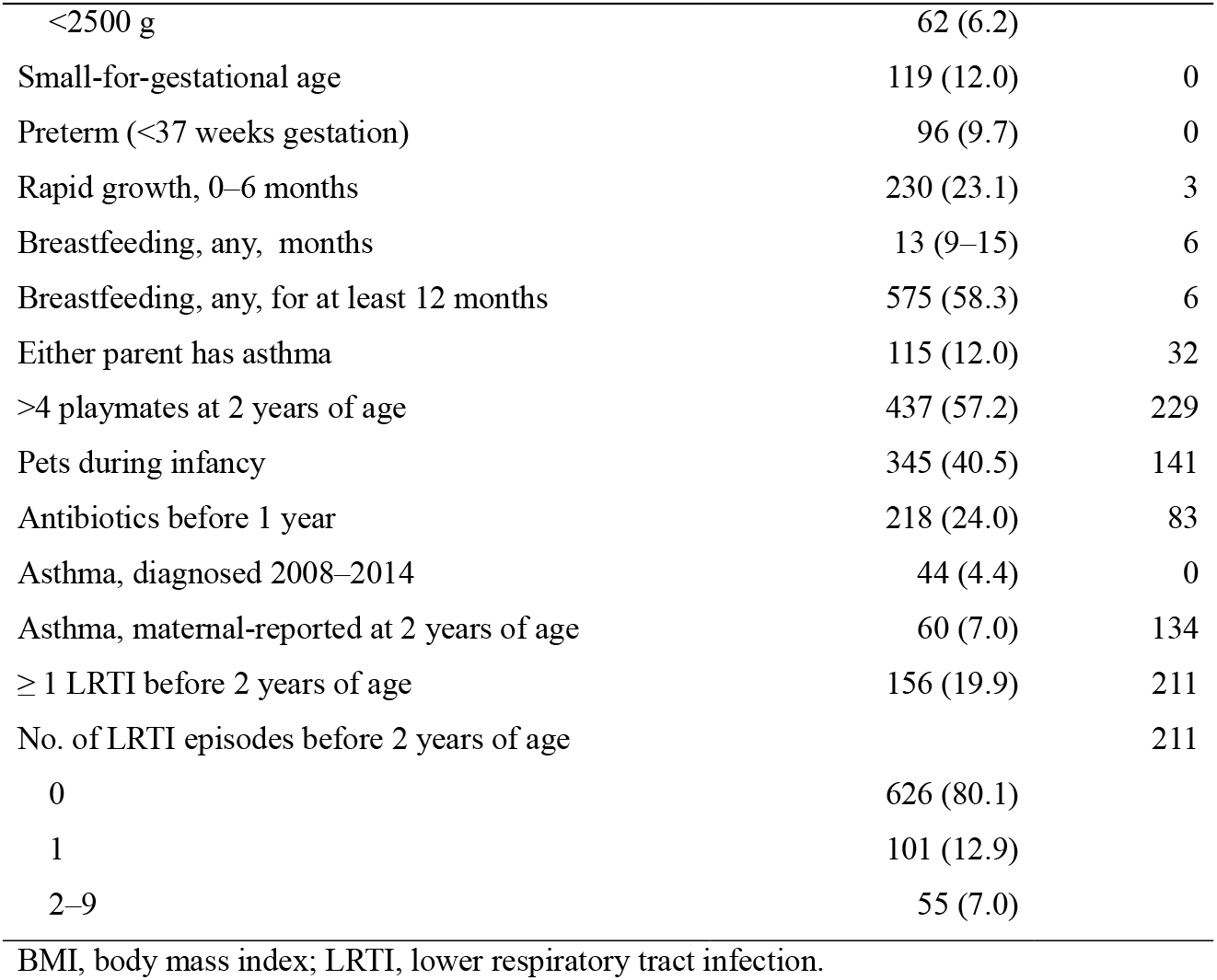
Study sample characteristics (HUMIS, Norway, n=993).

|  | *n* (%) or<br>median (IQR) | Missing |
| --- | --- | --- |
| **Maternal characteristics** |  |  |
| Age at start of pregnancy, years |  | 0 |
| 16–26 | 249 (25.1) |  |
| 27–33 | 527 (53.1) |  |
| 34–44 | 217 (21.9) |  |
| Education, years completed |  | 14 |
| <12 | 139 (14.4) |  |
| 12 | 212 (22.0) |  |
| 13–16 | 392 (40.7) |  |
| ≥17 | 220 (22.8) |  |
| Pre-pregnancy BMI (kg/m<sup>2</sup>) |  | 28 |
| Underweight (<18.5) | 39 (4.0) |  |
| Normal weight (18.5–24.9) | 596 (61.8) |  |
| Overweight (25–29.9) | 229 (23.7) |  |
| Obese (≥30) | 101 (10.5) |  |
| Parity |  | 1 |
| Primiparous | 408 (41.1) |  |
| 1 | 360 (36.3) |  |
| ≥2 | 224 (22.6) |  |
| Caesarean-section | 166 (16.7) | 0 |
| Smoking during pregnancy | 108 (11.0) | 10 |
| Fatty fish consumption, servings | 23.1 (9.1–48.0) | 32 |
| Maternal asthma | 70 (7.3) | 32 |
| **Child characteristics** |  |  |
| Year of birth |  | 0 |
| 2002–2003 | 438 (44.1) |  |
| 2004–2005 | 455 (45.8) |  |
| 2006–2009 | 100 (10.1) |  |
| Sex: girl | 451 (45.4) | 0 |
| Birth weight, grams | 3560 (3180–3958) | 0 |

|  | ***n* (%) or**<br>**median (IQR)** | **Missing** |
| --- | --- | --- |
| <2500 g | 62 (6.2) |  |
| Small-for-gestational age | 119 (12.0) | 0 |
| Preterm (<37 weeks gestation) | 96 (9.7) | 0 |
| Rapid growth, 0–6 months | 230 (23.1) | 3 |
| Breastfeeding, any, months | 13 (9–15) | 6 |
| Breastfeeding, any, for at least 12 months | 575 (58.3) | 6 |
| Either parent has asthma | 115 (12.0) | 32 |
| >4 playmates at 2 years of age | 437 (57.2) | 229 |
| Pets during infancy | 345 (40.5) | 141 |
| Antibiotics before 1 year | 218 (24.0) | 83 |
| Asthma, diagnosed 2008–2014 | 44 (4.4) | 0 |
| Asthma, maternal-reported at 2 years of age | 60 (7.0) | 134 |
| ≥ 1 LRTI before 2 years of age | 156 (19.9) | 211 |
| No. of LRTI episodes before 2 years of age |  | 211 |
| 0 | 626 (80.1) |  |
| 1 | 101 (12.9) |  |
| 2–9 | 55 (7.0) |  |

BMI, body mass index; LRTI, lower respiratory tract infection.

### Chemical exposures

Within each chemical class, the highest breast milk concentrations were observed for DDE (46.4 ng/g), PCB-153 followed by PCB-138 (32.2 and 19.9 ng/g, respectively), PBDE-47 (1.1 ng/g), and PFOS (110 ng/L) (Table S2). 37% of pairwise correlations (*r*_pearson_) exceeded 0.5, and 13% exceeded 0.8 (Figure S4). Correlations within each chemical class were moderate to high (an IQR of *r*_p_=0.65–0.82 for PCBs, 0.31–0.65 for OCPs, 0.63–0.94 for PBDEs, and *r*_p_=0.56 for the two PFASs). Correlations between PCBs and OCPs were moderate (*r*_p_=0.44 for ∑_14_PCBs and ∑_4_OCPs), whereas correlations between other chemical classes were generally low or non-significant.

### Gut microbiota metabolites and diversity

The three most abundant SCFAs were acetic, propionic, and butyric acid, comprising a median of >95% of the total SCFAs at all sampling points (Table S3). During the six sampling periods in the first 2 years of life, the proportion of acetic acid decreased and the proportion of other SCFAs increased. Shannon diversity increased over time, with a marked increase from 4 months to 12 months of age, whereas total SCFAs showed more modest increases with age (Figure S5). At 4 months, 1 and 2 years, diversity and total SCFAs were only weakly correlated across (*r*_s=_–0.08–0.24) and within sampling periods (*r*_s=_0.05–0.13). Individual SCFAs were generally moderately correlated within sampling periods, except *i*-butyric and *i*-valeric acid, which reflect protein fermentation (Smith and Macfarlane 1998), were highly correlated (*r*_s_ >0.9; Figure S5).

### Associations with covariates

Associations between exposures or outcomes and covariates were generally in the expected direction (Table S4). Having a parent with asthma was associated with an increased risk of asthma diagnosis. POP concentrations generally increased with maternal age, decreased with parity, and increased with earlier calendar year of sampling (child age).

### Associations between POPs and respiratory outcomes

Three exposures were consistently selected in penalized models as associated with registry-based asthma (when most children were around 10 years of age): β-HCH was associated with an increased risk, and PCB-118 and 138 were associated with a decreased risk (Figure 1 and Tables S5). The associations for β-HCH and the PCB-138 were the most precise in selected-subset multivariable regression model [odds ratio (OR)=2.99 (95% CI: 1.65, 5.43) per 2-SD increase in β-HCH; and OR=0.43 (95% CI: 0.20, 0.91) for PCB-138 (Table S8)], and both were selected in further adjusted models (Table S5).

**Figure 1.**
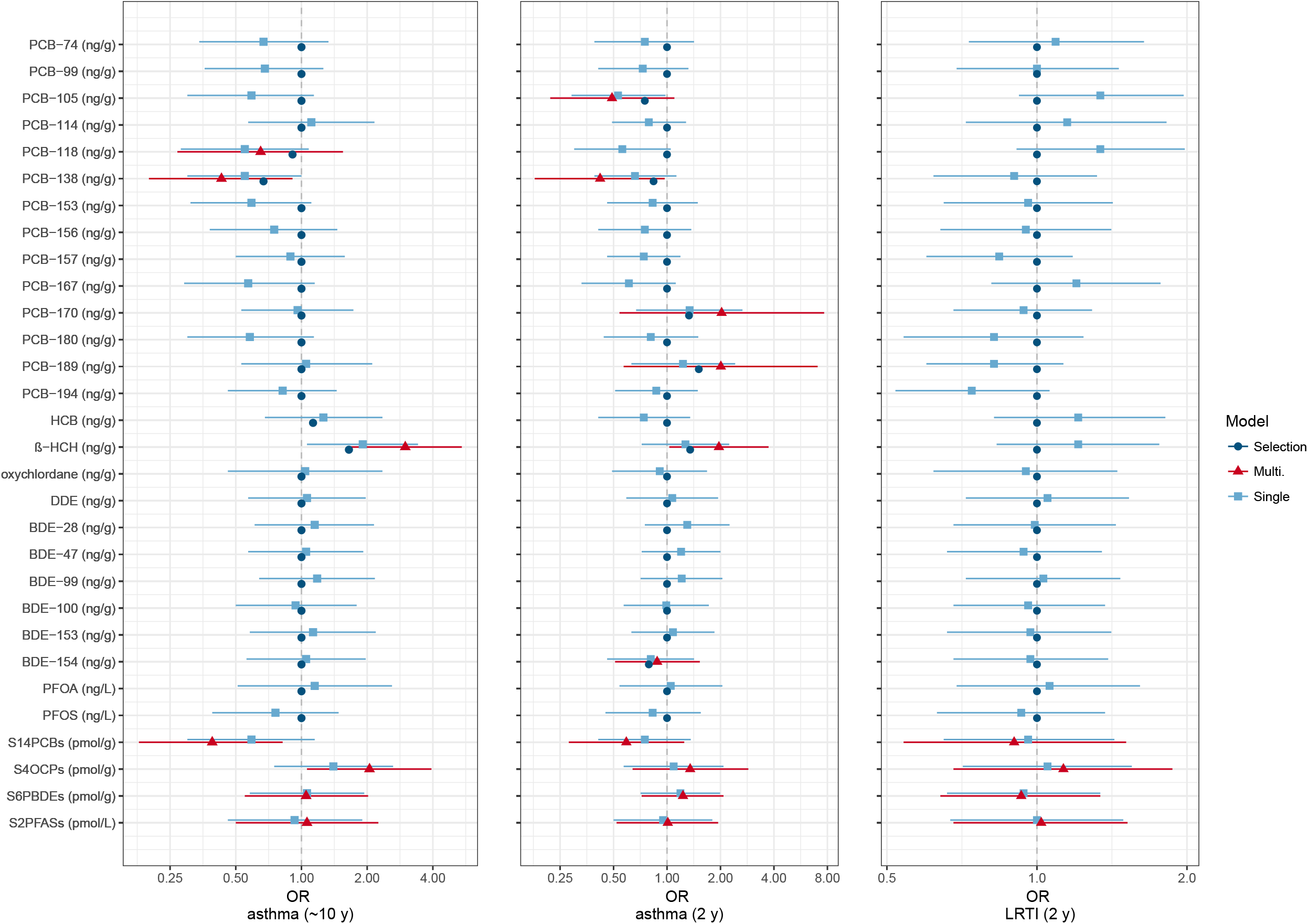
Odds ratios (OR) and 95% confidence intervals for the three respiratory health outcomes per 2-standard deviation increase in ln-transformed exposure concentrations. Multivariable elastic net penalized logistic regression selection and coefficients (dark blue; circle), and the selected subset of chemicals remodeled in a multivariable logistic regression model (red; triangle) are presented. For comparison, the single-pollutant (unpenalized) logistic regression coefficients (light blue; square) are also presented. Coefficients from single- and multivariable logistic regression models are also presented for the summed chemical groups. Models were adjusted for maternal age, parity, pre-pregnancy BMI, and maternal education; missing data was multiply imputed (n=993; exposure increments and numerical results are provided in Tables S5–S8).

Six exposures were associated with maternal-reported asthma at 2 years of age (Table S6). The same direction of associations were observed for β-HCH and PCB-138, and the estimates for PCB-138 (OR=0.42; 95% CI: 0.18, 0.97) and β-HCH (OR=1.96; 95% CI: 1.03, 3.73) were similarly most precise. In addition, PCB-170 and 189 were associated with an increased risk and PCB-105 and BDE-154 with a decreased risk of maternally reported asthma at 2 years of age, although variable selection results were less stable across penalized models than those for registry-based asthma by 10 years, and none of these exposures were consistently selected in further adjusted models (Table S6).

Associations with summed chemical groups showed the same directions of effect for both asthma outcomes, although estimates were also more precise for registry-based asthma at 10 years (OR=2.05; 95% CI: 1.06, 3.94 per 2-SD increase in ∑_4_OCPs and OR=0.39; 95% CI: 0.18, 0.82 for ∑_14_PCBs; Table S8) than for maternal-reported asthma at 2 years.

No exposures were associated with occurrence of an LRTI by 2 years. None of the single-pollutant–asthma associations survived correction for multiple testing. Results based on multiple imputation and complete case analyses were generally similar (Tables S5–S8). There was some evidence of effect modification by child sex for PCB-138 and maternal-reported asthma at 2 years; a positive association for girls and a negative association for boys. There was little evidence that other associations were modified by child sex (Table S8), or by maternal smoking or education (data not shown).

#### Associations between gut microbiota markers and respiratory outcomes

Shannon diversity at 4 months of age was positively associated with asthma ascertained both at age 2 and 10 years (Figure 2 and Table S9–S10). Diversity at 12 months of age was inversely associated, although imprecisely, with maternal-reported asthma at 2 years and occurrence of an LRTI by 2 years. Other estimates for diversity were closer to null. Associations for total SCFAs and most individual SCFAs were imprecise, and generally inconsistent in direction across the three early-life sampling time points. *i*-valeric acid at 12 months of age was positively associated with asthma at 10 years. Propionic and *i*-butyric acid at 2 years were associated with decreased risk of an LRTI by 2 years. Estimates for *i*-valeric and *i*-butyric acid were inconsistent in direction in some univariable versus multivariable regression analyses, likely due to their very high collinearity. In a sensitivity analysis with SCFAs analyzed compositionally as log-contrasts and compared to reduced models only including confounders, only the inverse association between SCFAs at 2 years and LRTI was significant (likelihood ratio test *p*=0.01). Regression coefficients based on multiply imputed data were somewhat inconsistent with complete case analyses, particularly for weak and imprecise associations, and for the microbiota markers for which a large fraction was imputed (i.e., SCFAs at 4 months and diversity at 2 years, for which n<100 for covariate-adjusted associations).

**Figure 2.**
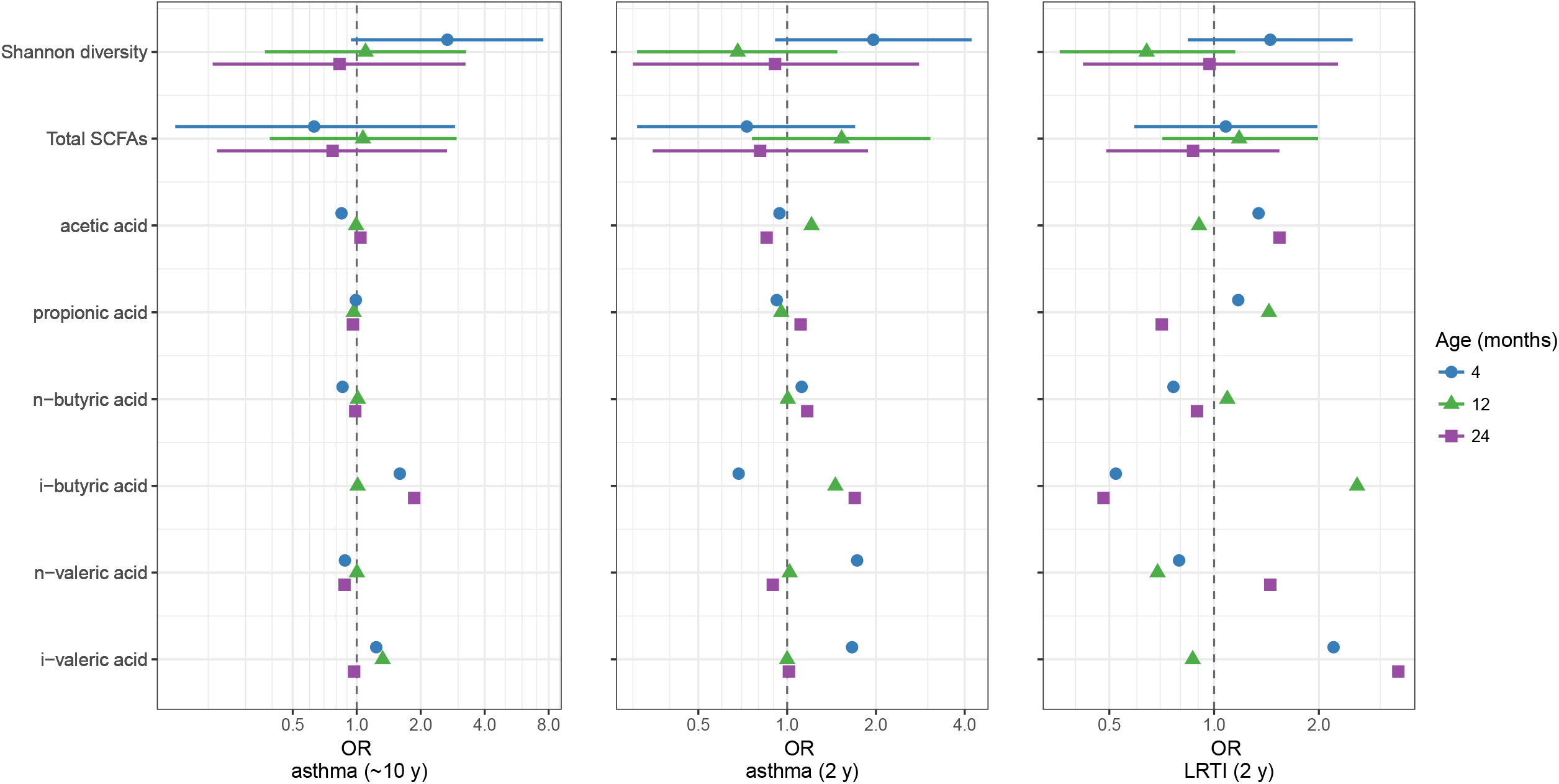
Odds ratios (OR) and 95% confidence intervals for the three respiratory health outcomes per 2-standard deviation increase in Shannon diversity and SCFA levels (mmol/kg) across 3 sampling time periods in early life. Logistic regression coefficients are presented for Shannon diversity and total SCFAs, and multivariable elastic net penalized logistic regression coefficients are presented for the individual SCFAs; models were estimated using multiply imputed data (n=438). Models were adjusted for preterm birth, maternal smoking during and after pregnancy, and maternal education (increments and numerical results for penalized and unpenalized models are provided in Tables S9–S10).

#### Associations between POPs and gut microbiota markers

∑_14_PCBs were associated with reduced Shannon diversity at 2 years (Figure 3; Table S11). This appeared to be driven by PCB-114, based on selection within penalized regression models (Table S12). ∑_4_OCPs was imprecisely associated with diversity, both inversely at 1 year and positively at 2 years of age; β-HCH was selected at both time points in penalized models. ∑_14_PCBs was positively associated with total SCFAs at 4 months and 1 year, and ∑_4_OCPs was inversely associated with total SCFAs at 2 years, although associations with total SCFAs were imprecise. Associations were close to null for ∑_6_PBDEs and ∑_2_PFASs, except for an inverse association between ∑_2_PFASs and total SCFAs at 2 years, which was seemingly driven by an inverse association between PFOA and acetic acid (Table S12). Regression coefficients were generally attenuated and less precise for analyses based on multiply imputed compared to complete case data.

**Figure 3.**
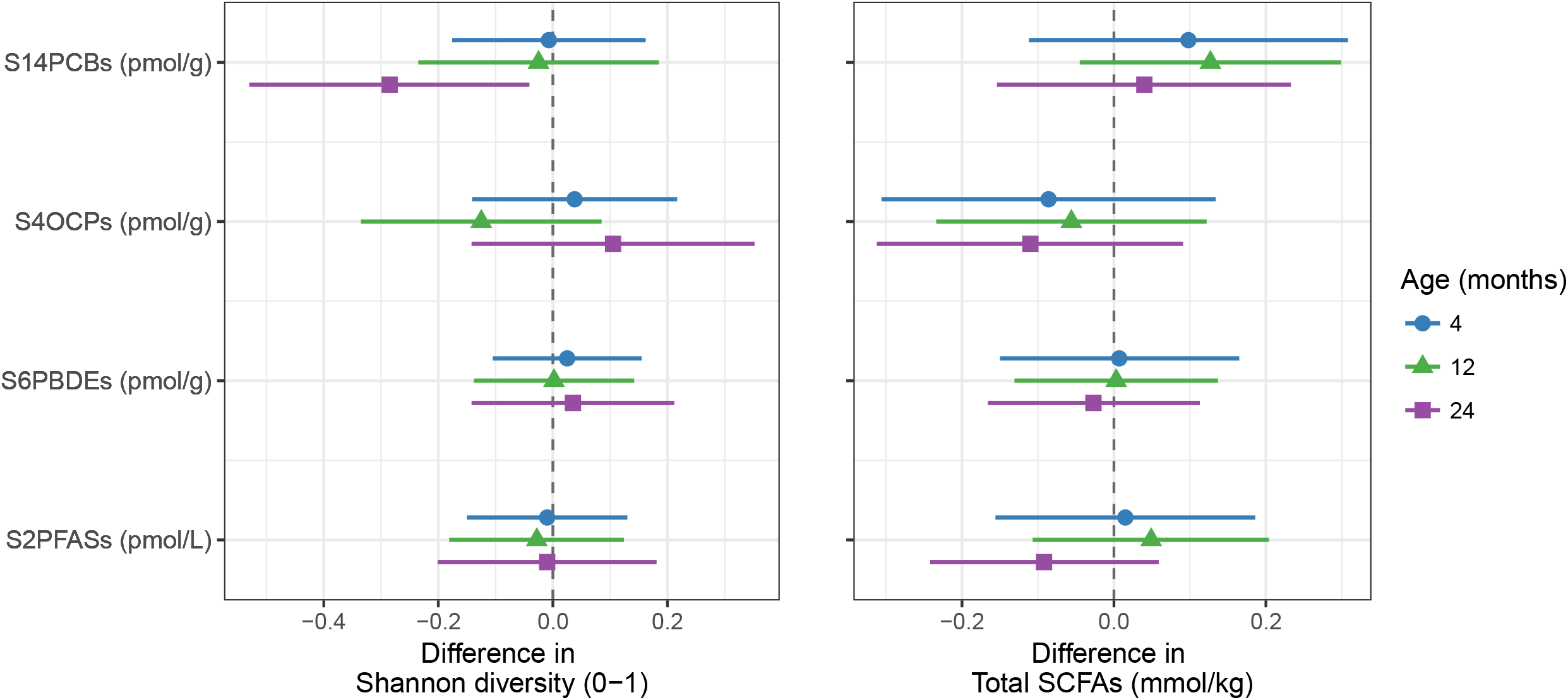
Differences in 2-standard deviations of Shannon diversity and total SCFA concentrations across 3 sampling time periods in early life associated with a 2-standard deviation increase in natural-log transformed exposure concentrations. Multivariable logistic regression coefficients are presented, estimated based on multiply imputed data (n=298). Models were adjusted for maternal education, breastfeeding, C-section, and recent antibiotics (numerical results for unpenalized models, including results for individual SCFAs, are provided in Table S11 and for individual POPs based on penalized models in Table S12

### Mediation analyses

We tested if the β-HCH and asthma association was mediated by total SCFAs at 4 months, or if the ∑DL-PCBs and asthma association was mediated by Shannon diversity at 4 months. Neither path showed a significant mediation; i.e., the confidence intervals of the natural indirect (mediated) effects included zero (Figure S3).

## Discussion

In this prospective cohort study, we observed that β-HCH was consistently associated with an increased odds of asthma at 2 years of age and later childhood asthma. Several PCBs were positively and others inversely associated with asthma, including a robust inverse association between the non-dioxin-like PCB-138. We did not observe consistent associations between POPs and LRTIs. We observed both positive and inverse associations between gut microbial diversity and later asthma, and also for several SCFAs, although effect estimates were generally less stable and imprecise. NDL and DL-PCBs were associated with reduced microbial diversity at 2 years of age, and OCPs with lower SCFAs at 2 years of age. Microbial diversity and metabolites did not appear to mediate the POP– asthma associations.

### POPs and respiratory outcomes

We found associations for β-HCH and PCB-138, highly persistent legacy POPs which have shown declining serum concentrations in the past three decades in Norway (Nost et al. 2013). β-HCH is one of the isomers of the technical mixture HCH, which along with isolated γ-HCH (known as Lindane), was widely used as an insecticide until the 1990s (Vijgen et al. 2011). To our knowledge, this is the first prospective study to assess HCH exposure in relation to risk of asthma. A prospective Danish study (n=872) that assessed maternal serum levels in association with offspring asthma medication use between 6 and 20 years of age reported a positive association with the organochlorine fungicide HCB, and a null association with DDE (Hansen et al. 2014). This study also found positive associations with DL-PCB-118 and weak positive associations with five other PCBs tested. We found both inverse and weak positive associations between PCBs and asthma, but the most robust association was an inverse association between with non-dioxin-like PCB-138. In the same Danish study population, PCBs, HCB, and DDE were also associated with airway obstruction, but not allergic sensitization (Hansen et al. 2016).

This relatively large study did not confirm findings of a meta-analysis of 10 European birth cohorts (n=4608) which reported a robust positive association between prenatal DDE exposure and ever-bronchitis and ever-wheeze by around 18 months, and a weaker and inconsistent association with PCB-153 (Gascon et al. 2014). This meta-analysis included the present study population (HUMIS), although a smaller sample size was then available with measured POP concentrations (n=386 versus 993 now). Part of the discrepancy may be attributable to the lower levels of prenatal exposure to POPs in our study population and Norway in general, compared to other European countries (van den Berg et al. 2017). Other, smaller studies have reported both positive (Dallaire et al. 2006; Grandjean et al. 2010; Stolevik et al. 2011; Weisglas-Kuperus et al. 2004) and inverse associations (Grandjean et al. 2010; Weisglas-Kuperus et al. 2004) between early-life PCB and OCP exposures and markers of allergic and respiratory diseases, including wheeze, eczema, IgE, and upper and lower RTIs. It has been postulated (Stolevik et al. 2011) that early-life exposure to some PCBs, specifically DL-PCBs, may reduce the risk of developing allergic diseases through aryl hydrocarbon receptor (AhR)-mediated immunosuppression (Quintana et al. 2008).

We did not identify associations between PFOS or PFOA and childhood asthma. A cross-sectional analysis in the US (NHANES, n=1,877) reported a positive association for PFOA, an inverse association for PFOS, and null results for two other PFASs (Humblet et al. 2014). Two prospective birth cohort studies in Norway reported associations between prenatal exposure to multiple PFASs and increased respiratory tract infections, and largely null findings for asthma (n=641 and n=921/1207) (Impinen et al. 2018; Impinen et al. 2019). Prenatal PFAS exposure has been linked to reduced vaccine antibody titers in childhood, providing suggestive evidence that PFASs exert immunosuppressive effects (Grandjean et al. 2012; Granum et al. 2013; von Holst et al. 2021). A few small case-control studies have assessed levels of PBDEs in indoor dust in relation to asthma. A prospective study reported null findings for multiple PBDEs and organophosphate flame retardants (Canbaz et al. 2016), and a cross-sectional study reported a weak inverse association with BDE-99 and null findings for BDE-47 and 209 (Meng et al. 2016).

### Diversity/SCFAs and respiratory outcomes

In a seminal study, Arrieta et al. (2015) examined early-life gut microbiota composition and SCFAs in relation to wheezing and skin atopy at age 1 year in Canadian infants. Although the sample size for the SCFAs analysis was small (n=13 cases with atopy and wheeze and n=13 controls), they observed reduced acetic acid levels at 3 months of age for cases. Furthermore, in a larger sample size (n=324), they found that compositional dysbiosis (of the genera *Faecalibacterium, Lachnospira, Veillonella*, and *Rothia)*, but not diversity, was associate with risk of asthma and allergic disease. In a prospective cohort study in Denmark, α-diversity in early-life was not associated with childhood asthma (n=648; 60 asthma cases at age 5 years), although for children born to asthmatic mothers, microbiota composition and population structure at 1 year was associated with asthma (Stokholm et al. 2018). A small study (n=47) in Sweden previously observed that Shannon diversity at 1 week and 1 month, but not 12 months, was inversely associated with asthma at 7 years of age; diversity was not associated with other atopic disease markers (allergic rhinoconjunctivitis, eczema or skin prick reactivity) (Abrahamsson et al. 2014). A larger study of 411 children in Denmark found that gut microbiota diversity at 1 month and 12 months of age was inversely associated with several markers of allergic sensitization, but not asthma (Bisgaard et al. 2011). A study of 917 Canadian found that gut microbiota α-diversity at 12 months of age was associated with a reduced risk of asthma at age 5 years (Patrick et al. 2020).

We observed the strongest associations for gut microbiota diversity at 4 months and increased risk of asthma, counter to the *a priori* hypothesis that diversity is protective. However, high diversity in very early life is tied to lack of breastfeeding (Backhed et al. 2015) and thus at this age high diversity may not be beneficial. The different sample sizes with data available on microbial markers hampered our ability to compare coefficients and identify sensitive windows of exposure. A recent review of mouse-model and human studies on microbial immune system priming and allergic disease development concluded that the first 100 days may be the most important early-life critical window (Stiemsma and Turvey 2017). A lack of diversity in early-life may have longer-term consequences for host immunity, whereas SCFAs may have more immediate effects of suppressing immune responses and asthma symptoms. This is supported by a randomized control trial in asthmatics that found a reduction in clinical symptoms and cytokine production following probiotic supplementation with *Lactobacillus gasseri* (Chen et al. 2010), and by experimental mouse models, in which increased acetic acid resulting from a high-fiber diet suppressed airway inflammation via transcription of Foxp3, and promotion of T-regulatory cells (Thorburn et al. 2015). SCFAs have generally been shown to exert anti-inflammatory effects, inhibiting histone deacetylase (HDAC) and inducing T-regulatory cells, and acting as predominantly agonists of G protein-coupled receptors. Early-life nasopharyngeal (upper respiratory tract) microbiota compositional changes (Teo et al. 2015) have also been found to antedate asthma development. It is unclear if pathways are similar for gut and lung microbiota and immune system signaling.

Maternal and child antibiotic use has been associated with subsequent asthma, (Foliaki et al. 2009; Loewen et al. 2018) and this has been supported by experimental evidence from animal models (Russell et al. 2012). This has been attributed to gut microbiota dysbiosis of the child, as antibiotics have been shown to lead to both rapid transient and long-lasting changes to gut and lung microbiota (Korpela et al. 2016), and maternal microbiota influence offspring composition through vertical transmission. However, antibiotics prescribed for non-respiratory illnesses in early life were not associated with an increased risk of asthma (Ortqvist et al. 2014). The causal mechanisms of the antibiotic–asthma association requires further research, and may be partly attributable to common risk factors and underlying susceptibility (Stokholm et al. 2014).

### POPs and diversity/SCFAs

We found limited evidence that early-life exposure to PCBs and OCPs perturbed gut microbiota diversity or SCFA production. ∑_14_PCBs showed a robust association with lower diversity at 2 years of age; however, other associations were closer to null or less precise. We previously reported that several POPs, including PBDE-28, PFOA, PFOS, and PCB-167, were associated microbiota compositional differences at 1 month of age in the present study population (Iszatt et al. 2019). Recent experimental studies support that the interplay between POPs and microbiota is mediated by the AhR (Zhang et al. 2015); POPs are potent ligands of the AhR, and AhR activity can modulate microbiota, and conversely, microbial metabolites can activate the AhR, which in turns mediates the toxicity of xenobiotics (Zhang et al. 2017).

### Strengths and limitations

To our knowledge, this study included the largest number of chemicals evaluated simultaneously in relation to respiratory and allergic diseases, and is one of the largest studies to date. We used a multi-pollutant approach to reduce confounding bias due to correlated co-exposures (Lenters et al. 2018). Another strength of this study is that detailed data on important potential confounders, such as breastfeeding and antibiotic use, was available. Breast milk levels of POPs served as a proxy of early-life offspring exposures, and directly reflect the dose via breastfeeding. We did not attempt to model temporal variability in prenatal and postnatal exposure levels over time, which are highly correlated. Absolute levels of *in utero* exposures to protein-bound PFASs are underestimated by breast milk levels, as transfer of protein-bound PFASs from maternal serum into breast milk is less efficient than for lipophilic POPs (Cariou et al. 2015), although the relative ranking would be expected to remain the same.

This study has limitations. Reliance on questionnaire data led to a loss to follow-up in maternal-reported outcomes for the child at age 2 (13% for asthma and 21% for LRTI), and thereby a loss of statistical power and potential selection bias; this was not a problem for the registry-based asthma outcome. Based on prevalence studies of asthma in Norway, it is likely that the national register captured severe cases, but less severe asthma or chronic wheeze cases, along with children who were younger at the time of linkage, were more likely to have been misclassified as non-asthmatic. Differential misclassification of outcomes with respect to chemical exposures is possible, in that accuracy of maternal-reported outcomes and likelihood of visiting the hospital may differ somewhat with socioeconomic position, and exposures may too; however, associations with maternal education were non-significant in this study population, and these biases are expected to be minimal. Prevalence of asthma in Norwegian population-based studies is heterogeneous; ever-asthma (lifetime asthma) based on parental-reports and medications in 10 years olds recruited in Oslo was 20.2% (Lodrup Carlsen et al. 2006); current asthma was 5.7% at 3 years and 5.1% at 7 years of age in a large Norwegian birth cohort, ascertained based on parental-report in combination with asthma medication use (Magnus et al. 2015).

We cannot exclude that effect estimates suffer from residual confounding due to unmeasured exposures. A small number of prospective studies provide suggestive, although limited, evidence that early-life exposure to other environmental chemicals may be associated with asthma and other allergic diseases, including certain antimicrobial compounds, organophosphate pesticides, phthalates, and bisphenol A (Gascon et al. 2015; Jackson-Browne et al. 2019; Ku et al. 2015; Raanan et al. 2015; Smit et al. 2015; Spanier et al. 2014; Whyatt et al. 2014); however, the magnitude of potential confounding bias is expected to be small given that these non-persistent exposures generally exhibit low correlation levels with the POPs considered in the present study (Robinson et al. 2015).

Furthermore, we cannot preclude that the mediation analyses did not meet the assumption of no residual confounding of the various relationships, including confounding of the mediator–outcome relationship (Figure S3), nor the presumed temporal ordering of the POP–microbial markers associations, which may exhibit bidirectional effects (Claus et al. 2016). With a more complete matrix of repeated measures of gut microbiota markers, we would have pursued longitudinal modeling which has the advantage of increased statistical power. We did not study phylogenetic composition as it was beyond the scope of this study; however, it has been postulated that diversity rather than single species are most relevant for optimal immune system development (Gensollen et al. 2016; Lynch et al. 2017).

In conclusion, we found evidence that some organochlorine pesticides were associated with increased risk of developing asthma, and also observed that several PCBs were inversely associated with asthma. The present study provides weak support for the hypothesis that reduced gut microbial diversity and reduced production of microbial metabolites predispose asthma development. Although our study did not identify consistent evidence that gut microbiota diversity and function modulated associations between environmental chemicals and asthma, the interplay between chemical exposures, gut microbiota, and allergic disease warrants further research.

## Data Availability

Data cannot be made publicly available as the dataset contains sensitive and identifying information. The data may be made available upon request.

## Acknowledgments

This research was supported in part by the Research Council of Norway (NevroNor, grant agreement no. 226402) and by the European Commission’s Seventh Framework Program (DENAMIC project, grant agreement FP7-ENV-2011-282957).

