## Supplementary Material for "Early-life exposure to persistent organic pollutants, gut microbiota diversity and metabolites, and respiratory health in Norwegian children"

### Appendix S1. Supplemental Methods

#### Study population

Recruitment and eligibility criteria: Participants were eligible to participate if they spoke fluent Norwegian and were a resident of the county in which they were recruited. The majority of participants were from Finnmark, Oppland, Østfold, Rogaland, Telemark, and Troms, and a small number (<20 of the 993 sample included in the present study) were from Akershus and Nordland. These counties cover northern, southern, inland, and coastal areas of Norway. 868 (98.1%) of the parents were both white, and the others were both Asian (0.8%), were both Inuit (0.7%), or another combination (0.4%; or this data was missing, 10.6%).

Breast milk sample collection and handling: Mothers who consented to participate received pre-washed containers in which to collect their breast milk. They were asked to collect 25 mL of breast milk by hand in the morning on 8 consecutive days before the child reached 2 months of age. Minor deviations in this sampling protocol, such as the total number of sampling days and collection by breast pump, were accepted. 55% of women collected breast milk for fewer than 8 days. The median number of days over which breast milk was collected was 8 [interquartile range (IQR), 5–8 days; data missing for 19%]. 32% reported using a pump instead of hand milking. Milk samples were transferred to a 250 mL container and were kept frozen after collection; they were allowed to thaw as they were sent by regular mail to the coordinating center, the Norwegian Institute of Public Health (NIPH), Oslo, Norway. The procedure differed for Østfold County, where samples were collected by study personnel and were kept frozen until transport. Samples were stored at NIPH at −20°C until analyzed.

Fecal sample collection and handling: Mothers who consented to participate received containers for fecal sample collection at the maternity ward. Study personnel retrieved maternal and child fecal samples, and kept them frozen during transport. Samples were stored at −20°C at NIPH until analyzed.

#### Chemical exposure assessment

Analytical protocols and quality control (QC) measures have been described in detail elsewhere. We provide the relevant references and briefly describe the protocols here. To date, up to n=1028 breast milk samples (specifically, aliquots from the samples pooled for each individual) have been assayed for some chemicals. We describe the methods for the set of chemicals measured and detected in ≥70% of samples for the study sample used in the present study (n=993).

PFASs: We used data for PFOS and PFOA, which were measured in 884 samples of the 993, as previously described (Forns et al. 2015). Breast milk samples (n~80%) were analyzed for PFASs at the Department of Environmental Exposure and Epidemiology (formerly the Department of Analytical Chemistry), NIPH (Oslo, Norway), using high performance liquid chromatography/tandem mass spectrometry (LC-MS/MS) according to a previously described protocol (Haug et al. 2009; Thomsen et al. 2010a). A second batch of samples (n~20%) were analyzed at the Institute for Environmental Studies (IVM), Faculty of Earth and Life Sciences, VU University (Amsterdam, the Netherlands), using LC-MS/MS on a triple quadrupole mass spectrometer following a previously described protocol (de Cock et al. 2014).

PBDEs: We used data for six polybrominated diphenyl ethers (PBDE) congeners (BDE-28, 47, 99, 100, 153, and 154) which were measured in 957 samples of the 993. One batch (n~40%) was measured at NIPH using automated solid-phase extraction (SPE) extraction and a gas chromatography–mass spectrometry (GC–MS) system, as previously described (Thomsen et al. 2007; Thomsen et al. 2010b). The PBDE standards were obtained from Cambridge Isotope Laboratories (Andover, MA, USA). A second batch (n~60%) was subsequently analyzed at NIPH. The same SPE extraction procedure was used (Thomsen et al. 2010b), while a GC–high resolution MS (GC-HRMS) with electron impact ionization was used for quantification (Caspersen et al. 2016; Frederiksen et al. 2010).

PCBs and OCPs: Fourteen polychlorinated biphenyls [PCBs; non-dioxin-like (74, 99, 138, 153, 170, 180, and 194), and dioxin-like (105, 114, 118, 156, 157, 167, and 189)] and 4 organochlorine pesticides [(OCPs): hexachlorobenzene (HCB), β-hexachlorocyclohexane (β-HCH), 4,4′dichlorodiphenyldichloroethylene (*p,p′*-DDE), and oxychlordane] were measured in two batches. The first batch (n~50%) was measured at the Norwegian University of Life Sciences (formerly the Norwegian School of Veterinary Science; Oslo, Norway), using GC-electron capture detector (GC-ECD) for OCPs and non-dioxin like PCBs, and using GC-low resolution MS (GC–LRMS) for the dioxin-like mono-*ortho* PCBs, as previously described (Brevik 1978; Eggesbø et al. 2009; Polder et al. 2008; Polder et al. 2009). A second batch of samples (n~50%) were measured (at the same time as the second batch analysis of PBDEs) at NIPH using the same SPE extraction procedure (Thomsen et al. 2010b), and GC-HRMS as described in (Caspersen et al. 2016; Forns et al. 2016).

Evaluating potential for batch effects: The subsets of samples analyzed were oversampled for outcomes which might be related to the outcomes assessed in the present study (n=157 were oversampled for preterm birth, small for gestational age, or rapid growth). We tested for systematic differences in measured levels between analysis batches ('batch effects') using principal component analysis (PCA) visualization and linear regression models, adjusted for birth year, maternal age, parity and oversampling-stratum. We found no clear evidence of batch effects, and therefore did not attempt to apply normalization methods to correct for batch effects.

**Analysis of fecal SCFAs**

Short-chain fatty acids (SCFAs) are saturated aliphatic organic acids with fewer than 6 carbon atoms. SCFAs in fecal samples represent the fraction unabsorbed in the gastrointestinal tract. The following fatty acids were analyzed: acetic ($C_2$), propionic (or the IUPAC name, propanoic) ($C_3$), *iso*-butyric (i-$C_4$), *n*-butyric ($C_4$) (or butanoic), *iso*-valeric (i-$C_5$), *n*-valeric (or pentanoic) ($C_5$), *iso*-caproic (i-$C_6$), and *n*-caproic (or hexanoic) ($C_6$) acids. Caproic acid is often classified as a medium-chain fatty acid (MCFA). Other SCFAs, e.g., succinic acid, lactic acid and formic acid, can be detected with lower frequencies, and were not measured in the present study. The majority of SCFAs are produced by carbohydrate fermentation (saccharolytic fermentation), although SCFAs are also produced by protein breakdown and amino acid fermentation (proteolytic fermentation). The branched-chain fatty acids (BCFA), i.e. *i*-butyric, *i*-valeric, *i*-caproic and also 2-methylbutyrate, are principally produced by amino acid fermentation (specifically valine, leucine and isoleucine) (Smith and Macfarlane 1997, 1998).

All fecal samples with a sufficient volume for the analysis (0.5 g) were analyzed. Two batches (n = 734 and n = 65) were analyzed at the Unger-Vetlesen Institute, Lovisenberg Diaconal Hospital (Oslo, Norway), with a limit of detection of 0.06 mmol/kg for all SCFAs. The fecal material (0.5 g) was homogenized after addition of distilled water containing 3 mmol/L of 2-ethylbutyric acid (as internal standard) and 0.5 mmol/L of $H_2SO_4$; 2.5 mL of the homogenate was vacuum distilled, according to the method of Zijlstra et al. (1977), as modified by Høverstad et al. (1984). The distillate was analyzed with gas chromatography (Agilent 7890 A, Agilent Technologies, Santa Clara, CA, USA), using a capillary column (serial no. USE400345H, Agilent J&W GC columns, CA, USA), and quantified using internal standardization. Flame ionization detection was employed. A third batch (n=336) was analyzed at the Department of Microbiology, Tumor and Cell Biology, Karolinska Institute (Stockholm, Sweden) using the same protocol (Midtvedt et al. 1988), with an LOD of 0.03 mmol/kg. SCFAs were assessed on a wet weight basis (mmol/kg).

**Analysis of gut microbiota diversity**

We calculated the Shannon diversity index, a measure of within sample (alpha) diversity: $H = -\Sigma p_i \ln(p_i)$, where $p_i$ denotes the relative frequency of operational taxonomic unit (OTU) *i.* Not all samples were analyzed, for several reasons, partly due to an industrial production error of the PCR plates used.

<u>DNA extraction</u>: Analysis of OTUs was performed at the Knight Lab at the Department of Chemistry and Biochemistry, University of Colorado (Boulder, Colorado, USA; now at the University of California, San Diego). First fecal samples were prepared for analysis by adding 1 mL Solution 1 (50 mM glucose, 25 mM Tris–HCl pH 8.0, and 10 mM EDTA pH 8.0) per 0.2 g feces. The samples were mixed by vortexing and left for 30–60 min on ice before 400 µl of the supernatant was diluted 1:2 in 4 M guanidinium thiocyanate (GTC). Five hundred microliters of sample were transferred to a sterile FastPrep®-tube (Qbiogene Inc., Carlsbad, CA, USA) containing 250 mg glass beads (106 microns and finer, Sigma-Aldrich, Steinheim, Germany), and samples were homogenized for 40 seconds in FastPrep® Instrument (Qbiogene). Wells in a 96-well Greiner U-plate (Greiner bio-one, Frickenhausen, Germany) were filled with 170 µL sample and 10 µL Silica particles (Merck, Darmstadt, Germany) and transferred to a Biomek® 2000 Workstation (Beckman Coulter, Fullerton, CA, USA). One percent Sarkosyl was added, and the plate was incubated at 65°C for 10 min. and at room temperature for 10 min. The supernatant was removed, and the paramagnetic beads were washed twice with 50% ethanol. DNA was eluted from the silica particles by suspension of the particles in 100 µL Buffer C (1 mM EDTA pH 8.0, 10 mM Tris–HCl pH 8·0) at 65°C for 30 min. The adequacy of the automated DNA extraction procedure was evaluated by repeating the DNA extraction in 20 samples using the modified MoBio 96-well manual extraction method adopted by the Earth Microbiome Project (http://press.igsb.anl.gov/earthmicrobiome/emp-standard-protocols/dna-extraction-protocol/). The samples gave very similar results regardless of DNA extraction method used.

<u>PCR amplification</u>: 1 µL DNA extracted from fecal samples was amplified by PCR reactions by 16S specific primers (515F-806R) (http://www.earthmicrobiome.org/emp-standard-protocols/16s/). All reactions were set up as 25 µL samples in 96 well Thermo-fast 96, low profile, 0.2 mL, non-skirted PCR plates (ABgene Thermo scientific, UK) with Cas1200 Corbett robot (Qiagen). 10 µL HotMastermix enzyme (5PRIME GmbH, Germany), 0.2 µM forward-/ reverse primers (ILHS_515fa/ IL_806rcbc) and 13 µL PCR grade water (Qiagen) were used.

<u>Sequence processing</u>: Sequencing of the V4 region of the 16S ribosomal RNA (rRNA) gene using the Illumina HiSeq instrument (Caporaso et al. 2012) resulted in a total of 2,546 samples for all time points

after demultiplexing, and quality-filtering (excluding samples of poor quality, with number of OTUs <1000). All data processing was performed using Quantitative Insights Into Microbial Ecology (QIIME) pipeline version 1.7.0.2 (Caporaso et al. 2010). OTUs were assigned to bacterial taxa using a closed-reference OTU picking procedure (97% sequence similarity) and mapped against the Greengenes (release 13-8) reference database of 16S rRNA sequences (Kuczynski et al. 2012). For data based on diversity, data were rarefied to 5906 sequences per sample, the minimum number of reads (OTU count) observed in all samples. The median number was 94 229 for 4 months, 124 489 for 12 months, and 152 266 for 2 years.

**Description of the multiple imputation procedure**

- Method: Multiple imputation by chained equations; estimates combined using Rubin's rules
- Software: *mice* package in R (Buuren and Groothuis-Oudshoorn 2011)
- 3 sets of 100 imputed datasets were created: (1) chemicals–respiratory outcomes (n=993); (2) microbiota diversity and SCFAs–respiratory outcomes (n=438); and (3) chemicals–microbiota diversity and SCFAs (n=298)
- Variables included in the imputation procedure: outcomes, exposures, and covariates (refer to main text for the full set of potential confounders)
- Treatment of variables: predictive mean matching for numeric data, and logistic regression imputation for binary data (factor with 2 levels); environmental chemical exposure variables were log-transformed
- No statistical interactions were included in the imputation models

**Table S1.** Selected characteristics of the study subset for whom fecal samples had been collected from the children and thus Shannon diversity or SCFA data were available (n=438[a]).

| | ***n* (%) or median (IQR)** | **Missing** |
|---|---|---|
| **Maternal characteristics** | | |
| Age at start of pregnancy, years | | 0 |
| 16–26 | 83 (18.9) | |
| 27–33 | 250 (57.1) | |
| 34–44 | 105 (24.0) | |
| Education, years completed | | 34 |
| <12 | 67 (16.6) | |
| 12 | 114 (28.2) | |
| 13-16 | 148 (36.6) | |
| ≥17 | 75 (18.6) | |
| Caesarean-section | 133 (30.4) | 0 |
| Smoking during pregnancy | 61 (14.1) | 5 |
| Smoking 1 year after delivery | 56 (14.5) | 51 |
| **Child characteristics** | | |
| Year of birth | | 0 |
| 2002–2003 | 157 (35.8) | |
| 2004–2005 | 281 (64.2) | |
| Sex: girl | 209 (47.7) | 0 |
| Preterm (<37 weeks gestation) | 99 (22.6) | 0 |
| Breastfeeding duration, any, months | 10.0 (4.3–13.0) | 12 |
| Breastfeeding, any, for at least 12 months | 181 (42.5) | 12 |
| Recent antibiotics, before age 12 months[b] | 24 (6.3) | 21 |
| Ever antibiotics, before age 24 months | 139 (39.2) | 83 |
| Asthma, diagnosed 2008–2014[c] | 18 (4.1) | 0 |
| Asthma, maternal-reported at 2 years of age | 35 (10.1) | 90 |
| LRTI ≥ 1 before 2 years of age | 81 (24.6) | 109 |

BMI, body mass index; IQR, interquartile range; LRTIs, lower respiratory tract infections (maternal-reported doctor diagnosis of bronchitis, respiratory syncytial virus, or pneumonia).

[a] This is the sample size for singletons with fecal samples analyzed for Shannon diversity and/or SCFAs for the samples collected at 4, 12 or 24 months.

[b] Antibiotics taken in the 2 weeks (or, if unavailable, in 1 month) before to when the age 12 month sample was collected.

[c] The median age of children in the sample of 438 at the time of registry linkage was 10.4 years old (IQR, 9.8–11.1; range, 9.3–11.7).

**Table S2.** Detection frequencies and concentrations of POP exposure biomarkers[a] in breast milk samples (n=993; HUMIS cohort, Norway, 2002–2009).

| Exposure | N[b] | % > LOD | Median LOD[c] | 5 P | 25 P | 50 P | 75 P | 95 P |
|---|---|---|---|---|---|---|---|---|
| PCB-74 (ng/g) | 993 | 100.0 | 0.005 | 1.651 | 2.400 | 3.250 | 4.360 | 7.004 |
| PCB-99 (ng/g) | 993 | 100.0 | 0.005 | 2.078 | 3.210 | 4.230 | 5.680 | 8.382 |
| PCB-105 (ng/g) | 993 | 100.0 | 0.002 | 0.638 | 0.980 | 1.350 | 1.852 | 3.221 |
| PCB-114 (ng/g) | 993 | 98.5 | 0.001 | 0.157 | 0.249 | 0.335 | 0.460 | 0.727 |
| PCB-118 (ng/g) | 993 | 100.0 | 0.001 | 3.047 | 4.540 | 6.170 | 8.400 | 14.202 |
| PCB-138 (ng/g) | 993 | 100.0 | 0.003 | 9.408 | 14.887 | 19.869 | 26.140 | 40.460 |
| PCB-153 (ng/g) | 993 | 100.0 | 0.005 | 16.660 | 24.900 | 32.180 | 42.820 | 68.606 |
| PCB-156 (ng/g) | 993 | 100.0 | 0.002 | 1.506 | 2.328 | 3.210 | 4.595 | 7.611 |
| PCB-157 (ng/g) | 993 | 99.8 | 0.002 | 0.288 | 0.471 | 0.670 | 1.020 | 1.794 |
| PCB-167 (ng/g) | 969 | 100.0 | 0.001 | 0.380 | 0.585 | 0.786 | 1.070 | 1.794 |
| PCB-170 (ng/g) | 993 | 99.7 | 0.002 | 1.570 | 4.560 | 6.192 | 8.430 | 13.741 |
| PCB-180 (ng/g) | 993 | 100.0 | 0.003 | 7.843 | 11.870 | 15.542 | 20.990 | 34.618 |
| PCB-189 (ng/g) | 993 | 98.8 | 0.001 | 0.100 | 0.162 | 0.230 | 0.320 | 0.552 |
| PCB-194 (ng/g) | 993 | 99.8 | 0.002 | 0.610 | 0.950 | 1.360 | 1.940 | 3.172 |
| $\sum_7$DL-PCBs (pmol/g) | — | — | — | 19.467 | 28.491 | 38.059 | 51.070 | 86.540 |
| $\sum_7$NDL-PCBs (pmol/g) | — | — | — | 119.789 | 176.911 | 228.843 | 301.537 | 473.785 |
| $\sum_{14}$PCBs (pmol/g) | — | — | — | 142.185 | 209.678 | 266.751 | 350.733 | 550.665 |
| HCB (ng/g) | 993 | 100.0 | 0.02 | 5.998 | 8.687 | 10.683 | 13.090 | 18.993 |
| β-HCH (ng/g) | 993 | 99.9 | 0.05 | 1.361 | 2.670 | 3.960 | 5.810 | 10.842 |
| oxychlordane (ng/g) | 882 | 100.0 | 0.05 | 1.417 | 2.262 | 3.018 | 4.140 | 7.295 |
| DDE (ng/g) | 993 | 100.0 | 0.05 | 18.282 | 32.048 | 46.406 | 71.270 | 152.787 |
| $\sum_4$OCPs (pmol/g) | — | — | — | 95.127 | 151.185 | 210.015 | 296.043 | 589.453 |
| BDE-28 (ng/g) | 955 | 97.5 | 0.02[d] | 0.031 | 0.076 | 0.123 | 0.214 | 0.493 |
| BDE-47 (ng/g) | 956 | 100.0 | 0.02[d] | 0.368 | 0.682 | 1.051 | 1.737 | 4.967 |
| BDE-99 (ng/g) | 956 | 99.9 | 0.02[d] | 0.105 | 0.182 | 0.278 | 0.457 | 1.359 |
| BDE-100 (ng/g) | 956 | 99.7 | 0.02[d] | 0.104 | 0.182 | 0.260 | 0.407 | 0.940 |
| BDE-153 (ng/g) | 953 | 99.8 | 0.02[d] | 0.234 | 0.365 | 0.501 | 0.700 | 1.390 |
| BDE-154 (ng/g) | 954 | 82.3 | 0.02[d] | <LOQ | 0.019 | 0.030 | 0.046 | 0.104 |
| $\sum_6$PBDEs (pmol/g) | — | — | — | 1.898 | 3.172 | 4.450 | 6.667 | 17.347 |
| PFOA (ng/L) | 884 | 89.7 | 10[d] | 9.957 | 25.000 | 40.000 | 63.000 | 110.000 |
| PFOS (ng/L) | 884 | 100.0 | 10[d] | 41.271 | 76.000 | 110.000 | 150.395 | 259.538 |
| $\sum_2$PFASs (pmol/L) | — | — | — | 132.507 | 227.043 | 322.788 | 454.105 | 740.038 |

β-HCH, β-hexachlorocyclohexane; DDE, dichlorodiphenyldichloroethylene; HCB, hexachlorobenzene; LOD, limit of detection; (N)DL, (non)-dioxin-like; OCP, organochlorine pesticide; P, percentile; (P)BDE, (poly)brominated diphenyl ether; PCB, polychlorinated biphenyl; PFAS, poly-and perfluoroalkyl substances; PFOA, perfluorooctanoate; PFOS, perfluorooctane sulfonate; POP, persistent organic pollutant.

[a] We present data for only those chemicals which were measured and detected in ≥ 70% (n ≥ 696) of the breast milk samples from the 993 mothers included in the present analysis. This excluded, among others, PCB-209 (61%) and

DDT (61%), and 3 perfluoroalkyl acids (49%) which were measured in a smaller number of samples. Values <LOD/LOD were imputed prior to calculating percentiles.

[b] The number of mothers whose breast milk samples were analyzed.

[c] Sample-specific LOD/LOQs were determined for PCBs and OCPs. The signal/noise (S/N) ratio of the calibration solutions was used to estimate the LOD (S/N = 3) and LOQ (S/N = 10).

[d] A single limit of quantification (LOQ) (and not an LOD) was reported by the laboratory for these analytes.

**Table S3.** Detection frequencies and levels[a] of SCFAs and Shannon diversity of microbiota from the fecal samples collected from the children at 4, 12, and 24 months of age.

| | n[b] | % <LOD | Absolute (mmol/kg) | | |
|---|---|---|---|---|---|
| | | | 25 P | 50 P | 75 P |
| **4 months** | | | | | |
| Acetic acid | 140 | 0.0 | 56.66 | 87.47 | 128.33 |
| Propionic acid | 140 | 1.4 | 4.42 | 10.22 | 19.99 |
| *n*-butyric acid | 140 | 2.1 | 0.74 | 2.68 | 6.22 |
| *i*-butyric acid | 140 | 12.9 | 0.33 | 0.94 | 1.66 |
| *n*-valeric acid | 140 | 52.1 | <LOD | <LOD | 0.17 |
| *i*-valeric acid | 140 | 12.9 | 0.24 | 1.10 | 2.10 |
| Total SCFAs[c] | 140 | 0.0 | 70.69 | 110.60 | 151.37 |
| Shannon diversity[d] | 390 | 0.0 | 2.69 | 3.11 | 3.51 |
| **12 months** | | | | | |
| Acetic acid | 324 | 0.0 | 55.71 | 75.24 | 106.85 |
| Propionic acid | 324 | 0.0 | 9.39 | 15.20 | 23.75 |
| *n*-butyric acid | 324 | 0.0 | 8.06 | 13.29 | 20.20 |
| *i*-butyric acid | 324 | 2.2 | 0.76 | 1.51 | 2.43 |
| *n*-valeric acid | 324 | 20.7 | 0.07 | 0.25 | 0.67 |
| *i*-valeric acid | 324 | 1.2 | 0.88 | 1.89 | 3.29 |
| Total SCFAs | 324 | 0.0 | 83.93 | 112.83 | 157.50 |
| Shannon diversity | 301 | 0.0 | 3.87 | 4.37 | 4.79 |
| **24 months** | | | | | |
| Acetic acid | 253 | 0.0 | 54.74 | 73.23 | 91.02 |
| Propionic acid | 253 | 0.0 | 11.65 | 17.58 | 25.05 |
| *n*-butyric acid | 253 | 0.0 | 11.91 | 18.07 | 26.13 |
| *i*-butyric acid | 253 | 0.8 | 1.14 | 1.88 | 2.84 |
| *n*-valeric acid | 253 | 3.6 | 0.50 | 1.60 | 2.58 |
| *i*-valeric acid | 253 | 0.0 | 1.34 | 2.43 | 3.83 |
| Total SCFAs | 253 | 0.0 | 88.86 | 115.96 | 147.91 |
| Shannon diversity | 127 | 0.0 | 4.41 | 4.94 | 5.29 |

[a] The absolute concentrations (mmol/kg) of SCFAs. 74–84% of the *n*-caproic acid and *i*-caproic acid values were <LOD (not shown in the table).

[b] The number of samples that were analyzed (refer to the Supplemental Methods) at the last 3 collection time points.

[c] Calculated with values <LOD imputed.

[d] Shannon diversity index is unitless, on a scale of 0–1.

**Table S4.** Adjusted associations [*β* or odds ratio (OR)][a] between covariates and the respiratory health outcomes, selected gut microbiota markers, and selected POP exposures.

| Covariate (2SD increment) | Ever asthma diagnosis<br>OR | Asthma at 2 years<br>OR | LRTIs ≥1 by 2 years<br>OR | ∑SCFAs (135 mmol/kg)[b]<br>*β* | Shannon diversity (1.44)[b]<br>*β* | ln-$\sum_{14}$PCBs (0.83 pmol/g)<br>*β* | ln-$\sum_4$OCPs (1.13 pmol/g)<br>*β* | ln-$\sum_6$PBDEs (1.39 pmol/g)<br>*β* | ln-$\sum_2$PFASs (1.08 pmol/L)<br>*β* |
|---|---|---|---|---|---|---|---|---|---|
| Age at start of pregnancy (9.3 years) | 1.72 | 0.80 | 0.99 | 0.071 | -0.053 | **0.506** | **0.397** | 0.065 | **0.100** |
| Mat. education, years completed: >12 | 1.21 | 1.22 | 1.16 | 0.003 | -0.114 | 0.021 | -0.006 | -0.072 | -0.057 |
| Pre-pregnancy BMI (8.8 kg/m$^2$) | **0.40** | 0.84 | 1.20 | -0.074 | 0.032 | **-0.087** | **0.064** | 0.066 | -0.052 |
| Parity: multiparous | 1.37 | 1.26 | 1.04 | 0.103 | 0.048 | **-0.308** | **-0.312** | **-0.075** | **-0.329** |
| Caesarean-section | 1.20 | 0.72 | 1.26 | 0.141 | -0.007 | 0.020 | 0.076 | 0.053 | 0.047 |
| Smoking during pregnancy | 1.31 | 1.29 | 1.31 | -0.094 | -0.133 | 0.025 | -0.009 | 0.034 | -0.016 |
| Mat. fatty fish (>23 servings/year) | 0.52 | **0.49** | 1.04 | -0.047 | -0.002 | **0.056** | 0.044 | 0.047 | 0.048 |
| Child age in 2015 (2.79 years) | 1.16 | 1.22 | 1.06 | **0.270** | -0.059 | **0.182** | **0.174** | **0.163** | **0.268** |
| Sex: male | 1.19 | **2.99** | **1.58** | -0.104 | -0.033 | -0.015 | 0.036 | 0.024 | 0.002 |
| Small for gestational age (<10$^{th}$ P) | 1.46 | 1.71 | 0.57 | -0.097 | -0.191 | 0.019 | -0.013 | -0.043 | 0.055 |
| Preterm (<37 weeks gestation) | 2.03 | **2.30** | 1.69 | -0.027 | 0.033 | -0.077 | -0.053 | 0.082 | -0.004 |
| Breastfeeding duration (10.7 months) | 0.89 | 0.94 | 0.98 | -0.070 | -0.126 | **-0.123** | **-0.102** | 0.000 | 0.017 |
| Rapid growth, 0–6 months | 1.75 | 1.74 | 0.96 | 0.048 | 0.074 | 0.006 | 0.019 | -0.074 | 0.069 |
| Either parent has asthma | **3.91** | 1.73 | 1.07 | 0.009 | 0.106 | -0.019 | -0.016 | 0.031 | 0.032 |
| No. of playmates at 2 years: >4 | 0.78 | 0.84 | 1.29 | -0.016 | 0.037 | -0.001 | -0.004 | 0.033 | **0.087** |
| Pets during infancy | 1.41 | 1.19 | 0.97 | 0.046 | 0.009 | 0.007 | -0.058 | 0.000 | 0.015 |
| Antibiotics <1 year old | **3.14** | **2.54** | **1.94** | -0.016 | **-0.198** | **-0.110** | -0.049 | 0.031 | -0.029 |
| Antibiotics <4 weeks (12 mo. sample) | – | – | – | -0.071 | -0.275 | – | – | – | – |

Abbreviations: BMI, body mass index; LRTIs, lower respiratory tract infections; OCPs, organochlorine pesticides; PBDEs, polybrominated diphenyl ethers; PCBs, polychlorinated biphenyls; PFASs, poly-and perfluoroalkyl substances; POP, persistent organic pollutant; SCFA, short-chain fatty acids; SD, standard deviation.

[a] Coefficients from multivariable models are presented (the number of playmates and pets, and subsequently, antibiotics, were added separately to models as they resulted in a smaller sample size). The coefficient is bolded if the 95% confidence interval excluded zero. Continuous exposures and covariates were scaled to 2-standard deviations (these increments are presented in brackets). Sample sizes ranged from n=139–208 for SCFAs and diversity models; and n=592–927 for the asthma and LRTI outcomes and POP exposures. The maximum variance inflation factor ranged from 1.31–1.53 across models.

[b] ∑SCFAs and Shannon diversity index for child fecal samples sampled at 1 year of age.

**Table S5.** Associations between POP exposures and registry-based asthma by 2014 (median 10 years of age): single-pollutant unpenalized logistic regression models and multi-pollutant elastic net penalized logistic regression models.

| | | Minimally adjusted[a] | | | | | | | | | | | | Further adjusted[a] | | | | | | | | | | | |
|---|---|---|---|---|---|---|---|---|---|---|---|---|---|---|---|---|---|---|---|---|---|---|---|---|---|
| | | Unpenalized logistic regression | | | | | | | | | | ENET[b] | | Unpenalized logistic regression | | | | | | | | | | ENET[b] | |
| | | Complete case | | | | | Multiple imputation (n=993) | | | | | (n=993) | | Complete case | | | | | Multiple imputation (n=993) | | | | | (n=993) | |
| **Exposure** | Incr.[c] | n | OR | 95% CI | | *p*-val | OR | 95% CI | | *p*-val | *q*-val[d] | % | OR[d] | n | OR | 95% CI | | *p*-val | OR | 95% CI | | *p*-val | *q*-val[d] | % | OR |
| PCB-74 (ng/g) | 2.502 | 953 | 0.67 | 0.34 | 1.34 | 0.256 | 0.67 | 0.34 | 1.33 | 0.253 | 0.73 | 0 | 1 | 718 | 0.61 | 0.25 | 1.49 | 0.277 | 0.66 | 0.32 | 1.33 | 0.239 | 0.62 | 0 | 1 |
| PCB-99 (ng/g) | 2.423 | 953 | 0.67 | 0.36 | 1.26 | 0.215 | 0.68 | 0.36 | 1.26 | 0.220 | 0.72 | 0 | 1 | 718 | 0.60 | 0.27 | 1.34 | 0.211 | 0.63 | 0.32 | 1.21 | 0.164 | 0.58 | 0 | 1 |
| PCB-105 (ng/g) | 2.688 | 953 | 0.57 | 0.30 | 1.11 | 0.100 | 0.59 | 0.30 | 1.14 | 0.113 | 0.43 | 0 | 1 | 718 | 0.56 | 0.24 | 1.28 | 0.170 | 0.62 | 0.31 | 1.24 | 0.177 | 0.58 | 0 | 1 |
| PCB-114 (ng/g) | 5.520 | 953 | 1.09 | 0.56 | 2.13 | 0.803 | 1.11 | 0.57 | 2.16 | 0.765 | 0.92 | 6 | 1 | 718 | 0.79 | 0.26 | 2.39 | 0.679 | 1.19 | 0.57 | 2.46 | 0.642 | 1.00 | 7 | 1 |
| PCB-118 (ng/g) | 2.522 | 953 | 0.54 | 0.27 | 1.06 | 0.075 | 0.55 | 0.28 | 1.08 | 0.081 | 0.43 | 89 | **0.91** | 718 | 0.54 | 0.23 | 1.27 | 0.158 | 0.57 | 0.28 | 1.15 | 0.117 | 0.58 | 5 | 1 |
| PCB-138 (ng/g) | 2.518 | 953 | **0.54** | **0.29** | **0.98** | **0.043** | 0.55 | 0.30 | 1.00 | 0.050 | 0.43 | 91 | **0.67** | 718 | **0.36** | **0.16** | **0.77** | **0.009** | **0.47** | **0.25** | **0.91** | **0.026** | 0.56 | 80 | **0.69** |
| PCB-153 (ng/g) | 2.357 | 953 | 0.57 | 0.30 | 1.10 | 0.094 | 0.59 | 0.31 | 1.11 | 0.103 | 0.43 | 4 | 1 | 718 | **0.38** | **0.15** | **0.93** | **0.034** | 0.53 | 0.27 | 1.06 | 0.073 | 0.58 | 0 | 1 |
| PCB-156 (ng/g) | 2.749 | 953 | 0.73 | 0.37 | 1.45 | 0.369 | 0.75 | 0.38 | 1.46 | 0.396 | 0.92 | 0 | 1 | 718 | 0.64 | 0.25 | 1.61 | 0.342 | 0.78 | 0.38 | 1.59 | 0.492 | 0.95 | 0 | 1 |
| PCB-157 (ng/g) | 3.498 | 953 | 0.88 | 0.50 | 1.55 | 0.648 | 0.89 | 0.50 | 1.58 | 0.699 | 0.92 | 0 | 1 | 718 | 0.80 | 0.39 | 1.67 | 0.558 | 0.89 | 0.45 | 1.75 | 0.737 | 1.00 | 0 | 1 |
| PCB-167 (ng/g) | 2.608 | 929 | 0.61 | 0.30 | 1.22 | 0.162 | 0.57 | 0.29 | 1.15 | 0.115 | 0.43 | 1 | 1 | 698 | 0.49 | 0.19 | 1.30 | 0.154 | 0.57 | 0.27 | 1.20 | 0.139 | 0.58 | 0 | 1 |
| PCB-170 (ng/g) | 5.271 | 953 | 0.94 | 0.52 | 1.69 | 0.824 | 0.96 | 0.53 | 1.73 | 0.880 | 0.92 | 0 | 1 | 718 | 0.79 | 0.34 | 1.81 | 0.574 | 0.76 | 0.37 | 1.52 | 0.431 | 0.95 | 0 | 1 |
| PCB-180 (ng/g) | 2.523 | 953 | 0.54 | 0.26 | 1.12 | 0.097 | 0.58 | 0.30 | 1.14 | 0.115 | 0.43 | 8 | 1 | 718 | **0.33** | **0.12** | **0.92** | **0.034** | 0.57 | 0.29 | 1.15 | 0.116 | 0.58 | 0 | 1 |
| PCB-189 (ng/g) | 4.851 | 953 | 1.06 | 0.53 | 2.13 | 0.866 | 1.05 | 0.53 | 2.11 | 0.886 | 0.92 | 9 | 1 | 718 | 0.82 | 0.37 | 1.81 | 0.617 | 0.98 | 0.44 | 2.17 | 0.964 | 1.00 | 2 | 1 |
| PCB-194 (ng/g) | 3.313 | 953 | 0.81 | 0.45 | 1.43 | 0.460 | 0.82 | 0.46 | 1.45 | 0.490 | 0.92 | 0 | 1 | 718 | 0.70 | 0.37 | 1.31 | 0.260 | 0.79 | 0.41 | 1.50 | 0.466 | 0.95 | 0 | 1 |
| HCB (ng/g) | 2.093 | 953 | 1.27 | 0.67 | 2.40 | 0.459 | 1.26 | 0.68 | 2.35 | 0.464 | 0.92 | 12 | 1 | 718 | 1.31 | 0.55 | 3.11 | 0.542 | 1.25 | 0.64 | 2.42 | 0.511 | 0.95 | 12 | 1 |
| β-HCH (ng/g) | 3.886 | 953 | **1.91** | **1.06** | **3.44** | **0.032** | **1.91** | **1.06** | **3.42** | **0.031** | 0.43 | 91 | **1.65** | 718 | **2.27** | **1.02** | **5.04** | **0.045** | **1.95** | **1.02** | **3.72** | **0.043** | 0.56 | 80 | **1.45** |
| oxychlordane (ng/g) | 2.668 | 844 | 0.68 | 0.34 | 1.36 | 0.275 | 1.04 | 0.46 | 2.35 | 0.934 | 0.93 | 17 | 1 | 633 | 0.56 | 0.22 | 1.43 | 0.228 | 1.00 | 0.43 | 2.33 | 0.992 | 1.00 | 6 | 1 |
| DDE (ng/g) | 3.783 | 953 | 1.06 | 0.57 | 1.98 | 0.847 | 1.06 | 0.57 | 1.97 | 0.863 | 0.92 | 1 | 1 | 718 | 0.67 | 0.29 | 1.56 | 0.359 | 1.02 | 0.51 | 2.01 | 0.965 | 1.00 | 1 | 1 |
| BDE-28 (ng/g) | 5.834 | 916 | 1.20 | 0.66 | 2.18 | 0.553 | 1.15 | 0.61 | 2.15 | 0.665 | 0.92 | 9 | 1 | 690 | 1.27 | 0.58 | 2.74 | 0.551 | 1.08 | 0.54 | 2.17 | 0.826 | 1.00 | 2 | 1 |
| BDE-47 (ng/g) | 5.083 | 917 | 1.11 | 0.61 | 2.02 | 0.740 | 1.05 | 0.57 | 1.92 | 0.888 | 0.92 | 0 | 1 | 690 | 0.91 | 0.42 | 1.97 | 0.803 | 1.01 | 0.53 | 1.94 | 0.971 | 1.00 | 0 | 1 |
| BDE-99 (ng/g) | 5.079 | 917 | 1.27 | 0.70 | 2.28 | 0.432 | 1.18 | 0.64 | 2.17 | 0.591 | 0.92 | 4 | 1 | 690 | 0.99 | 0.46 | 2.12 | 0.969 | 1.14 | 0.59 | 2.22 | 0.695 | 1.00 | 4 | 1 |
| BDE-100 (ng/g) | 4.241 | 917 | 1.00 | 0.54 | 1.84 | 0.997 | 0.94 | 0.50 | 1.79 | 0.858 | 0.92 | 1 | 1 | 690 | 0.74 | 0.33 | 1.67 | 0.468 | 0.88 | 0.44 | 1.76 | 0.711 | 1.00 | 1 | 1 |
| BDE-153 (ng/g) | 3.172 | 914 | 1.13 | 0.61 | 2.09 | 0.705 | 1.13 | 0.58 | 2.19 | 0.720 | 0.92 | 22 | 1 | 688 | 0.84 | 0.40 | 1.74 | 0.631 | 1.08 | 0.54 | 2.17 | 0.827 | 1.00 | 9 | 1 |
| BDE-154 (ng/g) | 4.366 | 915 | 1.12 | 0.61 | 2.04 | 0.714 | 1.05 | 0.56 | 1.97 | 0.869 | 0.92 | 0 | 1 | 689 | 0.90 | 0.41 | 1.95 | 0.781 | 1.04 | 0.54 | 2.02 | 0.902 | 1.00 | 0 | 1 |
| PFOA (ng/L) | 4.101 | 846 | 1.25 | 0.59 | 2.65 | 0.559 | 1.15 | 0.51 | 2.60 | 0.729 | 0.92 | 19 | 1 | 634 | 1.42 | 0.53 | 3.80 | 0.485 | 1.00 | 0.43 | 2.31 | 0.997 | 1.00 | 13 | 1 |
| PFOS (ng/L) | 2.981 | 846 | 0.76 | 0.38 | 1.51 | 0.431 | 0.76 | 0.39 | 1.48 | 0.416 | 0.92 | 19 | 1 | 634 | 0.65 | 0.25 | 1.70 | 0.381 | 0.63 | 0.31 | 1.30 | 0.212 | 0.61 | 41 | 1 |
| $\sum_7$DL-PCBs (pmol/g) | 2.442 | 929 | 0.62 | 0.31 | 1.23 | 0.170 | 0.60 | 0.30 | 1.20 | 0.148 | | | | 698 | 0.56 | 0.23 | 1.41 | 0.219 | 0.62 | 0.30 | 1.29 | 0.200 | | | |
| $\sum_7$NDL-PCBs (pmol/g) | 2.319 | 953 | 0.57 | 0.29 | 1.12 | 0.101 | 0.59 | 0.31 | 1.13 | 0.112 | | | | 718 | **0.38** | **0.15** | **0.95** | **0.039** | 0.54 | 0.27 | 1.08 | 0.081 | | | |
| $\sum_{14}$PCBs (pmol/g) | 2.297 | 929 | 0.60 | 0.30 | 1.19 | 0.146 | 0.59 | 0.30 | 1.15 | 0.119 | | | | 698 | 0.40 | 0.15 | 1.05 | 0.064 | 0.55 | 0.27 | 1.12 | 0.097 | | | |
| $\sum_4$OCPs (pmol/g) | 3.109 | 844 | 1.32 | 0.69 | 2.53 | 0.409 | 1.40 | 0.75 | 2.63 | 0.290 | | | | 633 | 0.82 | 0.33 | 2.08 | 0.678 | 1.40 | 0.70 | 2.78 | 0.342 | | | |
| $\sum_6$PBDEs (pmol/g) | 4.015 | 913 | 1.14 | 0.63 | 2.06 | 0.666 | 1.06 | 0.58 | 1.94 | 0.845 | | | | 688 | 0.92 | 0.42 | 2.00 | 0.837 | 1.03 | 0.54 | 1.98 | 0.927 | | | |
| $\sum_2$PFASs (pmol/L) | 2.813 | 846 | 0.93 | 0.45 | 1.91 | 0.844 | 0.93 | 0.46 | 1.90 | 0.844 | | | | 634 | 0.92 | 0.35 | 2.39 | 0.859 | 0.78 | 0.37 | 1.64 | 0.506 | | | |

Abbreviations: (P)BDE, (poly)brominated diphenyl ether; BMI, body mass index; β-HCH, β-hexachlorocyclohexane; DDE, dichlorodiphenyldichloroethylene; ENET, elastic net; HCB, hexachlorobenzene; Incr., increment; OCP, organochlorine pesticides; PCB, polychlorinated biphenyl; PFAS, poly-and perfluoroalkyl substances; PFOA, perfluorooctanoate; PFOS, perfluorooctane sulfonate.

Odds ratios per a 2-standard deviation increase in natural-log (ln)–transformed exposure concentration.

[a] Models were adjusted for as described in Figure S4A.

[b] For elastic net modelling, we used 10-fold cross validation for model optimization (to determine $\lambda$ with $\alpha$=0.8), selecting the model yielding the minimum prediction error. We then averaged coefficients from the elastic net models for those exposures which were selected (OR≠1) in >50% of the 100 imputed datasets.

[c] Exposures were ln-transformed, mean-centered and rescaled to 2 times their standard deviations (increment presented on the ln-scale).

[d] $q$-values for the single-pollutant models were determined: corrected for multiple comparisons with a false discovery rate controlled at <5% (Benjamini and Hochberg 1995).

**Table S6.** Associations between POP exposures and <u>maternal-reported asthma at 2 years of age</u>: single-pollutant unpenalized logistic regression models and multi-pollutant elastic net penalized logistic regression models.

| | | **Minimally adjusted**[a] | | | | | | | | | | | | **Further adjusted**[a] | | | | | | | | | | | |
|---|---|---|---|---|---|---|---|---|---|---|---|---|---|---|---|---|---|---|---|---|---|---|---|---|---|
| | | **OLS logistic regression** | | | | | | | | | | **ENET**[b] | | **OLS logistic regression** | | | | | | | | | | **ENET**[b] | |
| | | Complete case | | | | | Multiple imputation (n=993) | | | | | (n=993) | | Complete case | | | | | Multiple imputation (n=993) | | | | | (n=993) | |
| **Exposure** | Incr.[c] | n | OR | 95% CI | | *p*-val | OR | 95% CI | | *p*-val | *q*-val[d] | % | OR | n | OR | 95% CI | | *p*-val | OR | 95% CI | | *p*-val | *q*-val[d] | % | OR |
| PCB-74 (ng/g) | 2.502 | 826 | 0.68 | 0.35 | 1.31 | 0.246 | 0.75 | 0.39 | 1.42 | 0.373 | 0.73 | 6 | 1 | 712 | 0.54 | 0.26 | 1.11 | 0.094 | 0.72 | 0.38 | 1.39 | 0.327 | 0.82 | 3 | 1 |
| PCB-99 (ng/g) | 2.423 | 826 | 0.66 | 0.36 | 1.20 | 0.170 | 0.73 | 0.41 | 1.32 | 0.301 | 0.73 | 8 | 1 | 712 | **0.45** | **0.23** | **0.87** | **0.018** | 0.68 | 0.37 | 1.27 | 0.226 | 0.82 | 1 | 1 |
| PCB-105 (ng/g) | 2.688 | 826 | **0.51** | **0.28** | **0.96** | **0.037** | 0.53 | 0.29 | 0.98 | **0.042** | 0.73 | 73 | **0.75** | 712 | **0.39** | **0.19** | **0.79** | **0.009** | 0.55 | 0.29 | 1.03 | 0.063 | 0.74 | 44 | 1 |
| PCB-114 (ng/g) | 5.520 | 826 | 0.72 | 0.43 | 1.21 | 0.213 | 0.79 | 0.49 | 1.28 | 0.338 | 0.73 | 33 | 1 | 712 | **0.45** | **0.24** | **0.86** | **0.015** | 0.76 | 0.44 | 1.33 | 0.335 | 0.82 | 21 | 1 |
| PCB-118 (ng/g) | 2.522 | 826 | 0.56 | 0.29 | 1.05 | 0.072 | 0.56 | 0.30 | 1.05 | 0.071 | 0.73 | 23 | 1 | 712 | **0.43** | **0.21** | **0.88** | **0.021** | 0.57 | 0.30 | 1.09 | 0.089 | 0.74 | 13 | 1 |
| PCB-138 (ng/g) | 2.518 | 826 | **0.55** | **0.31** | **0.97** | **0.039** | 0.66 | 0.39 | 1.13 | 0.131 | 0.73 | 56 | **0.84** | 712 | **0.38** | **0.20** | **0.73** | **0.004** | 0.60 | 0.34 | 1.07 | 0.085 | 0.74 | 28 | 1 |
| PCB-153 (ng/g) | 2.357 | 826 | 0.74 | 0.40 | 1.37 | 0.340 | 0.83 | 0.46 | 1.49 | 0.532 | 0.73 | 3 | 1 | 712 | 0.52 | 0.25 | 1.07 | 0.077 | 0.79 | 0.43 | 1.46 | 0.452 | 0.82 | 1 | 1 |
| PCB-156 (ng/g) | 2.749 | 826 | 0.68 | 0.36 | 1.30 | 0.240 | 0.75 | 0.41 | 1.37 | 0.355 | 0.73 | 0 | 1 | 712 | 0.61 | 0.29 | 1.29 | 0.198 | 0.77 | 0.40 | 1.46 | 0.416 | 0.82 | 0 | 1 |
| PCB-157 (ng/g) | 3.498 | 826 | 0.72 | 0.45 | 1.15 | 0.169 | 0.74 | 0.46 | 1.19 | 0.209 | 0.73 | 49 | 1 | 712 | 0.66 | 0.39 | 1.14 | 0.134 | 0.70 | 0.41 | 1.22 | 0.208 | 0.82 | 16 | 1 |
| PCB-167 (ng/g) | 2.608 | 804 | 0.61 | 0.32 | 1.17 | 0.136 | 0.61 | 0.33 | 1.12 | 0.107 | 0.73 | 31 | 1 | 692 | 0.47 | 0.22 | 1.01 | 0.054 | 0.59 | 0.31 | 1.13 | 0.114 | 0.74 | 12 | 1 |
| PCB-170 (ng/g) | 5.271 | 826 | 1.30 | 0.64 | 2.65 | 0.468 | 1.34 | 0.67 | 2.66 | 0.408 | 0.73 | 59 | **1.33** | 712 | 1.06 | 0.46 | 2.43 | 0.891 | 1.20 | 0.53 | 2.72 | 0.658 | 0.82 | 15 | 1 |
| PCB-180 (ng/g) | 2.523 | 826 | 0.66 | 0.33 | 1.30 | 0.226 | 0.81 | 0.44 | 1.50 | 0.506 | 0.73 | 10 | 1 | 712 | 0.52 | 0.24 | 1.15 | 0.108 | 0.80 | 0.42 | 1.53 | 0.504 | 0.82 | 3 | 1 |
| PCB-189 (ng/g) | 4.851 | 826 | 1.18 | 0.60 | 2.30 | 0.634 | 1.23 | 0.63 | 2.42 | 0.542 | 0.73 | 57 | **1.51** | 712 | 1.05 | 0.50 | 2.23 | 0.897 | 1.18 | 0.55 | 2.54 | 0.670 | 0.82 | 20 | 1 |
| PCB-194 (ng/g) | 3.313 | 826 | 0.80 | 0.48 | 1.33 | 0.381 | 0.87 | 0.51 | 1.49 | 0.618 | 0.77 | 5 | 1 | 712 | 0.76 | 0.41 | 1.40 | 0.382 | 0.86 | 0.46 | 1.59 | 0.623 | 0.82 | 1 | 1 |
| HCB (ng/g) | 2.093 | 826 | 0.65 | 0.35 | 1.21 | 0.176 | 0.74 | 0.41 | 1.35 | 0.322 | 0.73 | 27 | 1 | 712 | 0.56 | 0.27 | 1.15 | 0.116 | 0.69 | 0.36 | 1.33 | 0.269 | 0.82 | 11 | 1 |
| β-HCH (ng/g) | 3.886 | 826 | 1.31 | 0.73 | 2.36 | 0.367 | 1.27 | 0.72 | 2.24 | 0.401 | 0.73 | 63 | **1.35** | 712 | 1.35 | 0.65 | 2.81 | 0.421 | 1.24 | 0.65 | 2.37 | 0.513 | 0.82 | 27 | 1 |
| oxychlordane (ng/g) | 2.668 | 730 | 0.78 | 0.41 | 1.48 | 0.449 | 0.91 | 0.49 | 1.68 | 0.759 | 0.88 | 30 | 1 | 628 | 0.59 | 0.29 | 1.21 | 0.147 | 0.88 | 0.47 | 1.65 | 0.694 | 0.82 | 7 | 1 |
| DDE (ng/g) | 3.783 | 826 | 0.95 | 0.52 | 1.74 | 0.864 | 1.07 | 0.59 | 1.94 | 0.821 | 0.89 | 44 | 1 | 712 | 0.68 | 0.34 | 1.35 | 0.269 | 1.00 | 0.53 | 1.89 | 0.993 | 0.99 | 15 | 1 |
| BDE-28 (ng/g) | 5.834 | 795 | 1.57 | 0.92 | 2.67 | 0.098 | 1.30 | 0.75 | 2.25 | 0.348 | 0.73 | 48 | 1 | 684 | 1.23 | 0.66 | 2.30 | 0.510 | 1.26 | 0.70 | 2.29 | 0.442 | 0.82 | 20 | 1 |
| BDE-47 (ng/g) | 5.083 | 796 | 1.38 | 0.83 | 2.30 | 0.218 | 1.20 | 0.72 | 2.00 | 0.479 | 0.73 | 32 | 1 | 684 | 1.09 | 0.60 | 1.97 | 0.788 | 1.17 | 0.68 | 2.00 | 0.579 | 0.82 | 8 | 1 |
| BDE-99 (ng/g) | 5.079 | 796 | 1.33 | 0.79 | 2.22 | 0.284 | 1.21 | 0.71 | 2.05 | 0.484 | 0.73 | 43 | 1 | 684 | 1.10 | 0.61 | 1.98 | 0.764 | 1.16 | 0.66 | 2.05 | 0.600 | 0.82 | 13 | 1 |
| BDE-100 (ng/g) | 4.241 | 796 | 1.04 | 0.61 | 1.79 | 0.882 | 0.99 | 0.57 | 1.72 | 0.973 | 0.97 | 18 | 1 | 684 | 0.85 | 0.45 | 1.60 | 0.609 | 0.97 | 0.55 | 1.73 | 0.927 | 0.96 | 6 | 1 |
| BDE-153 (ng/g) | 3.172 | 793 | 0.99 | 0.58 | 1.72 | 0.983 | 1.08 | 0.63 | 1.85 | 0.778 | 0.88 | 24 | 1 | 682 | 0.91 | 0.52 | 1.59 | 0.741 | 1.09 | 0.63 | 1.89 | 0.759 | 0.86 | 10 | 1 |
| BDE-154 (ng/g) | 4.366 | 794 | 0.74 | 0.42 | 1.30 | 0.292 | 0.81 | 0.46 | 1.42 | 0.465 | 0.73 | 57 | **0.79** | 683 | 0.63 | 0.34 | 1.19 | 0.156 | 0.83 | 0.48 | 1.46 | 0.526 | 0.82 | 22 | 1 |
| PFOA (ng/L) | 4.101 | 733 | 1.30 | 0.66 | 2.60 | 0.451 | 1.05 | 0.54 | 2.05 | 0.887 | 0.92 | 33 | 1 | 628 | 1.28 | 0.61 | 2.72 | 0.515 | 0.96 | 0.49 | 1.90 | 0.912 | 0.96 | 12 | 1 |
| PFOS (ng/L) | 2.981 | 733 | 0.94 | 0.49 | 1.80 | 0.849 | 0.83 | 0.45 | 1.55 | 0.564 | 0.73 | 35 | 1 | 628 | 0.69 | 0.32 | 1.46 | 0.326 | 0.69 | 0.35 | 1.35 | 0.277 | 0.82 | 27 | 1 |
| $\sum_7$DL-PCBs (pmol/g) | 2.442 | 804 | 0.57 | 0.30 | 1.10 | 0.093 | 0.58 | 0.31 | 1.09 | 0.090 | | | | 692 | **0.45** | **0.21** | **0.95** | **0.037** | 0.59 | 0.31 | 1.14 | 0.114 | | | |
| $\sum_7$NDL-PCBs (pmol/g) | 2.319 | 826 | 0.67 | 0.36 | 1.27 | 0.220 | 0.79 | 0.44 | 1.42 | 0.423 | | | | 712 | **0.47** | **0.23** | **0.99** | **0.046** | 0.74 | 0.40 | 1.39 | 0.350 | | | |
| $\sum_{14}$PCBs (pmol/g) | 2.297 | 804 | 0.68 | 0.36 | 1.28 | 0.230 | 0.75 | 0.41 | 1.36 | 0.335 | | | | 692 | 0.48 | 0.23 | 1.02 | 0.056 | 0.71 | 0.38 | 1.34 | 0.291 | | | |
| $\sum_4$OCPs (pmol/g) | 3.109 | 730 | 1.03 | 0.53 | 1.99 | 0.933 | 1.09 | 0.57 | 2.08 | 0.788 | | | | 628 | 0.66 | 0.31 | 1.45 | 0.302 | 1.01 | 0.50 | 2.06 | 0.970 | | | |
| $\sum_6$PBDEs (pmol/g) | 4.015 | 792 | 1.34 | 0.81 | 2.22 | 0.249 | 1.19 | 0.71 | 1.99 | 0.514 | | | | 682 | 1.08 | 0.60 | 1.95 | 0.798 | 1.17 | 0.68 | 2.02 | 0.582 | | | |
| $\sum_2$PFASs (pmol/L) | 2.813 | 733 | 1.09 | 0.56 | 2.15 | 0.794 | 0.95 | 0.50 | 1.80 | 0.873 | | | | 628 | 0.91 | 0.43 | 1.92 | 0.800 | 0.81 | 0.42 | 1.58 | 0.536 | | | |

Refer to Table S5 footnotes.

**Table S7.** Associations between POP exposures and one or more <u>lower respiratory tract infections by 2 years of age</u>: single-pollutant unpenalized logistic regression models and multi-pollutant elastic net penalized logistic regression models.

| | | Minimally adjusted[a] | | | | | | | | | | | | Further adjusted[a] | | | | | | | | | | | |
|---|---|---|---|---|---|---|---|---|---|---|---|---|---|---|---|---|---|---|---|---|---|---|---|---|---|
| | | OLS logistic regression | | | | | | | | | | ENET[b] | | OLS logistic regression | | | | | | | | | | ENET[b] | |
| | | Complete case | | | | | Multiple imputation (n=993) | | | | | (n=993) | | Complete case | | | | | Multiple imputation (n=993) | | | | | (n=993) | |
| **Exposure** | Incr.[c] | n | OR | 95% CI | | *p*-val | OR | 95% CI | | *p*-val | *q*-val[d] | % | OR | n | OR | 95% CI | | *p*-val | OR | 95% CI | | *p*-val | *q*-val[d] | % | OR |
| PCB-74 (ng/g) | 2.502 | 755 | 1.14 | 0.75 | 1.75 | 0.542 | 1.09 | 0.73 | 1.64 | 0.674 | 0.94 | 2 | 1 | 661 | 1.13 | 0.71 | 1.77 | 0.612 | 1.05 | 0.70 | 1.59 | 0.816 | 0.93 | 6 | 1 |
| PCB-99 (ng/g) | 2.423 | 755 | 0.98 | 0.66 | 1.45 | 0.902 | 1.00 | 0.69 | 1.46 | 0.995 | 1.00 | 4 | 1 | 661 | 0.96 | 0.63 | 1.47 | 0.864 | 0.95 | 0.65 | 1.40 | 0.805 | 0.93 | 7 | 1 |
| PCB-105 (ng/g) | 2.688 | 755 | 1.40 | 0.94 | 2.09 | 0.096 | 1.34 | 0.92 | 1.97 | 0.132 | 0.94 | 36 | 1 | 661 | 1.29 | 0.84 | 1.98 | 0.238 | 1.32 | 0.89 | 1.95 | 0.168 | 0.93 | 24 | 1 |
| PCB-114 (ng/g) | 5.520 | 755 | 1.24 | 0.74 | 2.09 | 0.420 | 1.15 | 0.72 | 1.82 | 0.567 | 0.94 | 15 | 1 | 661 | 1.07 | 0.60 | 1.91 | 0.813 | 1.11 | 0.68 | 1.82 | 0.666 | 0.93 | 17 | 1 |
| PCB-118 (ng/g) | 2.522 | 755 | 1.44 | 0.96 | 2.15 | 0.076 | 1.34 | 0.91 | 1.98 | 0.133 | 0.94 | 27 | 1 | 661 | 1.34 | 0.88 | 2.07 | 0.177 | 1.32 | 0.89 | 1.96 | 0.162 | 0.93 | 22 | 1 |
| PCB-138 (ng/g) | 2.518 | 755 | 0.90 | 0.60 | 1.34 | 0.602 | 0.90 | 0.62 | 1.32 | 0.595 | 0.94 | 9 | 1 | 661 | 0.86 | 0.55 | 1.33 | 0.487 | 0.86 | 0.58 | 1.27 | 0.442 | 0.93 | 11 | 1 |
| PCB-153 (ng/g) | 2.357 | 755 | 0.96 | 0.64 | 1.46 | 0.862 | 0.96 | 0.65 | 1.42 | 0.846 | 0.94 | 0 | 1 | 661 | 0.94 | 0.60 | 1.47 | 0.771 | 0.92 | 0.61 | 1.39 | 0.694 | 0.93 | 2 | 1 |
| PCB-156 (ng/g) | 2.749 | 755 | 0.93 | 0.61 | 1.41 | 0.719 | 0.95 | 0.64 | 1.41 | 0.799 | 0.94 | 0 | 1 | 661 | 0.90 | 0.57 | 1.43 | 0.661 | 0.93 | 0.62 | 1.41 | 0.747 | 0.93 | 1 | 1 |
| PCB-157 (ng/g) | 3.498 | 755 | 0.80 | 0.56 | 1.13 | 0.204 | 0.84 | 0.60 | 1.18 | 0.310 | 0.94 | 29 | 1 | 661 | 0.78 | 0.53 | 1.13 | 0.189 | 0.82 | 0.57 | 1.17 | 0.270 | 0.93 | 18 | 1 |
| PCB-167 (ng/g) | 2.608 | 739 | 1.27 | 0.84 | 1.92 | 0.262 | 1.20 | 0.81 | 1.77 | 0.366 | 0.94 | 7 | 1 | 646 | 1.17 | 0.74 | 1.85 | 0.492 | 1.17 | 0.78 | 1.76 | 0.448 | 0.93 | 12 | 1 |
| PCB-170 (ng/g) | 5.271 | 755 | 0.94 | 0.67 | 1.32 | 0.732 | 0.94 | 0.68 | 1.29 | 0.691 | 0.94 | 2 | 1 | 661 | 0.87 | 0.58 | 1.31 | 0.513 | 0.84 | 0.58 | 1.22 | 0.364 | 0.93 | 6 | 1 |
| PCB-180 (ng/g) | 2.523 | 755 | 0.77 | 0.49 | 1.21 | 0.259 | 0.82 | 0.54 | 1.24 | 0.340 | 0.94 | 16 | 1 | 661 | 0.78 | 0.47 | 1.28 | 0.318 | 0.80 | 0.52 | 1.22 | 0.294 | 0.93 | 13 | 1 |
| PCB-189 (ng/g) | 4.851 | 755 | 0.81 | 0.59 | 1.12 | 0.206 | 0.82 | 0.60 | 1.13 | 0.226 | 0.94 | 25 | 1 | 661 | 0.86 | 0.60 | 1.24 | 0.417 | 0.80 | 0.57 | 1.11 | 0.182 | 0.93 | 19 | 1 |
| PCB-194 (ng/g) | 3.313 | 755 | 0.74 | 0.51 | 1.06 | 0.101 | 0.74 | 0.52 | 1.06 | 0.104 | 0.94 | 47 | 1 | 661 | 0.72 | 0.49 | 1.06 | 0.093 | 0.71 | 0.49 | 1.02 | 0.067 | 0.93 | 31 | 1 |
| HCB (ng/g) | 2.093 | 755 | 1.11 | 0.73 | 1.68 | 0.621 | 1.21 | 0.82 | 1.81 | 0.337 | 0.94 | 24 | 1 | 661 | 1.10 | 0.70 | 1.73 | 0.670 | 1.21 | 0.80 | 1.84 | 0.364 | 0.93 | 20 | 1 |
| β-HCH (ng/g) | 3.886 | 755 | 1.15 | 0.77 | 1.73 | 0.486 | 1.21 | 0.83 | 1.76 | 0.321 | 0.94 | 22 | 1 | 661 | 1.07 | 0.66 | 1.72 | 0.798 | 1.17 | 0.77 | 1.76 | 0.468 | 0.93 | 15 | 1 |
| oxychlordane (ng/g) | 2.668 | 665 | 1.11 | 0.72 | 1.72 | 0.624 | 0.95 | 0.62 | 1.45 | 0.812 | 0.94 | 14 | 1 | 580 | 1.03 | 0.65 | 1.64 | 0.905 | 0.95 | 0.62 | 1.45 | 0.794 | 0.93 | 13 | 1 |
| DDE (ng/g) | 3.783 | 755 | 1.02 | 0.69 | 1.50 | 0.926 | 1.05 | 0.72 | 1.53 | 0.801 | 0.94 | 4 | 1 | 661 | 0.99 | 0.65 | 1.51 | 0.966 | 0.98 | 0.67 | 1.45 | 0.934 | 0.94 | 10 | 1 |
| BDE-28 (ng/g) | 5.834 | 731 | 0.95 | 0.66 | 1.37 | 0.791 | 0.99 | 0.68 | 1.44 | 0.963 | 1.00 | 6 | 1 | 640 | 0.95 | 0.63 | 1.42 | 0.790 | 0.93 | 0.63 | 1.38 | 0.733 | 0.93 | 11 | 1 |
| BDE-47 (ng/g) | 5.083 | 732 | 0.93 | 0.65 | 1.34 | 0.694 | 0.94 | 0.66 | 1.35 | 0.749 | 0.94 | 9 | 1 | 640 | 1.00 | 0.68 | 1.48 | 0.995 | 0.90 | 0.62 | 1.31 | 0.567 | 0.93 | 12 | 1 |
| BDE-99 (ng/g) | 5.079 | 732 | 1.09 | 0.77 | 1.55 | 0.630 | 1.03 | 0.72 | 1.47 | 0.868 | 0.94 | 7 | 1 | 640 | 1.17 | 0.80 | 1.71 | 0.431 | 0.99 | 0.68 | 1.43 | 0.940 | 0.94 | 9 | 1 |
| BDE-100 (ng/g) | 4.241 | 732 | 1.00 | 0.70 | 1.43 | 0.978 | 0.96 | 0.68 | 1.37 | 0.821 | 0.94 | 2 | 1 | 640 | 1.11 | 0.75 | 1.63 | 0.614 | 0.93 | 0.64 | 1.33 | 0.678 | 0.93 | 4 | 1 |
| BDE-153 (ng/g) | 3.172 | 729 | 0.98 | 0.68 | 1.41 | 0.915 | 0.97 | 0.66 | 1.41 | 0.857 | 0.94 | 7 | 1 | 638 | 1.04 | 0.71 | 1.52 | 0.856 | 0.97 | 0.66 | 1.41 | 0.856 | 0.93 | 9 | 1 |
| BDE-154 (ng/g) | 4.366 | 730 | 1.02 | 0.72 | 1.46 | 0.899 | 0.97 | 0.68 | 1.39 | 0.872 | 0.94 | 5 | 1 | 639 | 1.13 | 0.77 | 1.65 | 0.538 | 0.96 | 0.67 | 1.38 | 0.834 | 0.93 | 9 | 1 |
| PFOA (ng/L) | 4.101 | 672 | 1.10 | 0.72 | 1.67 | 0.676 | 1.06 | 0.69 | 1.61 | 0.803 | 0.94 | 11 | 1 | 587 | 1.13 | 0.72 | 1.80 | 0.592 | 1.04 | 0.68 | 1.60 | 0.860 | 0.93 | 12 | 1 |
| PFOS (ng/L) | 2.981 | 672 | 0.97 | 0.64 | 1.48 | 0.900 | 0.93 | 0.63 | 1.37 | 0.722 | 0.94 | 10 | 1 | 587 | 0.82 | 0.51 | 1.33 | 0.427 | 0.84 | 0.55 | 1.28 | 0.413 | 0.93 | 21 | 1 |
| $\sum_7$DL-PCBs (pmol/g) | 2.442 | 739 | 1.23 | 0.82 | 1.86 | 0.316 | 1.21 | 0.82 | 1.78 | 0.345 | | | | 646 | 1.15 | 0.74 | 1.79 | 0.537 | 1.19 | 0.80 | 1.78 | 0.398 | | | |
| $\sum_7$NDL-PCBs (pmol/g) | 2.319 | 755 | 0.90 | 0.60 | 1.37 | 0.632 | 0.92 | 0.62 | 1.36 | 0.659 | | | | 661 | 0.87 | 0.55 | 1.38 | 0.561 | 0.87 | 0.58 | 1.31 | 0.508 | | | |
| $\sum_{14}$PCBs (pmol/g) | 2.297 | 739 | 0.93 | 0.61 | 1.42 | 0.722 | 0.96 | 0.65 | 1.43 | 0.848 | | | | 646 | 0.87 | 0.54 | 1.39 | 0.556 | 0.92 | 0.61 | 1.39 | 0.693 | | | |
| $\sum_4$OCPs (pmol/g) | 3.109 | 665 | 1.05 | 0.68 | 1.60 | 0.840 | 1.05 | 0.71 | 1.55 | 0.802 | | | | 580 | 0.98 | 0.62 | 1.56 | 0.943 | 1.00 | 0.67 | 1.50 | 1.000 | | | |
| $\sum_6$PBDEs (pmol/g) | 4.015 | 728 | 0.96 | 0.67 | 1.37 | 0.805 | 0.94 | 0.66 | 1.34 | 0.731 | | | | 638 | 1.03 | 0.70 | 1.52 | 0.871 | 0.90 | 0.62 | 1.30 | 0.572 | | | |
| $\sum_2$PFASs (pmol/L) | 2.813 | 672 | 1.06 | 0.69 | 1.62 | 0.802 | 1.00 | 0.67 | 1.49 | 0.997 | | | | 587 | 0.96 | 0.59 | 1.56 | 0.871 | 0.93 | 0.61 | 1.40 | 0.713 | | | |

Refer to Table S5 footnotes.

**Table S8.** Unpenalized multi-pollutant logistic regression models of (i) the elastic net selected subset, (ii) summed chemical groups, and (iii) assessment of potential effect modification by child sex of the associations between POP exposures and respiratory health outcomes.

| Outcome/ Selected exposures (2SD (ln ng/g)) | Overall | | | | | Stratified by child sex | | | | |
|---|---|---|---|---|---|---|---|---|---|---|
| | Complete case | | | Multiple imputation (n=993) | | Girl (n=375/359/339) | | Boy (n=468/436/415) | | |
| | N | OR | 95% CI | OR | 95% CI | OR | 95% CI | OR | 95% CI | $p_{\text{interaction}}$ |
| **Asthma by 2014** | | | | | | | | | | |
| *ENET-selected subset* | 953 | | | | | *Reduced subset (full selected subset VIFs >3)* | | | | |
| PCB-118 (DL) | | 0.64 | 0.27, 1.51 | 0.65 | 0.27, 1.55 | – | | – | | |
| PCB-138 (NDL) | | **0.42** | **0.19, 0.89** | **0.43** | **0.20, 0.91** | 0.42 | 0.15, 1.16 | **0.31** | **0.13, 0.74** | 0.34 |
| *β*-HCH | | **3.02** | **1.66, 5.50** | **2.99** | **1.65, 5.43** | **3.35** | **1.50, 7.51** | 2.34 | 0.90, 6.05 | 0.47 |
| | | VIFs: 1.39–1.85 | | | | | | | | |
| *Summed groups* | 714 | | | | | | | | | |
| $\sum_{14}$PCBs | | **0.36** | **0.13, 0.99** | **0.39** | **0.18, 0.82** | | | | | |
| $\sum_{4}$OCPs | | **2.42** | **1.01, 5.78** | **2.05** | **1.06, 3.94** | | | | | |
| $\sum_{6}$PBDEs | | 0.99 | 0.44, 2.21 | 1.05 | 0.55, 2.02 | | | | | |
| $\sum_{2}$PFASs | | 1.05 | 0.44, 2.50 | 1.06 | 0.50, 2.25 | | | | | |
| | | VIFs:[a] 1.12–2.00 | | | | | | | | |
| **Asthma at 2 years** | | | | | | | | | | |
| *ENET-selected subset* | 794 | | | | | *Reduced subset (full selected subset VIFs >3)* | | | | |
| PCB-105 (DL) | | 0.49 | 0.21, 1.15 | 0.49 | 0.22, 1.10 | – | | – | | |
| PCB-138 (NDL) | | 0.39 | 0.13, 1.17 | **0.42** | **0.18, 0.97** | 1.62 | 0.33, 8.03 | **0.26** | **0.11, 0.59** | 0.05 |
| PCB-170 (NDL) | | 2.12 | 0.51, 8.78 | 2.03 | 0.54, 7.67 | – | | – | | |
| PCB-189 (DL) | | 1.98 | 0.51, 7.69 | 2.01 | 0.57, 7.05 | – | | | | |
| *β*-HCH | | **2.09** | **1.04, 4.22** | **1.96** | **1.03, 3.73** | 1.54 | 0.38, 6.27 | **2.41** | **1.05, 5.52** | 0.40 |
| BDE-154 | | 0.85 | 0.49, 1.50 | 0.88 | 0.51, 1.53 | – | | – | | |
| | | VIFs: 1.08–3.56 | | | | | | | | |
| *Summed groups* | 617 | | | | | | | | | |
| $\sum_{14}$PCBs | | 0.66 | 0.22, 2.01 | 0.59 | 0.28, 1.25 | | | | | |
| $\sum_{4}$OCPs | | 1.47 | 0.49, 4.43 | 1.35 | 0.64, 2.87 | | | | | |
| $\sum_{6}$PBDEs | | 1.48 | 0.80, 2.75 | 1.23 | 0.72, 2.08 | | | | | |
| $\sum_{2}$PFASs | | 1.06 | 0.49, 2.33 | 1.01 | 0.52, 1.94 | | | | | |
| | | VIFs: 1.09–2.87 | | | | | | | | |

| Outcome/ Selected exposures (2SD (ln ng/g)) | Overall | | | | | Stratified by child sex | | | | |
|---|---|---|---|---|---|---|---|---|---|---|
| | Complete case | | | Multiple imputation (n=993) | | Girl (n=375/359/339) | | Boy (n=468/436/415) | | |
| | N | OR | 95% CI | OR | 95% CI | OR | 95% CI | OR | 95% CI | $p_{interaction}$ |
| **LRTI by 2 years** | | | | | | | | | | |
| *Summed groups* | 564 | | | | | | | | | |
| $\sum_{14}$PCBs | | 1.26 | 0.60, 2.65 | 0.90 | 0.54, 1.51 | | | | | |
| $\sum_{4}$OCPs | | 0.81 | 0.38, 1.71 | 1.13 | 0.68, 1.87 | | | | | |
| $\sum_{6}$PBDEs | | 0.96 | 0.61, 1.51 | 0.93 | 0.64, 1.34 | | | | | |
| $\sum_{2}$PFASs | | 1.05 | 0.64, 1.74 | 1.02 | 0.68, 1.52 | | | | | |
| | | VIFs: 1.11–3.01 | | | | | | | | |

Abbreviations: β-HCH, β-hexachlorocyclohexane; DL, dioxin-like; ENET, elastic net regression; NDL, non-dioxin-like; (P)BDE, (poly)brominated diphenyl ether; OCP, organochlorine pesticides; PCB, polychlorinated biphenyl; PFAS, poly-and perfluoroalkyl substances; VIF, variance inflation factor. Odds ratios per a 2-standard deviation increase in natural-log (ln)–transformed exposure concentration. Models were adjusted for maternal age, education, pre-pregnancy BMI, and parity.

[a] Of the 4 summed chemical groups, VIFs were highest for the $\sum_{14}$PCBs and $\sum_{4}$OCPs.

**Table S9.** Associations between Shannon diversity and SCFAs and the three respiratory health outcomes: unpenalized logistic regression models.

| | | **Unadjusted** | | | | | **Minimally adjusted**[a] | | | | | | | | | **Further adjusted**[a] | | | | | | | | |
|---|---|---|---|---|---|---|---|---|---|---|---|---|---|---|---|---|---|---|---|---|---|---|---|---|
| | | Complete case | | | | | Complete case | | | | | Multiple imp. (n=438) | | | | Complete case | | | | | Multiple imp. (n=438) | | | |
| | Incr.[b] | n | OR | 95% CI | | *p*-val | n | OR | 95% CI | | *p*-val | OR | 95% CI | | *p*-val | n | OR | 95% CI | | *p*-val | OR | 95% CI | | *p*-val |
| **Asthma by 2014 (median 10.4 years of age), registry-based** | | | | | | | | | | | | | | | | | | | | | | | | |
| **4 months** | | | | | | | | | | | | | | | | | | | | | | | | |
| Diversity | 1.30 | 390 | **2.76** | **1.01** | **7.53** | **0.047** | 254 | **3.73** | **1.02** | **13.60** | **0.046** | 2.67 | 0.94 | 7.55 | 0.065 | 251 | **4.12** | **1.05** | **16.23** | **0.043** | 2.98 | 0.96 | 9.24 | 0.059 |
| ∑SCFAs | 145.89 | 140 | 0.34 | 0.04 | 2.66 | 0.302 | 86 | 0.07 | 0.00 | 2.07 | 0.124 | 0.63 | 0.14 | 2.90 | 0.550 | 86 | 0.01 | 0.00 | 3.99 | 0.133 | 0.63 | 0.13 | 3.15 | 0.571 |
| Acetic | 125.41 | 140 | 0.33 | 0.04 | 2.72 | 0.300 | 86 | 0.06 | 0.00 | 2.01 | 0.117 | 0.59 | 0.12 | 2.75 | 0.494 | 86 | 0.02 | 0.00 | 5.00 | 0.159 | 0.58 | 0.12 | 2.90 | 0.503 |
| Propionic | 29.00 | 140 | 0.66 | 0.10 | 4.27 | 0.662 | 86 | 0.23 | 0.01 | 5.09 | 0.350 | 0.92 | 0.24 | 3.52 | 0.906 | 86 | 0.11 | 0.00 | 4.61 | 0.247 | 1.00 | 0.24 | 4.21 | 0.994 |
| *n*-butyric | 10.93 | 140 | 0.39 | 0.04 | 3.80 | 0.414 | 86 | 0.45 | 0.02 | 12.85 | 0.638 | 0.62 | 0.11 | 3.38 | 0.579 | 86 | 0.23 | 0.00 | 36.04 | 0.566 | 0.63 | 0.11 | 3.72 | 0.607 |
| *i*-butyric | 2.11 | 140 | 1.23 | 0.29 | 5.20 | 0.775 | 86 | 0.23 | 0.01 | 4.40 | 0.326 | 1.49 | 0.47 | 4.71 | 0.498 | 86 | 0.14 | 0.00 | 4.54 | 0.266 | 1.59 | 0.44 | 5.73 | 0.480 |
| *n*-valeric | 0.77 | 140 | 0.34 | 0.01 | 13.25 | 0.567 | 86 | 0.96 | 0.01 | 66.44 | 0.984 | 0.62 | 0.07 | 5.93 | 0.676 | 86 | 1.01 | 0.01 | 169.29 | 0.997 | 0.58 | 0.06 | 6.07 | 0.644 |
| *i*-valeric | 3.32 | 140 | 1.08 | 0.25 | 4.72 | 0.921 | 86 | 0.30 | 0.02 | 5.89 | 0.428 | 1.40 | 0.45 | 4.36 | 0.562 | 86 | 0.15 | 0.00 | 8.67 | 0.359 | 1.57 | 0.43 | 5.72 | 0.490 |
| **12 months** | | | | | | | | | | | | | | | | | | | | | | | | |
| Diversity | 1.47 | 301 | 1.38 | 0.46 | 4.11 | 0.563 | 193 | 1.38 | 0.35 | 5.45 | 0.649 | 1.10 | 0.37 | 3.27 | 0.863 | 191 | 1.37 | 0.34 | 5.54 | 0.662 | 1.09 | 0.36 | 3.31 | 0.875 |
| ∑SCFAs | 129.78 | 324 | 1.08 | 0.38 | 3.04 | 0.883 | 201 | 1.64 | 0.56 | 4.87 | 0.369 | 1.07 | 0.39 | 2.95 | 0.895 | 197 | 1.71 | 0.54 | 5.39 | 0.359 | 1.06 | 0.38 | 2.97 | 0.915 |
| Acetic | 94.37 | 324 | 0.95 | 0.31 | 2.85 | 0.923 | 201 | 1.31 | 0.45 | 3.83 | 0.624 | 0.94 | 0.31 | 2.81 | 0.909 | 197 | 1.35 | 0.43 | 4.22 | 0.610 | 0.91 | 0.29 | 2.85 | 0.875 |
| Propionic | 28.21 | 324 | 0.98 | 0.33 | 2.90 | 0.977 | 201 | 1.58 | 0.39 | 6.47 | 0.522 | 1.01 | 0.35 | 2.91 | 0.987 | 197 | 1.66 | 0.40 | 6.81 | 0.485 | 1.07 | 0.38 | 3.06 | 0.896 |
| *n*-butyric | 26.37 | 324 | 1.31 | 0.50 | 3.39 | 0.585 | 201 | 1.92 | 0.67 | 5.49 | 0.224 | 1.22 | 0.47 | 3.17 | 0.679 | 197 | 1.82 | 0.65 | 5.11 | 0.255 | 1.16 | 0.45 | 2.97 | 0.755 |
| *i*-butyric | 2.84 | 324 | 1.94 | 0.81 | 4.65 | 0.139 | 201 | 2.51 | 0.90 | 6.98 | 0.078 | 2.07 | 0.91 | 4.73 | 0.085 | 197 | 2.56 | 0.92 | 7.12 | 0.071 | 2.01 | 0.88 | 4.57 | 0.098 |
| *n*-valeric | 2.76 | 324 | 1.39 | 0.63 | 3.05 | 0.417 | 201 | 1.80 | 0.67 | 4.84 | 0.248 | 1.35 | 0.63 | 2.91 | 0.444 | 197 | 1.89 | 0.66 | 5.42 | 0.234 | 1.35 | 0.61 | 2.96 | 0.458 |
| *i*-valeric | 4.19 | 324 | 2.18 | 0.94 | 5.05 | 0.069 | 201 | **2.91** | **1.09** | **7.78** | **0.034** | **2.29** | **1.04** | **5.05** | **0.041** | 197 | **3.02** | **1.12** | **8.14** | **0.029** | **2.25** | **1.02** | **4.98** | **0.045** |
| **24 months** | | | | | | | | | | | | | | | | | | | | | | | | |
| Diversity | 1.44 | 127 | 0.62 | 0.16 | 2.46 | 0.497 | 83 | 1.08 | 0.09 | 12.56 | 0.949 | 0.83 | 0.21 | 3.25 | 0.782 | 82 | 1.19 | 0.14 | 10.27 | 0.875 | 0.84 | 0.21 | 3.38 | 0.802 |
| ∑SCFAs | 120.20 | 253 | 0.71 | 0.17 | 2.98 | 0.635 | 168 | 0.96 | 0.16 | 5.76 | 0.968 | 0.77 | 0.22 | 2.66 | 0.674 | 164 | 0.93 | 0.15 | 5.74 | 0.937 | 0.80 | 0.22 | 2.90 | 0.734 |
| Acetic | 73.19 | 253 | 0.94 | 0.26 | 3.44 | 0.929 | 168 | 1.42 | 0.26 | 7.83 | 0.687 | 0.93 | 0.28 | 3.04 | 0.899 | 164 | 1.14 | 0.22 | 5.85 | 0.878 | 0.96 | 0.28 | 3.23 | 0.941 |
| Propionic | 25.18 | 253 | 0.46 | 0.09 | 2.43 | 0.359 | 168 | 0.55 | 0.08 | 3.58 | 0.530 | 0.59 | 0.15 | 2.37 | 0.455 | 164 | 0.64 | 0.09 | 4.71 | 0.664 | 0.61 | 0.15 | 2.51 | 0.493 |
| *n*-butyric | 31.50 | 253 | 0.62 | 0.13 | 3.02 | 0.551 | 168 | 0.70 | 0.10 | 5.22 | 0.730 | 0.68 | 0.17 | 2.67 | 0.577 | 164 | 0.68 | 0.08 | 5.95 | 0.724 | 0.75 | 0.18 | 3.12 | 0.691 |
| *i*-butyric | 2.95 | 253 | 0.48 | 0.10 | 2.31 | 0.363 | 168 | 0.81 | 0.15 | 4.47 | 0.813 | 0.73 | 0.19 | 2.81 | 0.650 | 164 | 0.96 | 0.18 | 5.15 | 0.965 | 0.70 | 0.19 | 2.63 | 0.593 |
| *n*-valeric | 3.19 | 253 | 0.46 | 0.09 | 2.30 | 0.341 | 168 | 0.41 | 0.06 | 2.85 | 0.366 | 0.49 | 0.11 | 2.19 | 0.346 | 164 | 0.54 | 0.07 | 4.22 | 0.558 | 0.50 | 0.11 | 2.25 | 0.360 |
| *i*-valeric | 4.38 | 253 | 0.46 | 0.09 | 2.32 | 0.346 | 168 | 0.78 | 0.14 | 4.32 | 0.775 | 0.65 | 0.16 | 2.69 | 0.552 | 164 | 0.97 | 0.19 | 4.99 | 0.970 | 0.63 | 0.15 | 2.58 | 0.518 |
| **Asthma at 2 years, maternal-reported** | | | | | | | | | | | | | | | | | | | | | | | | |
| **4 months** | | | | | | | | | | | | | | | | | | | | | | | | |
| Diversity | 1.30 | **308** | **2.30** | **1.02** | **5.19** | **0.045** | 215 | 2.64 | 0.96 | 7.26 | 0.060 | 1.96 | 0.91 | 4.22 | 0.086 | 212 | 2.84 | 0.99 | 8.19 | 0.053 | 2.30 | 0.99 | 5.33 | 0.052 |
| ∑SCFAs | 145.89 | 113 | 0.16 | 0.02 | 1.62 | 0.121 | 73 | 0.11 | 0.00 | 4.99 | 0.258 | 0.73 | 0.31 | 1.70 | 0.456 | 73 | 0.01 | 0.00 | 3.01 | 0.114 | 0.75 | 0.30 | 1.83 | 0.519 |
| Acetic | 125.41 | 113 | 0.15 | 0.01 | 1.65 | 0.121 | 73 | 0.07 | 0.00 | 4.77 | 0.212 | 0.74 | 0.31 | 1.75 | 0.487 | 73 | 0.01 | 0.00 | 5.08 | 0.148 | 0.74 | 0.30 | 1.84 | 0.518 |
| Propionic | 29.00 | 113 | 0.18 | 0.01 | 2.48 | 0.202 | 73 | 0.70 | 0.06 | 8.34 | 0.777 | 0.72 | 0.31 | 1.68 | 0.448 | 73 | 0.22 | 0.01 | 7.67 | 0.401 | 0.77 | 0.32 | 1.86 | 0.556 |
| *n*-butyric | 10.93 | 113 | 1.30 | 0.37 | 4.63 | 0.681 | 73 | 1.37 | 0.18 | 10.58 | 0.761 | 1.00 | 0.41 | 2.45 | 0.997 | 73 | 0.75 | 0.07 | 8.06 | 0.810 | 1.10 | 0.44 | 2.74 | 0.839 |
| *i*-butyric | 2.11 | 113 | 0.53 | 0.10 | 2.89 | 0.464 | 73 | 0.35 | 0.02 | 5.98 | 0.467 | 0.65 | 0.25 | 1.72 | 0.383 | 73 | 0.13 | 0.00 | 3.80 | 0.236 | 0.69 | 0.25 | 1.90 | 0.464 |
| *n*-valeric | 0.77 | 113 | 1.47 | 0.23 | 9.43 | 0.688 | 73 | 0.06 | 0.00 | 106.14 | 0.467 | 1.53 | 0.42 | 5.61 | 0.514 | 73 | 0.03 | 0.00 | 47.27 | 0.343 | 1.55 | 0.40 | 6.07 | 0.521 |
| *i*-valeric | 3.32 | 113 | 0.73 | 0.15 | 3.57 | 0.695 | 73 | 0.65 | 0.06 | 7.61 | 0.730 | 0.85 | 0.37 | 1.95 | 0.703 | 73 | 0.26 | 0.01 | 7.57 | 0.436 | 0.95 | 0.40 | 2.30 | 0.914 |
| **12 months** | | | | | | | | | | | | | | | | | | | | | | | | |

| | | Unadjusted | | | | | Minimally adjusted[a] | | | | | | | | | Further adjusted[a] | | | | | | | | |
|---|---|---|---|---|---|---|---|---|---|---|---|---|---|---|---|---|---|---|---|---|---|---|---|---|
| | | Complete case | | | | | Complete case | | | | | Multiple imp. (n=438) | | | | Complete case | | | | | Multiple imp. (n=438) | | | |
| | Incr.[b] | n | OR | 95% CI | | *p*-val | n | OR | 95% CI | | *p*-val | OR | 95% CI | | *p*-val | n | OR | 95% CI | | *p*-val | OR | 95% CI | | *p*-val |
| Diversity | 1.47 | 255 | 0.77 | 0.35 | 1.73 | 0.531 | 171 | 0.58 | 0.21 | 1.59 | 0.289 | 0.68 | 0.31 | 1.48 | 0.324 | 169 | 0.57 | 0.20 | 1.64 | 0.297 | 0.67 | 0.29 | 1.55 | 0.349 |
| ∑SCFAs | 129.78 | 280 | 1.44 | 0.62 | 3.33 | 0.400 | 182 | 1.19 | 0.34 | 4.19 | 0.790 | 1.53 | 0.76 | 3.06 | 0.229 | 178 | 1.44 | 0.38 | 5.47 | 0.595 | 1.54 | 0.75 | 3.18 | 0.238 |
| Acetic | 94.37 | 280 | 1.42 | 0.62 | 3.24 | 0.406 | 182 | 0.99 | 0.28 | 3.47 | 0.990 | 1.51 | 0.76 | 3.03 | 0.242 | 178 | 1.15 | 0.30 | 4.49 | 0.840 | 1.55 | 0.74 | 3.26 | 0.245 |
| Propionic | 28.21 | 280 | 1.06 | 0.45 | 2.52 | 0.896 | 182 | 1.13 | 0.31 | 4.18 | 0.856 | 1.18 | 0.56 | 2.49 | 0.655 | 178 | 1.26 | 0.34 | 4.63 | 0.731 | 1.22 | 0.57 | 2.62 | 0.609 |
| *n*-butyric | 26.37 | 280 | 1.21 | 0.54 | 2.71 | 0.653 | 182 | 1.47 | 0.51 | 4.26 | 0.481 | 1.28 | 0.64 | 2.55 | 0.487 | 178 | 1.52 | 0.55 | 4.23 | 0.418 | 1.22 | 0.61 | 2.45 | 0.565 |
| *i*-butyric | 2.84 | 280 | 1.81 | 0.84 | 3.92 | 0.130 | 182 | 1.45 | 0.51 | 4.11 | 0.481 | 1.59 | 0.83 | 3.04 | 0.164 | 178 | 1.54 | 0.56 | 4.25 | 0.408 | 1.57 | 0.81 | 3.04 | 0.178 |
| *n*-valeric | 2.76 | 280 | 1.04 | 0.40 | 2.68 | 0.939 | 182 | 1.53 | 0.59 | 3.95 | 0.383 | 1.10 | 0.50 | 2.40 | 0.812 | 178 | 1.61 | 0.58 | 4.41 | 0.359 | 1.06 | 0.47 | 2.40 | 0.888 |
| *i*-valeric | 4.19 | 280 | 1.86 | 0.89 | 3.88 | 0.101 | 182 | 1.43 | 0.51 | 3.98 | 0.493 | 1.47 | 0.76 | 2.85 | 0.253 | 178 | 1.55 | 0.57 | 4.20 | 0.391 | 1.48 | 0.76 | 2.89 | 0.252 |
| **24 months** | | | | | | | | | | | | | | | | | | | | | | | | |
| Diversity | 1.44 | 114 | 0.62 | 0.20 | 1.95 | 0.410 | 81 | 0.71 | 0.16 | 3.22 | 0.657 | 0.91 | 0.30 | 2.80 | 0.865 | 80 | 0.62 | 0.11 | 3.37 | 0.578 | 0.89 | 0.28 | 2.79 | 0.838 |
| ∑SCFAs | 120.20 | 237 | 0.55 | 0.19 | 1.60 | 0.269 | 162 | 1.40 | 0.44 | 4.42 | 0.566 | 0.81 | 0.35 | 1.88 | 0.615 | 158 | 1.57 | 0.48 | 5.10 | 0.454 | 0.87 | 0.36 | 2.10 | 0.758 |
| Acetic | 73.19 | 237 | 0.49 | 0.17 | 1.45 | 0.196 | 162 | 1.23 | 0.37 | 4.12 | 0.735 | 0.76 | 0.33 | 1.76 | 0.517 | 158 | 1.29 | 0.37 | 4.44 | 0.689 | 0.80 | 0.34 | 1.90 | 0.611 |
| Propionic | 25.18 | 237 | 0.71 | 0.27 | 1.87 | 0.484 | 162 | 1.42 | 0.50 | 4.00 | 0.510 | 0.90 | 0.39 | 2.06 | 0.801 | 158 | 1.63 | 0.55 | 4.84 | 0.383 | 0.96 | 0.40 | 2.26 | 0.917 |
| *n*-butyric | 31.50 | 237 | 0.88 | 0.34 | 2.24 | 0.785 | 162 | 1.56 | 0.53 | 4.61 | 0.420 | 1.00 | 0.46 | 2.18 | 0.996 | 158 | 2.01 | 0.61 | 6.67 | 0.255 | 1.17 | 0.53 | 2.62 | 0.695 |
| *i*-butyric | 2.95 | 237 | 0.51 | 0.18 | 1.43 | 0.199 | 162 | 0.97 | 0.28 | 3.37 | 0.966 | 0.80 | 0.32 | 1.97 | 0.622 | 158 | 1.03 | 0.30 | 3.51 | 0.965 | 0.76 | 0.31 | 1.87 | 0.551 |
| *n*-valeric | 3.19 | 237 | 0.54 | 0.19 | 1.52 | 0.246 | 162 | 0.98 | 0.31 | 3.07 | 0.976 | 0.71 | 0.29 | 1.74 | 0.451 | 158 | 1.18 | 0.37 | 3.72 | 0.778 | 0.71 | 0.29 | 1.77 | 0.463 |
| *i*-valeric | 4.38 | 237 | 0.51 | 0.18 | 1.47 | 0.210 | 162 | 0.94 | 0.27 | 3.27 | 0.922 | 0.77 | 0.32 | 1.88 | 0.564 | 158 | 1.04 | 0.30 | 3.54 | 0.956 | 0.74 | 0.31 | 1.80 | 0.509 |
| **LRTI by 2 years, maternal-reported** | | | | | | | | | | | | | | | | | | | | | | | | |
| **4 months** | | | | | | | | | | | | | | | | | | | | | | | | |
| Diversity | 1.30 | 292 | 1.38 | 0.79 | 2.42 | 0.263 | 205 | 1.16 | 0.56 | 2.39 | 0.687 | 1.45 | 0.84 | 2.50 | 0.181 | 203 | 1.19 | 0.57 | 2.48 | 0.648 | 1.49 | 0.85 | 2.62 | 0.166 |
| ∑SCFAs | 145.89 | 111 | 1.39 | 0.60 | 3.25 | 0.442 | 73 | 1.16 | 0.25 | 5.37 | 0.847 | 1.08 | 0.59 | 1.98 | 0.802 | 73 | 1.17 | 0.22 | 6.30 | 0.853 | 1.09 | 0.59 | 2.02 | 0.779 |
| Acetic | 125.41 | 111 | 1.52 | 0.66 | 3.52 | 0.329 | 73 | 1.20 | 0.26 | 5.53 | 0.819 | 1.16 | 0.62 | 2.14 | 0.645 | 73 | 1.20 | 0.23 | 6.17 | 0.830 | 1.17 | 0.63 | 2.19 | 0.623 |
| Propionic | 29.00 | 111 | 1.17 | 0.51 | 2.71 | 0.714 | 73 | 1.54 | 0.51 | 4.64 | 0.448 | 1.00 | 0.55 | 1.82 | 0.993 | 73 | 1.77 | 0.52 | 6.06 | 0.364 | 1.01 | 0.55 | 1.87 | 0.973 |
| *n*-butyric | 10.93 | 111 | 0.55 | 0.18 | 1.72 | 0.305 | 73 | 0.06 | 0.00 | 1.69 | 0.099 | 0.64 | 0.30 | 1.38 | 0.248 | 73 | 0.02 | 0.00 | 1.08 | 0.054 | 0.62 | 0.28 | 1.39 | 0.242 |
| *i*-butyric | 2.11 | 111 | 0.39 | 0.13 | 1.25 | 0.113 | 73 | 0.43 | 0.09 | 2.10 | 0.298 | 0.55 | 0.25 | 1.22 | 0.141 | 73 | 0.36 | 0.06 | 2.01 | 0.242 | 0.53 | 0.24 | 1.21 | 0.130 |
| *n*-valeric | 0.77 | 111 | **0.01** | **0.00** | **0.70** | **0.035** | 73 | 0.00 | 0.00 | 3.42 | 0.074 | 0.59 | 0.20 | 1.75 | 0.340 | 73 | 0.00 | 0.00 | 1.53 | 0.057 | 0.57 | 0.19 | 1.75 | 0.323 |
| *i*-valeric | 3.32 | 111 | 0.52 | 0.17 | 1.60 | 0.251 | 73 | 0.54 | 0.11 | 2.65 | 0.446 | 0.72 | 0.36 | 1.45 | 0.354 | 73 | 0.45 | 0.08 | 2.65 | 0.380 | 0.71 | 0.34 | 1.45 | 0.342 |
| **12 months** | | | | | | | | | | | | | | | | | | | | | | | | |
| Diversity | 1.47 | 244 | 0.62 | 0.35 | 1.09 | 0.097 | 163 | **0.38** | **0.17** | **0.81** | **0.012** | 0.64 | 0.36 | 1.15 | 0.138 | 162 | **0.37** | **0.17** | **0.80** | **0.011** | 0.63 | 0.35 | 1.15 | 0.131 |
| ∑SCFAs | 129.78 | 270 | 1.15 | 0.65 | 2.03 | 0.631 | 175 | 0.85 | 0.34 | 2.16 | 0.733 | 1.18 | 0.71 | 1.99 | 0.521 | 172 | 0.92 | 0.36 | 2.35 | 0.861 | 1.17 | 0.70 | 1.97 | 0.549 |
| Acetic | 94.37 | 270 | 0.98 | 0.55 | 1.77 | 0.954 | 175 | 0.64 | 0.24 | 1.69 | 0.365 | 1.02 | 0.60 | 1.74 | 0.934 | 172 | 0.68 | 0.25 | 1.80 | 0.433 | 1.01 | 0.59 | 1.72 | 0.985 |
| Propionic | 28.21 | 270 | 1.67 | 0.92 | 3.02 | 0.092 | 175 | 1.37 | 0.54 | 3.50 | 0.512 | 1.51 | 0.83 | 2.72 | 0.173 | 172 | 1.50 | 0.58 | 3.90 | 0.401 | 1.52 | 0.84 | 2.76 | 0.164 |
| *n*-butyric | 26.37 | 270 | 1.15 | 0.66 | 2.00 | 0.624 | 175 | 1.42 | 0.68 | 2.99 | 0.354 | 1.23 | 0.73 | 2.07 | 0.429 | 172 | 1.50 | 0.70 | 3.21 | 0.295 | 1.23 | 0.73 | 2.06 | 0.438 |
| *i*-butyric | 2.84 | 270 | 1.64 | 0.94 | 2.87 | 0.084 | 175 | 1.40 | 0.64 | 3.07 | 0.403 | 1.48 | 0.87 | 2.52 | 0.146 | 172 | 1.42 | 0.64 | 3.15 | 0.393 | 1.48 | 0.87 | 2.53 | 0.147 |
| *n*-valeric | 2.76 | 270 | 0.80 | 0.42 | 1.52 | 0.491 | 175 | 0.55 | 0.18 | 1.70 | 0.297 | 0.81 | 0.44 | 1.50 | 0.500 | 172 | 0.58 | 0.19 | 1.77 | 0.334 | 0.81 | 0.44 | 1.50 | 0.498 |
| *i*-valeric | 4.19 | 270 | 1.56 | 0.90 | 2.70 | 0.116 | 175 | 1.26 | 0.58 | 2.76 | 0.556 | 1.36 | 0.80 | 2.32 | 0.250 | 172 | 1.27 | 0.57 | 2.82 | 0.553 | 1.36 | 0.80 | 2.32 | 0.251 |
| **24 months** | | | | | | | | | | | | | | | | | | | | | | | | |
| Diversity | 1.44 | 110 | 0.72 | 0.30 | 1.70 | 0.449 | 76 | 0.75 | 0.27 | 2.14 | 0.593 | 0.97 | 0.42 | 2.27 | 0.949 | 75 | 0.67 | 0.23 | 1.99 | 0.474 | 0.98 | 0.41 | 2.37 | 0.970 |
| ∑SCFAs | 120.20 | 225 | 0.59 | 0.29 | 1.24 | 0.163 | 154 | 0.45 | 0.15 | 1.30 | 0.138 | 0.87 | 0.49 | 1.54 | 0.631 | 151 | 0.46 | 0.15 | 1.40 | 0.171 | 0.87 | 0.49 | 1.55 | 0.634 |
| Acetic | 73.19 | 225 | 0.81 | 0.42 | 1.58 | 0.535 | 154 | 0.70 | 0.27 | 1.84 | 0.470 | 0.99 | 0.56 | 1.75 | 0.972 | 151 | 0.77 | 0.28 | 2.11 | 0.610 | 0.99 | 0.56 | 1.76 | 0.973 |
| Propionic | 25.18 | 225 | **0.38** | **0.17** | **0.86** | **0.020** | 154 | **0.30** | **0.10** | **0.96** | **0.041** | 0.67 | 0.36 | 1.27 | 0.220 | 151 | **0.28** | **0.09** | **0.93** | **0.037** | 0.67 | 0.36 | 1.27 | 0.222 |
| *n*-butyric | 31.50 | 225 | 0.60 | 0.28 | 1.29 | 0.189 | 154 | 0.33 | 0.09 | 1.18 | 0.088 | 0.88 | 0.49 | 1.57 | 0.659 | 151 | 0.34 | 0.09 | 1.25 | 0.104 | 0.88 | 0.49 | 1.58 | 0.663 |

| | | **Unadjusted** | | | | | **Minimally adjusted**[a] | | | | | | | | | **Further adjusted**[a] | | | | | | | | |
|---|---|---|---|---|---|---|---|---|---|---|---|---|---|---|---|---|---|---|---|---|---|---|---|---|
| | | Complete case | | | | | Complete case | | | | | Multiple imp. (n=438) | | | | Complete case | | | | | Multiple imp. (n=438) | | | |
| | Incr.[b] | n | OR | 95% CI | | *p*-val | n | OR | 95% CI | | *p*-val | OR | 95% CI | | *p*-val | n | OR | 95% CI | | *p*-val | OR | 95% CI | | *p*-val |
| *i*-butyric | 2.95 | 225 | **0.35** | **0.15** | **0.78** | **0.011** | 154 | **0.30** | **0.10** | **0.92** | **0.036** | 0.60 | 0.29 | 1.24 | 0.165 | 151 | **0.24** | **0.07** | **0.82** | **0.022** | 0.60 | 0.29 | 1.24 | 0.164 |
| *n*-valeric | 3.19 | 225 | 0.61 | 0.29 | 1.25 | 0.174 | 154 | 0.46 | 0.16 | 1.27 | 0.133 | 0.91 | 0.49 | 1.68 | 0.761 | 151 | 0.41 | 0.14 | 1.20 | 0.104 | 0.92 | 0.50 | 1.71 | 0.789 |
| *i*-valeric | 4.38 | 225 | **0.43** | **0.19** | **0.94** | **0.033** | 154 | 0.40 | 0.14 | 1.18 | 0.097 | 0.75 | 0.38 | 1.48 | 0.405 | 151 | 0.32 | 0.10 | 1.04 | 0.058 | 0.75 | 0.38 | 1.48 | 0.405 |

Abbreviations: CI, confidence interval; LRTI, lower respiratory tract infection; OR, odds ratio; SCFAs, short-chain fatty acids.

Regression coefficients were estimated from unpenalized logistic regression models. Odds ratios per a 2-standard deviation increase in untransformed diversity or absolute SCFA levels (mmol/kg).

[a] Models were adjusted for as described in Figure S4B.

[b] Exposures were mean-centered and rescaled to 2 times their standard deviations (increment).

**Table S10.** Associations[a] between individual SCFAs and the three respiratory health outcomes: penalized logistic regression models (n=438).

| | 4 months | | | 12 months | | | 24 months | | |
|---|---|---|---|---|---|---|---|---|---|
| | % >0 | OR[b] (α=0.1) | OR >50% sel.[c] | % >0 | mean OR (α=0.1) | OR >50% sel.[b] | % >0 | mean OR (α=0.1) | OR >50% sel.[b] |
| **Asthma at ~ 10 years** | | | | | | | | | |
| Acetic | 40 | 0.85 | 1 | 3 | 0.99 | 1 | 7 | 1.04 | 1 |
| Propionic | 29 | 0.99 | 1 | 10 | 0.97 | 1 | 18 | 0.96 | 1 |
| *n*-butyric | 39 | 0.86 | 1 | 2 | 1.01 | 1 | 9 | 0.98 | 1 |
| *i*-butyric | 37 | 1.60 | 1 | 5 | 1.01 | 1 | 8 | 1.86 | 1 |
| *n*-valeric | 37 | 0.88 | 1 | 1 | 1.00 | 1 | 35 | 0.88 | 1 |
| *i*-valeric | 29 | 1.24 | 1 | 40 | 1.32 | 1 | 10 | 0.97 | 1 |
| **Asthma at 2 years** | | | | | | | | | |
| Acetic | 58 | 0.94 | 0.94 | 50 | 1.21 | 1.21 | 42 | 0.85 | 1 |
| Propionic | 52 | 0.92 | 0.92 | 21 | 0.96 | 1 | 31 | 1.11 | 1 |
| *n*-butyric | 58 | 1.12 | 1.12 | 20 | 1.01 | 1 | 32 | 1.17 | 1 |
| *i*-butyric | 62 | 0.69 | 0.69 | 48 | 1.46 | 1 | 30 | 1.70 | 1 |
| *n*-valeric | 79 | 1.73 | 1.73 | 26 | 1.02 | 1 | 47 | 0.89 | 1 |
| *i*-valeric | 43 | 1.66 | 1 | 17 | 1.00 | 1 | 33 | 1.01 | 1 |
| **LRTI by 2 years** | | | | | | | | | |
| Acetic | 68 | 1.34 | 1.34 | 51 | 0.91 | 0.91 | 69 | 1.54 | 1.54 |
| Propionic | 63 | 1.17 | 1.17 | 80 | 1.44 | 1.44 | 75 | 0.71 | 0.71 |
| *n*-butyric | 81 | 0.76 | 0.76 | 55 | 1.09 | 1.09 | 63 | 0.89 | 0.89 |
| *i*-butyric | 83 | 0.52 | 0.52 | 71 | 2.58 | 2.58 | 80 | 0.48 | 0.48 |
| *n*-valeric | 83 | 0.79 | 0.79 | 82 | 0.69 | 0.69 | 66 | 1.45 | 1.45 |
| *i*-valeric | 55 | 2.20 | 2.20 | 34 | 0.87 | 1 | 61 | 3.38 | 3.38 |

[a] Model were minimally adjusted (see Figure S4B).

[b] Averaged coefficients from the elastic net models for all SCFAs across all 100 imputed datasets (results presented in main text, Figure 2). We present these results under with the view that mutually adjusted effect estimates (versus sparse selection) for all SCFAs are informative.

[c] Averaged coefficients from the elastic net models for those SCFAs which were selected in >50% of the 100 imputed datasets. We used 10-fold cross validation for model optimization (to determine λ with α=0.8).

**Table S11.** Associations between summed groups of POP exposures and Shannon diversity and SCFAs: unpenalized single- and multi-pollutant linear regression models.

| Exposure | Unadjusted, single-pollutant | | | | | Adjusted,[a] single-pollutant | | | | | | | | | Adjusted,[a] multi-pollutant | | | | | | | | |
|---|---|---|---|---|---|---|---|---|---|---|---|---|---|---|---|---|---|---|---|---|---|---|---|
| | Complete case | | | | | Complete case | | | | | Multiple imputation[b] (n=298) | | | | Complete case[c] | | | | | Multiple imp.[b] (n=298) | | | |
| (ln-2SD) | n | *β* | 95% CI | | *p*-val | n | *β* | 95% CI | | *p*-val | *β* | 95% CI | | *p*-val | n | *β* | 95% CI | | *p*-val | *β* | 95% CI | | *p*-val |
| **Diversity** | | | | | | | | | | | | | | | | | | | | | | | |
| **4 months** | | | | | | | | | | | | | | | | | | | | | | | |
| $\sum_7$DL-PCBs | 271 | 0.022 | -0.095 | 0.138 | 0.717 | 226 | -0.023 | -0.155 | 0.108 | 0.728 | 0.002 | -0.116 | 0.120 | 0.975 | | | | | | | | | |
| $\sum_7$NDL-PCBs | 273 | 0.058 | -0.058 | 0.175 | 0.330 | 228 | 0.026 | -0.105 | 0.156 | 0.700 | 0.028 | -0.090 | 0.146 | 0.637 | | | | | | | | | |
| $\sum_{14}$PCBs | 271 | 0.051 | -0.065 | 0.168 | 0.390 | 226 | 0.016 | -0.116 | 0.147 | 0.816 | 0.023 | -0.095 | 0.141 | 0.705 | 177 | -0.071 | -0.280 | 0.138 | 0.508 | -0.007 | -0.176 | 0.162 | 0.936 |
| $\sum_4$OCPs | 261 | 0.083 | -0.036 | 0.201 | 0.172 | 217 | 0.044 | -0.089 | 0.177 | 0.518 | 0.038 | -0.083 | 0.160 | 0.533 | . | 0.077 | -0.143 | 0.297 | 0.494 | 0.038 | -0.141 | 0.217 | 0.677 |
| $\sum_6$PBDEs | 263 | 0.012 | -0.107 | 0.132 | 0.839 | 221 | 0.022 | -0.104 | 0.148 | 0.732 | 0.034 | -0.086 | 0.154 | 0.579 | . | 0.067 | -0.075 | 0.210 | 0.355 | 0.025 | -0.105 | 0.155 | 0.704 |
| $\sum_2$PFASs | 226 | 0.032 | -0.098 | 0.163 | 0.628 | 189 | 0.007 | -0.131 | 0.144 | 0.922 | 0.001 | -0.131 | 0.133 | 0.987 | . | 0.001 | -0.153 | 0.155 | 0.989 | -0.010 | -0.150 | 0.130 | 0.888 |
| **12 months** | | | | | | | | | | | | | | | | | | | | | | | |
| $\sum_7$DL-PCBs | 204 | -0.108 | -0.242 | 0.026 | 0.116 | 179 | -0.107 | -0.258 | 0.045 | 0.168 | -0.122 | -0.262 | 0.018 | 0.087 | . | . | . | . | . | . | . | . | . |
| $\sum_7$NDL-PCBs | 207 | **-0.141** | **-0.276** | **-0.006** | **0.042** | 182 | **-0.160** | **-0.308** | **-0.012** | **0.035** | -0.113 | -0.247 | 0.022 | 0.099 | . | . | . | . | . | . | . | . | . |
| $\sum_{14}$PCBs | 204 | -0.109 | -0.242 | 0.024 | 0.109 | 179 | -0.121 | -0.269 | 0.028 | 0.112 | -0.117 | -0.252 | 0.018 | 0.090 | 143 | -0.008 | -0.265 | 0.249 | 0.952 | -0.025 | -0.235 | 0.185 | 0.812 |
| $\sum_4$OCPs | 200 | **-0.166** | **-0.296** | **-0.035** | **0.014** | 175 | **-0.191** | **-0.333** | **-0.048** | **0.010** | **-0.149** | **-0.278** | **-0.019** | **0.024** | . | -0.124 | -0.381 | 0.132 | 0.343 | -0.125 | -0.335 | 0.085 | 0.241 |
| $\sum_6$PBDEs | 198 | -0.041 | -0.166 | 0.083 | 0.515 | 175 | -0.057 | -0.188 | 0.074 | 0.395 | -0.047 | -0.180 | 0.086 | 0.485 | . | -0.004 | -0.147 | 0.139 | 0.954 | 0.002 | -0.138 | 0.142 | 0.973 |
| $\sum_2$PFASs | 176 | -0.050 | -0.195 | 0.095 | 0.499 | 155 | -0.102 | -0.255 | 0.051 | 0.194 | -0.066 | -0.213 | 0.082 | 0.381 | . | -0.048 | -0.211 | 0.114 | 0.561 | -0.028 | -0.181 | 0.124 | 0.713 |
| **24 months** | | | | | | | | | | | | | | | | | | | | | | | |
| $\sum_7$DL-PCBs | 88 | **-0.309** | **-0.502** | **-0.116** | **0.002** | 85 | **-0.321** | **-0.521** | **-0.121** | **0.002** | -0.169 | -0.377 | 0.040 | 0.111 | . | . | . | . | . | . | . | . | . |
| $\sum_7$NDL-PCBs | 89 | **-0.320** | **-0.503** | **-0.138** | **0.001** | 86 | **-0.317** | **-0.503** | **-0.131** | **0.001** | **-0.210** | **-0.400** | **-0.021** | **0.030** | . | . | . | . | . | . | . | . | . |
| $\sum_{14}$PCBs | 88 | **-0.319** | **-0.502** | **-0.136** | **0.001** | 85 | **-0.316** | **-0.503** | **-0.130** | **0.001** | **-0.207** | **-0.400** | **-0.015** | **0.035** | 68 | **-0.545** | **-0.789** | **-0.301** | **0.000** | **-0.285** | **-0.530** | **-0.041** | **0.022** |
| $\sum_4$OCPs | 84 | -0.059 | -0.249 | 0.131 | 0.543 | 81 | -0.040 | -0.244 | 0.165 | 0.704 | -0.081 | -0.274 | 0.112 | 0.405 | . | **0.393** | **0.079** | **0.708** | **0.017** | 0.105 | -0.142 | 0.352 | 0.399 |
| $\sum_6$PBDEs | 87 | 0.084 | -0.101 | 0.269 | 0.377 | 84 | 0.086 | -0.104 | 0.276 | 0.376 | 0.000 | -0.168 | 0.169 | 0.997 | . | 0.036 | -0.179 | 0.250 | 0.746 | 0.035 | -0.142 | 0.212 | 0.697 |
| $\sum_2$PFASs | 77 | -0.139 | -0.354 | 0.076 | 0.209 | 74 | -0.161 | -0.382 | 0.059 | 0.157 | -0.050 | -0.233 | 0.133 | 0.590 | . | -0.137 | -0.346 | 0.071 | 0.202 | -0.010 | -0.201 | 0.181 | 0.917 |
| **Total SCFAs** | | | | | | | | | | | | | | | | | | | | | | | |
| **4 months** | | | | | | | | | | | | | | | | | | | | | | | |
| $\sum_7$DL-PCBs | 87 | 0.002 | -0.205 | 0.208 | 0.988 | 74 | -0.045 | -0.263 | 0.172 | 0.683 | 0.016 | -0.140 | 0.173 | 0.835 | . | . | . | . | . | . | . | . | . |
| $\sum_7$NDL-PCBs | 89 | 0.071 | -0.123 | 0.264 | 0.476 | 76 | 0.050 | -0.150 | 0.250 | 0.623 | 0.048 | -0.103 | 0.199 | 0.529 | . | . | . | . | . | . | . | . | . |
| $\sum_{14}$PCBs | 87 | 0.060 | -0.140 | 0.259 | 0.559 | 74 | 0.026 | -0.181 | 0.233 | 0.807 | 0.044 | -0.107 | 0.195 | 0.566 | 65 | 0.202 | -0.070 | 0.473 | 0.151 | 0.098 | -0.112 | 0.308 | 0.359 |
| $\sum_4$OCPs | 89 | -0.051 | -0.276 | 0.173 | 0.654 | 76 | -0.198 | -0.439 | 0.044 | 0.113 | -0.013 | -0.165 | 0.139 | 0.868 | . | **-0.452** | **-0.812** | **-0.092** | **0.017** | -0.086 | -0.306 | 0.134 | 0.439 |
| $\sum_6$PBDEs | 84 | -0.068 | -0.326 | 0.189 | 0.605 | 71 | -0.120 | -0.376 | 0.137 | 0.363 | 0.005 | -0.141 | 0.151 | 0.948 | . | 0.035 | -0.248 | 0.317 | 0.812 | 0.007 | -0.150 | 0.165 | 0.926 |
| $\sum_2$PFASs | 79 | 0.067 | -0.180 | 0.313 | 0.598 | 69 | 0.042 | -0.193 | 0.277 | 0.726 | 0.018 | -0.146 | 0.183 | 0.825 | . | 0.063 | -0.198 | 0.323 | 0.639 | 0.015 | -0.156 | 0.186 | 0.862 |
| **12 months** | | | | | | | | | | | | | | | | | | | | | | | |
| $\sum_7$DL-PCBs | 221 | 0.097 | -0.035 | 0.229 | 0.149 | 196 | 0.079 | -0.053 | 0.211 | 0.243 | 0.069 | -0.058 | 0.196 | 0.285 | . | . | . | . | . | . | . | . | . |
| $\sum_7$NDL-PCBs | 224 | 0.126 | -0.003 | 0.255 | 0.057 | 199 | 0.096 | -0.031 | 0.222 | 0.139 | 0.105 | -0.020 | 0.231 | 0.100 | . | . | . | . | . | . | . | . | . |
| $\sum_{14}$PCBs | 221 | **0.135** | **0.004** | **0.265** | **0.044** | 196 | 0.107 | -0.022 | 0.236 | 0.104 | 0.102 | -0.024 | 0.227 | 0.113 | 157 | **0.339** | **0.147** | **0.531** | **0.001** | 0.127 | -0.045 | 0.299 | 0.146 |
| $\sum_4$OCPs | 213 | 0.067 | -0.065 | 0.199 | 0.320 | 188 | -0.044 | -0.176 | 0.088 | 0.513 | 0.044 | -0.082 | 0.171 | 0.490 | . | **-0.274** | **-0.478** | **-0.069** | **0.010** | -0.056 | -0.234 | 0.122 | 0.539 |
| $\sum_6$PBDEs | 215 | 0.018 | -0.108 | 0.144 | 0.785 | 191 | -0.005 | -0.123 | 0.113 | 0.937 | 0.022 | -0.105 | 0.149 | 0.733 | . | 0.054 | -0.075 | 0.183 | 0.411 | 0.003 | -0.131 | 0.137 | 0.964 |
| $\sum_2$PFASs | 191 | 0.079 | -0.050 | 0.209 | 0.232 | 173 | 0.026 | -0.112 | 0.164 | 0.710 | 0.067 | -0.080 | 0.214 | 0.372 | . | -0.014 | -0.162 | 0.134 | 0.853 | 0.049 | -0.107 | 0.204 | 0.536 |
| **24 months** | | | | | | | | | | | | | | | | | | | | | | | |

| | Unadjusted, single-pollutant | | | | | Adjusted,[a] single-pollutant | | | | | | | | | Adjusted,[a] multi-pollutant | | | | | | | | |
|---|---|---|---|---|---|---|---|---|---|---|---|---|---|---|---|---|---|---|---|---|---|---|---|
| Exposure | Complete case | | | | | Complete case | | | | | Multiple imputation[b] (n=298) | | | | Complete case[c] | | | | | Multiple imp.[b] (n=298) | | | |
| (ln-2SD) | n | *β* | 95% CI | | *p*-val | n | *β* | 95% CI | | *p*-val | *β* | 95% CI | | *p*-val | n | *β* | 95% CI | | *p*-val | *β* | 95% CI | | *p*-val |
| $\sum_7$DL-PCBs | 184 | -0.099 | -0.243 | 0.045 | 0.179 | 176 | -0.128 | -0.276 | 0.020 | 0.092 | -0.079 | -0.214 | 0.056 | 0.251 | . | . | . | . | . | . | . | . | . |
| $\sum_7$NDL-PCBs | 187 | -0.084 | -0.227 | 0.059 | 0.250 | 179 | -0.109 | -0.254 | 0.037 | 0.146 | -0.062 | -0.196 | 0.072 | 0.364 | . | . | . | . | . | . | . | . | . |
| $\sum_{14}$PCBs | 184 | -0.089 | -0.231 | 0.053 | 0.220 | 176 | -0.117 | -0.262 | 0.028 | 0.115 | -0.065 | -0.200 | 0.069 | 0.338 | 134 | 0.070 | -0.153 | 0.293 | 0.540 | 0.040 | -0.154 | 0.233 | 0.688 |
| $\sum_4$OCPs | 173 | **-0.156** | **-0.296** | **-0.016** | **0.030** | 165 | **-0.176** | **-0.321** | **-0.031** | **0.018** | -0.115 | -0.250 | 0.021 | 0.096 | . | -0.116 | -0.363 | 0.132 | 0.360 | -0.110 | -0.312 | 0.091 | 0.281 |
| $\sum_6$PBDEs | 180 | -0.128 | -0.264 | 0.008 | 0.068 | 172 | -0.125 | -0.264 | 0.013 | 0.078 | -0.066 | -0.196 | 0.065 | 0.324 | . | -0.068 | -0.225 | 0.088 | 0.395 | -0.027 | -0.166 | 0.113 | 0.707 |
| $\sum_2$PFASs | 156 | -0.148 | -0.299 | 0.002 | 0.056 | 151 | -0.138 | -0.294 | 0.017 | 0.082 | -0.113 | -0.254 | 0.028 | 0.116 | . | -0.100 | -0.272 | 0.072 | 0.256 | -0.092 | -0.242 | 0.059 | 0.230 |
| **Acetic** | | | | | | | | | | | | | | | | | | | | | | | |
| **4 months** | | | | | | | | | | | | | | | | | | | | | | | |
| $\sum_7$DL-PCBs | 87 | -0.039 | -0.245 | 0.168 | 0.714 | 74 | -0.074 | -0.293 | 0.145 | 0.511 | -0.025 | -0.178 | 0.128 | 0.749 | . | . | . | . | . | . | . | . | . |
| $\sum_7$NDL-PCBs | 89 | 0.016 | -0.178 | 0.210 | 0.870 | 76 | 0.004 | -0.196 | 0.205 | 0.967 | 0.002 | -0.146 | 0.150 | 0.981 | . | . | . | . | . | . | . | . | . |
| $\sum_{14}$PCBs | 87 | 0.014 | -0.185 | 0.214 | 0.888 | 74 | -0.008 | -0.216 | 0.200 | 0.939 | -0.003 | -0.151 | 0.146 | 0.973 | 65 | 0.182 | -0.093 | 0.457 | 0.200 | 0.045 | -0.162 | 0.252 | 0.670 |
| $\sum_4$OCPs | 89 | -0.121 | -0.344 | 0.102 | 0.291 | 76 | **-0.260** | **-0.499** | **-0.022** | **0.036** | -0.038 | -0.185 | 0.110 | 0.615 | . | **-0.407** | **-0.772** | **-0.042** | **0.033** | -0.047 | -0.263 | 0.168 | 0.663 |
| $\sum_6$PBDEs | 84 | -0.199 | -0.453 | 0.055 | 0.128 | 71 | **-0.260** | **-0.511** | **-0.008** | **0.047** | -0.075 | -0.219 | 0.068 | 0.302 | . | -0.131 | -0.417 | 0.156 | 0.375 | -0.072 | -0.226 | 0.082 | 0.356 |
| $\sum_2$PFASs | 79 | 0.058 | -0.195 | 0.312 | 0.653 | 69 | 0.028 | -0.214 | 0.270 | 0.819 | -0.001 | -0.161 | 0.160 | 0.994 | . | 0.081 | -0.183 | 0.344 | 0.552 | 0.011 | -0.154 | 0.177 | 0.895 |
| **12 months** | | | | | | | | | | | | | | | | | | | | | | | |
| $\sum_7$DL-PCBs | 221 | 0.117 | -0.015 | 0.249 | 0.083 | 196 | 0.108 | -0.024 | 0.240 | 0.112 | 0.073 | -0.053 | 0.199 | 0.256 | . | . | . | . | . | . | . | . | . |
| $\sum_7$NDL-PCBs | 224 | **0.136** | **0.007** | **0.265** | **0.040** | 199 | 0.126 | -0.001 | 0.252 | 0.053 | 0.106 | -0.019 | 0.230 | 0.096 | . | . | . | . | . | . | . | . | . |
| $\sum_{14}$PCBs | 221 | **0.147** | **0.017** | **0.277** | **0.028** | 196 | **0.140** | **0.011** | **0.268** | **0.034** | 0.102 | -0.023 | 0.227 | 0.108 | 157 | **0.360** | **0.173** | **0.547** | **0.000** | 0.088 | -0.083 | 0.259 | 0.312 |
| $\sum_4$OCPs | 213 | 0.105 | -0.027 | 0.238 | 0.120 | 188 | -0.005 | -0.138 | 0.128 | 0.937 | 0.075 | -0.051 | 0.201 | 0.242 | . | **-0.235** | **-0.434** | **-0.035** | **0.022** | 0.003 | -0.174 | 0.181 | 0.969 |
| $\sum_6$PBDEs | 215 | 0.021 | -0.106 | 0.148 | 0.746 | 191 | 0.000 | -0.120 | 0.119 | 0.994 | 0.020 | -0.105 | 0.144 | 0.756 | . | 0.044 | -0.082 | 0.169 | 0.494 | -0.010 | -0.143 | 0.122 | 0.877 |
| $\sum_2$PFASs | 191 | 0.104 | -0.020 | 0.229 | 0.101 | 173 | 0.048 | -0.086 | 0.181 | 0.486 | 0.080 | -0.072 | 0.232 | 0.300 | . | -0.005 | -0.150 | 0.139 | 0.946 | 0.059 | -0.102 | 0.220 | 0.471 |
| **24 months** | | | | | | | | | | | | | | | | | | | | | | | |
| $\sum_7$DL-PCBs | 184 | -0.035 | -0.176 | 0.107 | 0.634 | 176 | -0.055 | -0.197 | 0.088 | 0.451 | -0.036 | -0.169 | 0.098 | 0.597 | . | . | . | . | . | . | . | . | . |
| $\sum_7$NDL-PCBs | 187 | -0.029 | -0.172 | 0.115 | 0.694 | 179 | -0.048 | -0.192 | 0.095 | 0.512 | -0.018 | -0.152 | 0.115 | 0.786 | . | . | . | . | . | . | . | . | . |
| $\sum_{14}$PCBs | 184 | -0.032 | -0.171 | 0.107 | 0.652 | 176 | -0.054 | -0.193 | 0.085 | 0.447 | -0.021 | -0.155 | 0.113 | 0.758 | 134 | 0.114 | -0.108 | 0.335 | 0.316 | 0.070 | -0.118 | 0.257 | 0.464 |
| $\sum_4$OCPs | 173 | -0.114 | -0.258 | 0.030 | 0.124 | 165 | -0.134 | -0.282 | 0.013 | 0.075 | -0.072 | -0.208 | 0.064 | 0.297 | . | -0.125 | -0.370 | 0.121 | 0.322 | -0.081 | -0.277 | 0.115 | 0.416 |
| $\sum_6$PBDEs | 180 | -0.128 | -0.262 | 0.005 | 0.062 | 172 | -0.123 | -0.256 | 0.010 | 0.071 | -0.064 | -0.201 | 0.072 | 0.352 | . | -0.073 | -0.228 | 0.082 | 0.360 | -0.041 | -0.185 | 0.104 | 0.581 |
| $\sum_2$PFASs | 156 | -0.147 | -0.302 | 0.008 | 0.065 | 151 | -0.127 | -0.283 | 0.030 | 0.114 | -0.111 | -0.251 | 0.028 | 0.118 | . | -0.096 | -0.266 | 0.075 | 0.274 | -0.103 | -0.254 | 0.048 | 0.181 |
| **Propionic** | | | | | | | | | | | | | | | | | | | | | | | |
| **4 months** | | | | | | | | | | | | | | | | | | | | | | | |
| $\sum_7$DL-PCBs | 87 | 0.101 | -0.098 | 0.300 | 0.324 | 74 | 0.040 | -0.172 | 0.252 | 0.712 | 0.098 | -0.074 | 0.270 | 0.259 | . | . | . | . | . | . | . | . | . |
| $\sum_7$NDL-PCBs | 89 | **0.203** | **0.014** | **0.392** | **0.038** | 76 | 0.157 | -0.047 | 0.361 | 0.135 | 0.134 | -0.033 | 0.301 | 0.116 | . | . | . | . | . | . | . | . | . |
| $\sum_{14}$PCBs | 87 | 0.156 | -0.035 | 0.347 | 0.112 | 74 | 0.098 | -0.102 | 0.298 | 0.341 | 0.131 | -0.036 | 0.298 | 0.123 | 65 | 0.116 | -0.131 | 0.363 | 0.363 | 0.174 | -0.066 | 0.414 | 0.154 |
| $\sum_4$OCPs | 89 | 0.203 | -0.018 | 0.423 | 0.075 | 76 | 0.108 | -0.145 | 0.361 | 0.407 | 0.057 | -0.130 | 0.244 | 0.547 | . | -0.273 | -0.601 | 0.055 | 0.108 | -0.106 | -0.375 | 0.164 | 0.438 |
| $\sum_6$PBDEs | 84 | **0.312** | **0.071** | **0.553** | **0.013** | 71 | **0.309** | **0.068** | **0.551** | **0.015** | 0.157 | -0.074 | 0.389 | 0.179 | . | **0.444** | **0.187** | **0.702** | **0.001** | 0.155 | -0.091 | 0.400 | 0.211 |
| $\sum_2$PFASs | 79 | -0.007 | -0.231 | 0.217 | 0.952 | 69 | 0.024 | -0.213 | 0.262 | 0.841 | 0.008 | -0.188 | 0.204 | 0.934 | . | -0.061 | -0.298 | 0.176 | 0.617 | -0.031 | -0.241 | 0.180 | 0.772 |
| **12 months** | | | | | | | | | | | | | | | | | | | | | | | |
| $\sum_7$DL-PCBs | 221 | 0.038 | -0.095 | 0.171 | 0.573 | 196 | 0.008 | -0.122 | 0.137 | 0.909 | 0.051 | -0.080 | 0.182 | 0.444 | . | . | . | . | . | . | . | . | . |
| $\sum_7$NDL-PCBs | 224 | 0.082 | -0.047 | 0.212 | 0.214 | 199 | 0.019 | -0.104 | 0.143 | 0.760 | 0.081 | -0.048 | 0.209 | 0.217 | . | . | . | . | . | . | . | . | . |
| $\sum_{14}$PCBs | 221 | 0.078 | -0.053 | 0.210 | 0.244 | 196 | 0.017 | -0.110 | 0.143 | 0.795 | 0.078 | -0.051 | 0.207 | 0.234 | 157 | 0.119 | -0.085 | 0.323 | 0.256 | 0.160 | -0.013 | 0.333 | 0.069 |
| $\sum_4$OCPs | 213 | -0.024 | -0.156 | 0.108 | 0.722 | 188 | -0.041 | -0.169 | 0.087 | 0.532 | -0.011 | -0.137 | 0.116 | 0.870 | . | -0.128 | -0.346 | 0.089 | 0.249 | -0.117 | -0.293 | 0.058 | 0.188 |
| $\sum_6$PBDEs | 215 | -0.021 | -0.150 | 0.109 | 0.755 | 191 | -0.019 | -0.136 | 0.098 | 0.751 | -0.010 | -0.136 | 0.115 | 0.873 | . | 0.000 | -0.137 | 0.137 | 0.997 | -0.011 | -0.143 | 0.121 | 0.872 |

| Exposure | Unadjusted, single-pollutant | | | | | Adjusted,[a] single-pollutant | | | | | | | | | Adjusted,[a] multi-pollutant | | | | | | | | |
|---|---|---|---|---|---|---|---|---|---|---|---|---|---|---|---|---|---|---|---|---|---|---|---|
| | Complete case | | | | | Complete case | | | | | Multiple imputation[b] (n=298) | | | | Complete case[c] | | | | | Multiple imp.[b] (n=298) | | | |
| (ln-2SD) | n | *β* | 95% CI | | *p*-val | n | *β* | 95% CI | | *p*-val | *β* | 95% CI | | *p*-val | n | *β* | 95% CI | | *p*-val | *β* | 95% CI | | *p*-val |
| $\sum_2$PFASs | 191 | -0.003 | -0.140 | 0.134 | 0.967 | 173 | -0.017 | -0.155 | 0.121 | 0.806 | 0.012 | -0.122 | 0.147 | 0.855 | . | -0.024 | -0.182 | 0.134 | 0.767 | 0.004 | -0.137 | 0.144 | 0.960 |
| **24 months** | | | | | | | | | | | | | | | | | | | | | | | |
| $\sum_7$DL-PCBs | 184 | **-0.180** | **-0.326** | **-0.034** | **0.017** | 176 | **-0.208** | **-0.362** | **-0.054** | **0.009** | -0.106 | -0.253 | 0.041 | 0.156 | . | . | . | . | . | . | . | . | . |
| $\sum_7$NDL-PCBs | 187 | **-0.166** | **-0.307** | **-0.024** | **0.023** | 179 | **-0.180** | **-0.328** | **-0.032** | **0.018** | -0.105 | -0.248 | 0.038 | 0.150 | . | . | . | . | . | . | . | . | . |
| $\sum_{14}$PCBs | 184 | **-0.172** | **-0.316** | **-0.028** | **0.020** | 176 | **-0.190** | **-0.341** | **-0.039** | **0.015** | -0.107 | -0.251 | 0.037 | 0.143 | 134 | -0.026 | -0.257 | 0.205 | 0.826 | -0.029 | -0.238 | 0.179 | 0.780 |
| $\sum_4$OCPs | 173 | **-0.176** | **-0.316** | **-0.036** | **0.015** | 165 | **-0.181** | **-0.330** | **-0.032** | **0.019** | -0.127 | -0.270 | 0.017 | 0.083 | . | -0.096 | -0.352 | 0.161 | 0.466 | -0.090 | -0.309 | 0.130 | 0.421 |
| $\sum_6$PBDEs | 180 | -0.097 | -0.235 | 0.040 | 0.168 | 172 | -0.105 | -0.249 | 0.038 | 0.152 | -0.053 | -0.193 | 0.088 | 0.462 | . | -0.039 | -0.201 | 0.123 | 0.639 | -0.009 | -0.161 | 0.143 | 0.909 |
| $\sum_2$PFASs | 156 | -0.135 | -0.286 | 0.016 | 0.081 | 151 | -0.146 | -0.304 | 0.012 | 0.072 | -0.088 | -0.241 | 0.065 | 0.258 | . | -0.090 | -0.268 | 0.088 | 0.322 | -0.057 | -0.221 | 0.106 | 0.489 |
| ***n*-butyric** | | | | | | | | | | | | | | | | | | | | | | | |
| **4 months** | | | | | | | | | | | | | | | | | | | | | | | |
| $\sum_7$DL-PCBs | 87 | 0.074 | -0.131 | 0.279 | 0.483 | 74 | 0.071 | -0.150 | 0.291 | 0.531 | 0.048 | -0.162 | 0.257 | 0.651 | . | . | . | . | . | . | . | . | . |
| $\sum_7$NDL-PCBs | 89 | 0.048 | -0.146 | 0.242 | 0.628 | 76 | 0.079 | -0.122 | 0.281 | 0.443 | 0.035 | -0.159 | 0.230 | 0.717 | . | . | . | . | . | . | . | . | . |
| $\sum_{14}$PCBs | 87 | 0.068 | -0.130 | 0.266 | 0.504 | 74 | 0.087 | -0.121 | 0.296 | 0.415 | 0.039 | -0.157 | 0.234 | 0.695 | 65 | 0.208 | -0.080 | 0.496 | 0.162 | 0.048 | -0.246 | 0.343 | 0.744 |
| $\sum_4$OCPs | 89 | -0.020 | -0.245 | 0.204 | 0.860 | 76 | -0.029 | -0.278 | 0.219 | 0.819 | 0.005 | -0.204 | 0.214 | 0.965 | . | **-0.447** | **-0.829** | **-0.065** | **0.025** | -0.123 | -0.433 | 0.187 | 0.428 |
| $\sum_6$PBDEs | 84 | **0.296** | **0.046** | **0.545** | **0.023** | 71 | **0.302** | **0.049** | **0.554** | **0.022** | 0.203 | -0.017 | 0.423 | 0.070 | . | **0.445** | **0.145** | **0.744** | **0.005** | 0.217 | -0.015 | 0.450 | 0.066 |
| $\sum_2$PFASs | 79 | 0.177 | -0.079 | 0.434 | 0.179 | 69 | 0.112 | -0.141 | 0.366 | 0.389 | 0.107 | -0.093 | 0.307 | 0.291 | . | 0.064 | -0.212 | 0.340 | 0.653 | 0.094 | -0.122 | 0.311 | 0.387 |
| **12 months** | | | | | | | | | | | | | | | | | | | | | | | |
| $\sum_7$DL-PCBs | 221 | 0.021 | -0.112 | 0.153 | 0.760 | 196 | 0.017 | -0.126 | 0.161 | 0.812 | 0.012 | -0.120 | 0.145 | 0.855 | . | . | . | . | . | . | . | . | . |
| $\sum_7$NDL-PCBs | 224 | 0.039 | -0.091 | 0.169 | 0.560 | 199 | 0.021 | -0.117 | 0.158 | 0.769 | 0.029 | -0.103 | 0.162 | 0.662 | . | . | . | . | . | . | . | . | . |
| $\sum_{14}$PCBs | 221 | 0.043 | -0.089 | 0.175 | 0.522 | 196 | 0.029 | -0.112 | 0.169 | 0.691 | 0.027 | -0.105 | 0.159 | 0.686 | 157 | **0.244** | **0.035** | **0.452** | **0.023** | 0.104 | -0.078 | 0.285 | 0.260 |
| $\sum_4$OCPs | 213 | -0.023 | -0.153 | 0.108 | 0.735 | 188 | -0.127 | -0.267 | 0.012 | 0.075 | -0.043 | -0.177 | 0.091 | 0.528 | . | **-0.339** | **-0.561** | **-0.116** | **0.003** | -0.132 | -0.322 | 0.059 | 0.174 |
| $\sum_6$PBDEs | 215 | 0.024 | -0.102 | 0.150 | 0.708 | 191 | -0.004 | -0.131 | 0.122 | 0.946 | 0.025 | -0.107 | 0.157 | 0.710 | . | 0.091 | -0.049 | 0.231 | 0.205 | 0.040 | -0.098 | 0.179 | 0.567 |
| $\sum_2$PFASs | 191 | 0.011 | -0.127 | 0.149 | 0.874 | 173 | -0.022 | -0.175 | 0.131 | 0.780 | 0.015 | -0.125 | 0.156 | 0.829 | . | -0.025 | -0.186 | 0.137 | 0.766 | 0.017 | -0.131 | 0.164 | 0.825 |
| **24 months** | | | | | | | | | | | | | | | | | | | | | | | |
| $\sum_7$DL-PCBs | 184 | -0.101 | -0.248 | 0.046 | 0.181 | 176 | -0.137 | -0.288 | 0.015 | 0.079 | -0.075 | -0.217 | 0.067 | 0.297 | . | . | . | . | . | . | . | . | . |
| $\sum_7$NDL-PCBs | 187 | -0.074 | -0.217 | 0.069 | 0.311 | 179 | -0.104 | -0.250 | 0.042 | 0.166 | -0.060 | -0.200 | 0.081 | 0.402 | . | . | . | . | . | . | . | . | . |
| $\sum_{14}$PCBs | 184 | -0.080 | -0.225 | 0.065 | 0.282 | 176 | -0.114 | -0.262 | 0.035 | 0.135 | -0.064 | -0.205 | 0.077 | 0.373 | 134 | 0.059 | -0.152 | 0.271 | 0.584 | 0.030 | -0.179 | 0.239 | 0.779 |
| $\sum_4$OCPs | 173 | **-0.143** | **-0.276** | **-0.010** | **0.036** | 165 | **-0.166** | **-0.304** | **-0.028** | **0.020** | -0.112 | -0.253 | 0.029 | 0.119 | . | -0.117 | -0.352 | 0.118 | 0.330 | -0.125 | -0.343 | 0.093 | 0.257 |
| $\sum_6$PBDEs | 180 | -0.069 | -0.209 | 0.071 | 0.336 | 172 | -0.065 | -0.208 | 0.077 | 0.372 | -0.034 | -0.169 | 0.101 | 0.622 | . | -0.020 | -0.169 | 0.128 | 0.788 | 0.004 | -0.141 | 0.149 | 0.956 |
| $\sum_2$PFASs | 156 | -0.066 | -0.201 | 0.070 | 0.345 | 151 | -0.060 | -0.200 | 0.080 | 0.401 | -0.056 | -0.208 | 0.097 | 0.473 | . | -0.029 | -0.192 | 0.134 | 0.730 | -0.033 | -0.192 | 0.127 | 0.688 |
| ***i*-butyric** | | | | | | | | | | | | | | | | | | | | | | | |
| **4 months** | | | | | | | | | | | | | | | | | | | | | | | |
| $\sum_7$DL-PCBs | 87 | 0.180 | -0.022 | 0.383 | 0.084 | 74 | 0.111 | -0.107 | 0.329 | 0.321 | 0.131 | -0.074 | 0.337 | 0.207 | . | . | . | . | . | . | . | . | . |
| $\sum_7$NDL-PCBs | 89 | 0.176 | -0.014 | 0.367 | 0.073 | 76 | 0.131 | -0.069 | 0.332 | 0.204 | 0.121 | -0.075 | 0.318 | 0.221 | . | . | . | . | . | . | . | . | . |
| $\sum_{14}$PCBs | 87 | 0.175 | -0.021 | 0.371 | 0.083 | 74 | 0.117 | -0.090 | 0.324 | 0.271 | 0.125 | -0.072 | 0.323 | 0.210 | 65 | 0.076 | -0.215 | 0.367 | 0.610 | 0.142 | -0.147 | 0.432 | 0.329 |
| $\sum_4$OCPs | 89 | 0.183 | -0.038 | 0.404 | 0.108 | 76 | 0.091 | -0.157 | 0.340 | 0.473 | 0.068 | -0.152 | 0.289 | 0.537 | . | -0.118 | -0.504 | 0.268 | 0.552 | -0.037 | -0.353 | 0.278 | 0.814 |
| $\sum_6$PBDEs | 84 | 0.175 | -0.078 | 0.427 | 0.179 | 71 | 0.193 | -0.064 | 0.450 | 0.146 | 0.031 | -0.203 | 0.264 | 0.793 | . | 0.230 | -0.073 | 0.533 | 0.142 | 0.005 | -0.241 | 0.252 | 0.966 |
| $\sum_2$PFASs | 79 | 0.118 | -0.115 | 0.352 | 0.324 | 69 | 0.110 | -0.135 | 0.354 | 0.383 | 0.054 | -0.166 | 0.275 | 0.624 | . | 0.083 | -0.196 | 0.363 | 0.560 | 0.027 | -0.208 | 0.263 | 0.817 |
| **12 months** | | | | | | | | | | | | | | | | | | | | | | | |
| $\sum_7$DL-PCBs | 221 | -0.017 | -0.150 | 0.116 | 0.798 | 196 | -0.090 | -0.231 | 0.051 | 0.210 | -0.005 | -0.134 | 0.125 | 0.945 | . | . | . | . | . | . | . | . | . |
| $\sum_7$NDL-PCBs | 224 | -0.003 | -0.133 | 0.127 | 0.966 | 199 | -0.091 | -0.225 | 0.044 | 0.189 | 0.009 | -0.118 | 0.135 | 0.892 | . | . | . | . | . | . | . | . | . |
| $\sum_{14}$PCBs | 221 | -0.002 | -0.134 | 0.130 | 0.976 | 196 | -0.091 | -0.229 | 0.047 | 0.196 | 0.007 | -0.120 | 0.134 | 0.914 | 157 | -0.039 | -0.249 | 0.170 | 0.713 | 0.013 | -0.162 | 0.187 | 0.887 |

| Exposure | Unadjusted, single-pollutant | | | | | Adjusted,[a] single-pollutant | | | | | | | | | Adjusted,[a] multi-pollutant | | | | | | | | |
|---|---|---|---|---|---|---|---|---|---|---|---|---|---|---|---|---|---|---|---|---|---|---|---|
| | Complete case | | | | | Complete case | | | | | Multiple imputation[b] (n=298) | | | | Complete case[c] | | | | | Multiple imp.[b] (n=298) | | | |
| (ln-2SD) | n | *β* | 95% CI | | *p*-val | n | *β* | 95% CI | | *p*-val | *β* | 95% CI | | *p*-val | n | *β* | 95% CI | | *p*-val | *β* | 95% CI | | *p*-val |
| $\sum_4$OCPs | 213 | -0.012 | -0.144 | 0.120 | 0.854 | 188 | -0.071 | -0.211 | 0.069 | 0.324 | 0.002 | -0.124 | 0.128 | 0.978 | . | -0.089 | -0.312 | 0.134 | 0.437 | -0.019 | -0.198 | 0.160 | 0.835 |
| $\sum_6$PBDEs | 215 | 0.032 | -0.091 | 0.155 | 0.611 | 191 | 0.029 | -0.092 | 0.151 | 0.639 | 0.053 | -0.079 | 0.184 | 0.433 | . | 0.058 | -0.083 | 0.198 | 0.422 | 0.059 | -0.080 | 0.199 | 0.402 |
| $\sum_2$PFASs | 191 | -0.029 | -0.168 | 0.111 | 0.687 | 173 | -0.029 | -0.179 | 0.122 | 0.709 | -0.022 | -0.154 | 0.110 | 0.743 | . | 0.011 | -0.151 | 0.173 | 0.898 | -0.029 | -0.168 | 0.110 | 0.682 |
| **24 months** | | | | | | | | | | | | | | | | | | | | | | | |
| $\sum_7$DL-PCBs | 184 | -0.142 | -0.289 | 0.006 | 0.062 | 176 | -0.146 | -0.296 | 0.004 | 0.059 | -0.103 | -0.248 | 0.041 | 0.160 | . | . | . | . | . | . | . | . | . |
| $\sum_7$NDL-PCBs | 187 | -0.093 | -0.236 | 0.050 | 0.203 | 179 | -0.113 | -0.257 | 0.032 | 0.128 | -0.067 | -0.208 | 0.074 | 0.351 | . | . | . | . | . | . | . | . | . |
| $\sum_{14}$PCBs | 184 | -0.099 | -0.245 | 0.047 | 0.185 | 176 | -0.118 | -0.265 | 0.029 | 0.118 | -0.074 | -0.216 | 0.068 | 0.303 | 134 | -0.108 | -0.335 | 0.119 | 0.351 | -0.072 | -0.278 | 0.135 | 0.494 |
| $\sum_4$OCPs | 173 | -0.068 | -0.209 | 0.072 | 0.342 | 165 | -0.077 | -0.222 | 0.068 | 0.301 | -0.046 | -0.187 | 0.096 | 0.525 | . | 0.145 | -0.106 | 0.397 | 0.260 | 0.030 | -0.188 | 0.248 | 0.787 |
| $\sum_6$PBDEs | 180 | -0.121 | -0.260 | 0.018 | 0.090 | 172 | -0.111 | -0.251 | 0.030 | 0.124 | -0.053 | -0.198 | 0.092 | 0.473 | . | -0.120 | -0.279 | 0.039 | 0.143 | -0.038 | -0.196 | 0.121 | 0.640 |
| $\sum_2$PFASs | 156 | -0.111 | -0.266 | 0.043 | 0.160 | 151 | -0.134 | -0.292 | 0.023 | 0.097 | -0.073 | -0.217 | 0.071 | 0.317 | . | -0.152 | -0.327 | 0.023 | 0.092 | -0.058 | -0.212 | 0.096 | 0.461 |
| ***n*-valeric** | | | | | | | | | | | | | | | | | | | | | | | |
| **4 months** | | | | | | | | | | | | | | | | | | | | | | | |
| $\sum_7$DL-PCBs | 87 | -0.048 | -0.254 | 0.158 | 0.650 | 74 | -0.050 | -0.299 | 0.200 | 0.698 | 0.077 | -0.134 | 0.287 | 0.469 | . | . | . | . | . | . | . | . | . |
| $\sum_7$NDL-PCBs | 89 | -0.050 | -0.244 | 0.144 | 0.612 | 76 | -0.083 | -0.311 | 0.145 | 0.477 | 0.073 | -0.129 | 0.275 | 0.474 | . | . | . | . | . | . | . | . | . |
| $\sum_{14}$PCBs | 87 | -0.042 | -0.241 | 0.157 | 0.679 | 74 | -0.067 | -0.304 | 0.169 | 0.578 | 0.076 | -0.128 | 0.279 | 0.461 | 65 | -0.097 | -0.363 | 0.169 | 0.477 | 0.130 | -0.152 | 0.413 | 0.360 |
| $\sum_4$OCPs | 89 | 0.049 | -0.175 | 0.273 | 0.667 | 76 | 0.016 | -0.265 | 0.298 | 0.910 | 0.011 | -0.213 | 0.235 | 0.921 | . | 0.060 | -0.293 | 0.412 | 0.742 | -0.086 | -0.397 | 0.226 | 0.584 |
| $\sum_6$PBDEs | 84 | 0.189 | -0.066 | 0.444 | 0.149 | 71 | 0.206 | -0.089 | 0.501 | 0.176 | 0.038 | -0.199 | 0.274 | 0.750 | . | 0.226 | -0.050 | 0.503 | 0.114 | 0.037 | -0.213 | 0.288 | 0.765 |
| $\sum_2$PFASs | 79 | 0.017 | -0.184 | 0.217 | 0.871 | 69 | -0.022 | -0.245 | 0.200 | 0.847 | -0.001 | -0.226 | 0.224 | 0.993 | . | -0.042 | -0.297 | 0.213 | 0.750 | -0.018 | -0.252 | 0.217 | 0.880 |
| **12 months** | | | | | | | | | | | | | | | | | | | | | | | |
| $\sum_7$DL-PCBs | 221 | -0.032 | -0.165 | 0.100 | 0.632 | 196 | -0.035 | -0.172 | 0.103 | 0.622 | 0.028 | -0.102 | 0.158 | 0.669 | . | . | . | . | . | . | . | . | . |
| $\sum_7$NDL-PCBs | 224 | 0.007 | -0.123 | 0.137 | 0.915 | 199 | -0.021 | -0.153 | 0.111 | 0.758 | 0.051 | -0.080 | 0.182 | 0.444 | . | . | . | . | . | . | . | . | . |
| $\sum_{14}$PCBs | 221 | 0.002 | -0.130 | 0.134 | 0.978 | 196 | -0.026 | -0.160 | 0.109 | 0.710 | 0.049 | -0.082 | 0.179 | 0.461 | 157 | -0.107 | -0.288 | 0.074 | 0.250 | 0.144 | -0.040 | 0.329 | 0.125 |
| $\sum_4$OCPs | 213 | -0.061 | -0.192 | 0.070 | 0.362 | 188 | -0.056 | -0.191 | 0.079 | 0.418 | -0.046 | -0.174 | 0.082 | 0.482 | . | 0.115 | -0.078 | 0.307 | 0.246 | -0.165 | -0.354 | 0.025 | 0.089 |
| $\sum_6$PBDEs | 215 | -0.028 | -0.154 | 0.099 | 0.669 | 191 | -0.044 | -0.165 | 0.077 | 0.478 | 0.027 | -0.108 | 0.162 | 0.694 | . | -0.029 | -0.151 | 0.092 | 0.636 | 0.042 | -0.101 | 0.184 | 0.565 |
| $\sum_2$PFASs | 191 | 0.048 | -0.079 | 0.175 | 0.463 | 173 | 0.002 | -0.130 | 0.135 | 0.972 | 0.028 | -0.117 | 0.174 | 0.702 | . | -0.039 | -0.179 | 0.101 | 0.583 | 0.028 | -0.123 | 0.178 | 0.719 |
| **24 months** | | | | | | | | | | | | | | | | | | | | | | | |
| $\sum_7$DL-PCBs | 184 | -0.134 | -0.282 | 0.014 | 0.078 | 176 | -0.143 | -0.298 | 0.011 | 0.071 | -0.080 | -0.231 | 0.071 | 0.298 | . | . | . | . | . | . | . | . | . |
| $\sum_7$NDL-PCBs | 187 | **-0.148** | **-0.290** | **-0.006** | **0.042** | 179 | **-0.164** | **-0.311** | **-0.018** | **0.029** | -0.093 | -0.241 | 0.056 | 0.219 | . | . | . | . | . | . | . | . | . |
| $\sum_{14}$PCBs | 184 | **-0.147** | **-0.292** | **-0.002** | **0.048** | 176 | **-0.165** | **-0.316** | **-0.015** | **0.032** | -0.092 | -0.242 | 0.057 | 0.223 | 134 | -0.112 | -0.328 | 0.104 | 0.311 | -0.034 | -0.247 | 0.179 | 0.753 |
| $\sum_4$OCPs | 173 | **-0.161** | **-0.300** | **-0.021** | **0.025** | 165 | **-0.175** | **-0.321** | **-0.028** | **0.021** | -0.111 | -0.258 | 0.037 | 0.139 | . | 0.051 | -0.189 | 0.290 | 0.679 | -0.103 | -0.321 | 0.115 | 0.352 |
| $\sum_6$PBDEs | 180 | -0.065 | -0.205 | 0.075 | 0.364 | 172 | -0.082 | -0.226 | 0.062 | 0.267 | -0.001 | -0.142 | 0.139 | 0.987 | . | -0.034 | -0.186 | 0.117 | 0.658 | 0.038 | -0.110 | 0.186 | 0.610 |
| $\sum_2$PFASs | 156 | -0.097 | -0.242 | 0.049 | 0.195 | 151 | -0.107 | -0.258 | 0.045 | 0.169 | -0.017 | -0.175 | 0.140 | 0.829 | . | -0.085 | -0.252 | 0.081 | 0.317 | 0.011 | -0.155 | 0.177 | 0.897 |
| ***i*-valeric** | | | | | | | | | | | | | | | | | | | | | | | |
| **4 months** | | | | | | | | | | | | | | | | | | | | | | | |
| $\sum_7$DL-PCBs | 87 | 0.156 | -0.048 | 0.359 | 0.137 | 74 | 0.087 | -0.138 | 0.311 | 0.451 | 0.140 | -0.054 | 0.335 | 0.155 | . | . | . | . | . | . | . | . | . |
| $\sum_7$NDL-PCBs | 89 | 0.166 | -0.025 | 0.357 | 0.092 | 76 | 0.123 | -0.084 | 0.330 | 0.247 | 0.145 | -0.042 | 0.333 | 0.127 | . | . | . | . | . | . | . | . | . |
| $\sum_{14}$PCBs | 87 | 0.157 | -0.040 | 0.353 | 0.121 | 74 | 0.098 | -0.114 | 0.311 | 0.368 | 0.147 | -0.041 | 0.336 | 0.123 | 65 | 0.077 | -0.224 | 0.378 | 0.618 | 0.147 | -0.155 | 0.449 | 0.333 |
| $\sum_4$OCPs | 89 | 0.181 | -0.040 | 0.402 | 0.112 | 76 | 0.090 | -0.166 | 0.346 | 0.492 | 0.092 | -0.117 | 0.301 | 0.382 | . | -0.184 | -0.583 | 0.215 | 0.371 | -0.066 | -0.397 | 0.265 | 0.691 |
| $\sum_6$PBDEs | 84 | 0.224 | -0.025 | 0.473 | 0.082 | 71 | 0.239 | -0.022 | 0.499 | 0.078 | 0.168 | -0.076 | 0.411 | 0.173 | . | 0.294 | -0.019 | 0.607 | 0.071 | 0.148 | -0.112 | 0.409 | 0.258 |
| $\sum_2$PFASs | 79 | 0.126 | -0.116 | 0.368 | 0.312 | 69 | 0.101 | -0.156 | 0.359 | 0.443 | 0.079 | -0.138 | 0.295 | 0.468 | . | 0.085 | -0.204 | 0.373 | 0.567 | 0.037 | -0.192 | 0.267 | 0.746 |
| **12 months** | | | | | | | | | | | | | | | | | | | | | | | |
| $\sum_7$DL-PCBs | 221 | -0.042 | -0.175 | 0.091 | 0.537 | 196 | -0.119 | -0.262 | 0.024 | 0.106 | -0.021 | -0.151 | 0.110 | 0.753 | . | . | . | . | . | . | . | . | . |

| | Unadjusted, single-pollutant | | | | | Adjusted,[a] single-pollutant | | | | | | | | | Adjusted,[a] multi-pollutant | | | | | | | | |
|---|---|---|---|---|---|---|---|---|---|---|---|---|---|---|---|---|---|---|---|---|---|---|---|
| **Exposure** | **Complete case** | | | | | **Complete case** | | | | | **Multiple imputation[b] (n=298)** | | | | **Complete case[c]** | | | | | **Multiple imp.[b] (n=298)** | | | |
| **(ln-2SD)** | **n** | ***β*** | **95% CI** | | ***p*-val** | **n** | ***β*** | **95% CI** | | ***p*-val** | ***β*** | **95% CI** | | ***p*-val** | **n** | ***β*** | **95% CI** | | ***p*-val** | ***β*** | **95% CI** | | ***p*-val** |
| $\sum_7$NDL-PCBs | 224 | -0.034 | -0.164 | 0.096 | 0.611 | 199 | -0.123 | -0.260 | 0.013 | 0.079 | -0.015 | -0.142 | 0.113 | 0.819 | . | . | . | . | . | . | . | . | . |
| $\sum_{14}$PCBs | 221 | -0.034 | -0.165 | 0.098 | 0.619 | 196 | -0.125 | -0.265 | 0.015 | 0.081 | -0.016 | -0.143 | 0.112 | 0.811 | 157 | -0.092 | -0.304 | 0.121 | 0.399 | -0.024 | -0.199 | 0.152 | 0.789 |
| $\sum_4$OCPs | 213 | -0.017 | -0.150 | 0.115 | 0.798 | 188 | -0.073 | -0.217 | 0.071 | 0.322 | -0.002 | -0.129 | 0.125 | 0.976 | . | -0.044 | -0.270 | 0.182 | 0.704 | 0.005 | -0.174 | 0.184 | 0.957 |
| $\sum_6$PBDEs | 215 | 0.024 | -0.099 | 0.147 | 0.702 | 191 | 0.016 | -0.107 | 0.140 | 0.797 | 0.047 | -0.085 | 0.180 | 0.480 | . | 0.043 | -0.099 | 0.186 | 0.554 | 0.056 | -0.083 | 0.195 | 0.428 |
| $\sum_2$PFASs | 191 | -0.047 | -0.186 | 0.093 | 0.512 | 173 | -0.037 | -0.190 | 0.115 | 0.632 | -0.032 | -0.166 | 0.102 | 0.638 | . | 0.002 | -0.162 | 0.166 | 0.984 | -0.035 | -0.176 | 0.105 | 0.620 |
| **24 months** | | | | | | | | | | | | | | | | | | | | | | | |
| $\sum_7$DL-PCBs | 184 | -0.112 | -0.260 | 0.036 | 0.140 | 176 | -0.111 | -0.261 | 0.040 | 0.153 | -0.077 | -0.228 | 0.073 | 0.312 | . | . | . | . | . | . | . | . | . |
| $\sum_7$NDL-PCBs | 187 | -0.074 | -0.217 | 0.069 | 0.312 | 179 | -0.089 | -0.234 | 0.055 | 0.227 | -0.047 | -0.195 | 0.100 | 0.528 | . | . | . | . | . | . | . | . | . |
| $\sum_{14}$PCBs | 184 | -0.077 | -0.223 | 0.069 | 0.303 | 176 | -0.091 | -0.238 | 0.057 | 0.230 | -0.053 | -0.202 | 0.095 | 0.479 | 134 | -0.090 | -0.312 | 0.132 | 0.428 | -0.040 | -0.251 | 0.171 | 0.708 |
| $\sum_4$OCPs | 173 | -0.066 | -0.208 | 0.076 | 0.362 | 165 | -0.063 | -0.209 | 0.083 | 0.399 | -0.041 | -0.188 | 0.107 | 0.586 | . | 0.162 | -0.084 | 0.408 | 0.199 | 0.010 | -0.206 | 0.225 | 0.930 |
| $\sum_6$PBDEs | 180 | -0.116 | -0.255 | 0.023 | 0.104 | 172 | -0.111 | -0.251 | 0.029 | 0.123 | -0.048 | -0.190 | 0.094 | 0.505 | . | -0.149 | -0.305 | 0.007 | 0.063 | -0.035 | -0.185 | 0.115 | 0.645 |
| $\sum_2$PFASs | 156 | -0.099 | -0.250 | 0.051 | 0.199 | 151 | -0.119 | -0.272 | 0.035 | 0.131 | -0.058 | -0.209 | 0.093 | 0.446 | . | -0.145 | -0.316 | 0.026 | 0.099 | -0.046 | -0.206 | 0.114 | 0.570 |

Abbreviations: PBDE, polybrominated diphenyl ether; CI, confidence interval; LRTI, lower respiratory tract infection; OCP, organochlorine pesticides; PCB, polychlorinated biphenyls; PFAS, poly-and perfluoroalkyl substances.

Regression coefficients were estimated from unpenalized linear regression models; β (95% CI) represent a 2-standard deviation change in untransformed diversity or absolute SCFA levels (mmol/kg) per 2-standard deviation increase in ln-transformed chemical levels (refer to Tables S5 and S9 for the increments). Summed-PBDEs, PCBs, OCPs and PBDEs are in units of pmol/g and summed-PFASs in pg/L.

[a] Models were adjusted for maternal education, breastfeeding duration, C-section, and recent antibiotic use (refer to Figure S4C).

[b] Multiple imputation models: n=100 imputed datasets (n=298). Data was missing for 32.6–43.2% of the chemical data; 11.0–31.3% of the Shannon diversity and 26.0–68.0% of the SCFA data; and 0.0–18.9% of the confounder data.

[c] For multi-pollutant models, VIFs for exposures were <3, but highest (~2.3) for the $\sum_{14}$PCBs and $\sum_4$OCPs and lower (~1.2) for $\sum_6$PBDEs and $\sum_2$PFASs.

**Table S12.** Associations between individual POP exposures and Shannon diversity and SCFAs: penalized elastic net regression models (n=298).

| | Diversity | | | | | | Total SCFAs | | | | | |
|---|---|---|---|---|---|---|---|---|---|---|---|---|
| | 4 mo. | | 12 mo. | | 24 mo. | | 4 mo. | | 12 mo. | | 24 mo. | |
| Exposure | % | β[a] | % | β | % | OR | % | β | % | β | % | β |
| PCB-74 | 14 | 0 | 29 | 0 | 66 | -0.149 | 8 | 0 | 0 | 0 | 42 | 0 |
| PCB-99 | 12 | 0 | 4 | 0 | 31 | 0 | 3 | 0 | 0 | 0 | 10 | 0 |
| PCB-105 | 17 | 0 | 17 | 0 | 50 | -0.133 | 9 | 0 | 1 | 0 | 1 | 0 |
| PCB-114 | 35 | 0 | **77** | **-0.069** | **72** | **-0.012** | 19 | 0 | 25 | 0 | 17 | 0 |
| PCB-118 | 11 | 0 | 1 | 0 | 39 | 0 | 9 | 0 | 0 | 0 | 10 | 0 |
| PCB-138 | 8 | 0 | 30 | 0 | 67 | -0.342 | 7 | 0 | 0 | 0 | 3 | 0 |
| PCB-153 | 29 | 0 | 1 | 0 | 23 | 0 | 1 | 0 | 0 | 0 | 0 | 0 |
| PCB-156 | 23 | 0 | 15 | 0 | 34 | 0 | 7 | 0 | 0 | 0 | 0 | 0 |
| PCB-157 | 31 | 0 | 32 | 0 | 27 | 0 | 5 | 0 | 3 | 0 | 2 | 0 |
| PCB-167 | 24 | 0 | 23 | 0 | 34 | 0 | 4 | 0 | 0 | 0 | 1 | 0 |
| PCB-170 | 4 | 0 | 13 | 0 | 45 | 0 | 21 | 0 | 44 | 0 | 1 | 0 |
| PCB-180 | 31 | 0 | 2 | 0 | 13 | 0 | 6 | 0 | 15 | 0 | 4 | 0 |
| PCB-189 | 25 | 0 | 30 | 0 | 66 | 0.065 | 15 | 0 | 10 | 0 | 11 | 0 |
| PCB-194 | 30 | 0 | 51 | 0.049 | 58 | -0.001 | 33 | 0 | 26 | 0 | 17 | 0 |
| HCB | 33 | 0 | 31 | 0 | 65 | 0.005 | 15 | 0 | 0 | 0 | 17 | 0 |
| β-HCH | 30 | 0 | **84** | **-0.072** | **75** | **0.171** | 13 | 0 | 0 | 0 | 2 | 0 |
| oxychlor. | 32 | 0 | 30 | 0 | 48 | 0 | 17 | 0 | 1 | 0 | 17 | 0 |
| DDE | 16 | 0 | **71** | **-0.091** | 48 | 0 | 13 | 0 | 0 | 0 | 41 | 0 |
| BDE-28 | 35 | 0 | 26 | 0 | 61 | 0.038 | 15 | 0 | 1 | 0 | 25 | 0 |
| BDE-47 | 29 | 0 | 38 | 0 | 42 | 0 | 8 | 0 | 1 | 0 | 7 | 0 |
| BDE-99 | 19 | 0 | 19 | 0 | 48 | 0 | 12 | 0 | 0 | 0 | 46 | 0 |
| BDE-100 | 16 | 0 | 10 | 0 | 48 | 0 | 13 | 0 | 0 | 0 | 6 | 0 |
| BDE-153 | 24 | 0 | 36 | 0 | 58 | -0.065 | 15 | 0 | 1 | 0 | 8 | 0 |
| BDE-154 | 26 | 0 | 42 | 0 | 53 | 0.020 | 8 | 0 | 0 | 0 | 13 | 0 |
| PFOA | 42 | 0 | 67 | -0.085 | **83** | **0.181** | 26 | 0 | 18 | 0 | 59 | -3.732 |
| PFOS | 39 | 0 | 25 | 0 | **78** | **-0.153** | 36 | 0 | 1 | 0 | 18 | 0 |

**Table S12.** *Continued*

| | Acetic | | | | | | Propionic | | | | | | *n*-butyric | | | | | | *i*-butyric | | | | | | *n*-valeric | | | | | | *i*-valeric | | | | | |
|---|---|---|---|---|---|---|---|---|---|---|---|---|---|---|---|---|---|---|---|---|---|---|---|---|---|---|---|---|---|---|---|---|---|---|---|---|
| | 4 mo. | | 12 mo. | | 24 mo. | | 4 mo. | | 12 mo. | | 24 mo. | | 4 mo. | | 12 mo. | | 24 mo. | | 4 mo. | | 12 mo. | | 24 mo. | | 4 mo. | | 12 mo. | | 24 mo. | | 4 mo. | | 12 mo. | | 24 mo. | |
| **Exposure** | **%** | **β** | **%** | **β** | **%** | **β** | **%** | **β** | **%** | **β** | **%** | **β** | **%** | **β** | **%** | **β** | **%** | **β** | **%** | **β** | **%** | **β** | **%** | **β** | **%** | **β** | **%** | **β** | **%** | **β** | **%** | **β** | **%** | **β** | **%** | **β** |
| PCB-74 | 9 | 0 | 0 | 0 | 54 | -5.278 | 39 | 0 | 3 | 0 | 42 | 0 | 45 | 0 | 6 | 0 | 4 | 0 | 48 | 0 | 1 | 0 | 30 | 0 | 42 | 0 | 44 | 0 | 42 | 0 | 42 | 0 | 0 | 0 | 40 | 0 |
| PCB-99 | 2 | 0 | 0 | 0 | 29 | 0 | 16 | 0 | 1 | 0 | 2 | 0 | 38 | 0 | 0 | 0 | 1 | 0 | 19 | 0 | 0 | 0 | 7 | 0 | 24 | 0 | 27 | 0 | 1 | 0 | 29 | 0 | 0 | 0 | 10 | 0 |
| PCB-105 | 19 | 0 | 0 | 0 | 3 | 0 | 23 | 0 | 2 | 0 | 2 | 0 | 43 | 0 | 1 | 0 | 0 | 0 | 45 | 0 | 2 | 0 | 2 | 0 | 32 | 0 | 34 | 0 | 12 | 0 | 38 | 0 | 0 | 0 | 6 | 0 |
| PCB-114 | 21 | 0 | 12 | 0 | 25 | 0 | 52 | -0.305 | 3 | 0 | 21 | 0 | 56 | -0.355 | 8 | 0 | 18 | 0 | 38 | 0 | 8 | 0 | 28 | 0 | 52 | -0.017 | 54 | -0.226 | 23 | 0 | 53 | -0.100 | 7 | 0 | 27 | 0 |
| PCB-118 | 7 | 0 | 0 | 0 | 11 | 0 | 29 | 0 | 0 | 0 | 3 | 0 | 24 | 0 | 0 | 0 | 7 | 0 | 23 | 0 | 0 | 0 | 20 | 0 | 26 | 0 | 6 | 0 | 0 | 0 | 16 | 0 | 0 | 0 | 3 | 0 |
| PCB-138 | 2 | 0 | 0 | 0 | 10 | 0 | **77** | 5.500 | 3 | 0 | 0 | 0 | 34 | 0 | 0 | 0 | 2 | 0 | 48 | 0 | 1 | 0 | 0 | 0 | 35 | 0 | 7 | 0 | 2 | 0 | 29 | 0 | 0 | 0 | 3 | 0 |
| PCB-153 | 2 | 0 | 0 | 0 | 0 | 0 | 14 | 0 | 0 | 0 | 1 | 0 | 31 | 0 | 0 | 0 | 0 | 0 | 19 | 0 | 0 | 0 | 3 | 0 | 15 | 0 | 14 | 0 | 3 | 0 | 12 | 0 | 0 | 0 | 18 | 0 |
| PCB-156 | 4 | 0 | 0 | 0 | 0 | 0 | 15 | 0 | 2 | 0 | 3 | 0 | 32 | 0 | 0 | 0 | 4 | 0 | 23 | 0 | 1 | 0 | 7 | 0 | 22 | 0 | 44 | 0 | 10 | 0 | 19 | 0 | 0 | 0 | 3 | 0 |
| PCB-157 | 12 | 0 | 0 | 0 | 12 | 0 | 47 | 0 | 1 | 0 | 6 | 0 | 46 | 0 | 3 | 0 | 2 | 0 | 47 | 0 | 0 | 0 | 8 | 0 | **75** | 0.130 | 59 | 0.353 | 4 | 0 | 36 | 0 | 0 | 0 | 6 | 0 |
| PCB-167 | 4 | 0 | 0 | 0 | 1 | 0 | 30 | 0 | 4 | 0 | 2 | 0 | 41 | 0 | 1 | 0 | 3 | 0 | 35 | 0 | 2 | 0 | 11 | 0 | 31 | 0 | 49 | 0 | 0 | 0 | 43 | 0 | 0 | 0 | 3 | 0 |
| PCB-170 | 20 | 0 | 22 | 0 | 4 | 0 | 13 | 0 | 27 | 0 | 3 | 0 | 44 | 0 | 1 | 0 | 0 | 0 | 18 | 0 | 1 | 0 | 0 | 0 | 28 | 0 | 19 | 0 | 2 | 0 | 18 | 0 | 0 | 0 | 2 | 0 |
| PCB-180 | 9 | 0 | 18 | 0 | 2 | 0 | 29 | 0 | 0 | 0 | 10 | 0 | 31 | 0 | 0 | 0 | 3 | 0 | 26 | 0 | 0 | 0 | 6 | 0 | 33 | 0 | 24 | 0 | 15 | 0 | 21 | 0 | 0 | 0 | 3 | 0 |
| PCB-189 | 14 | 0 | 0 | 0 | 34 | 0 | 45 | 0 | 29 | 0 | 10 | 0 | 66 | 0.628 | 3 | 0 | 5 | 0 | 52 | 0.038 | 2 | 0 | 28 | 0 | 45 | 0 | 35 | 0 | 13 | 0 | 60 | 0.115 | 3 | 0 | 31 | 0 |
| PCB-194 | 16 | 0 | 0 | 0 | 52 | 1.509 | 41 | 0 | 31 | 0 | 2 | 0 | 52 | 0.343 | 16 | 0 | 1 | 0 | 69 | 0.134 | 1 | 0 | 5 | 0 | 65 | 0.020 | 41 | 0 | 14 | 0 | **85** | **0.236** | 0 | 0 | 14 | 0 |
| HCB | 21 | 0 | 0 | 0 | 40 | 0 | 50 | -1.179 | 3 | 0 | 10 | 0 | 68 | 0.479 | 1 | 0 | 3 | 0 | 47 | 0 | 0 | 0 | 13 | 0 | 51 | 0.015 | 38 | 0 | 23 | 0 | 47 | 0 | 0 | 0 | 18 | 0 |
| β-HCH | 14 | 0 | 0 | 0 | 14 | 0 | 36 | 0 | 2 | 0 | 23 | 0 | 52 | -0.171 | 8 | 0 | 3 | 0 | 47 | 0 | 5 | 0 | 14 | 0 | 37 | 0 | 42 | 0 | 9 | 0 | 33 | 0 | 0 | 0 | 17 | 0 |
| oxychlor. | 18 | 0 | 0 | 0 | 26 | 0 | 44 | 0 | 3 | 0 | 8 | 0 | 65 | 0.151 | 7 | 0 | 10 | 0 | 55 | 0.002 | 0 | 0 | 14 | 0 | 60 | 0.001 | 41 | 0 | 14 | 0 | 46 | 0 | 0 | 0 | 19 | 0 |
| DDE | 14 | 0 | 0 | 0 | 21 | 0 | 60 | -1.800 | 9 | 0 | 29 | 0 | **77** | **-1.118** | 4 | 0 | 53 | -0.984 | 60 | -0.021 | 1 | 0 | 5 | 0 | 58 | -0.019 | 47 | 0 | 29 | 0 | 53 | -0.090 | 0 | 0 | 10 | 0 |
| BDE-28 | 8 | 0 | 0 | 0 | 44 | 0 | 41 | 0 | 1 | 0 | 4 | 0 | **75** | **1.333** | 4 | 0 | 10 | 0 | 61 | 0.100 | 2 | 0 | 10 | 0 | 65 | -0.021 | 35 | 0 | 21 | 0 | 54 | 0.127 | 3 | 0 | 21 | 0 |
| BDE-47 | 17 | 0 | 0 | 0 | 14 | 0 | 69 | 2.491 | 1 | 0 | 7 | 0 | 41 | 0 | 1 | 0 | 8 | 0 | 43 | 0 | 2 | 0 | 12 | 0 | 44 | 0 | 24 | 0 | 3 | 0 | 44 | 0 | 2 | 0 | 14 | 0 |
| BDE-99 | 13 | 0 | 0 | 0 | 60 | -2.626 | 47 | 0 | 3 | 0 | 23 | 0 | 91 | 2.906 | 1 | 0 | 8 | 0 | 43 | 0 | 3 | 0 | 31 | 0 | 53 | 0.005 | 24 | 0 | 6 | 0 | 66 | 0.159 | 2 | 0 | 29 | 0 |
| BDE-100 | 29 | 0 | 0 | 0 | 14 | 0 | 34 | 0 | 8 | 0 | 1 | 0 | 47 | 0 | 2 | 0 | 2 | 0 | 35 | 0 | 2 | 0 | 3 | 0 | 32 | 0 | 30 | 0 | 11 | 0 | 41 | 0 | 0 | 0 | 7 | 0 |
| BDE-153 | 21 | 0 | 0 | 0 | 24 | 0 | 47 | 0 | 3 | 0 | 5 | 0 | 47 | 0 | 3 | 0 | 24 | 0 | 48 | 0 | 3 | 0 | 20 | 0 | 39 | 0 | 32 | 0 | 7 | 0 | 46 | 0 | 0 | 0 | 21 | 0 |
| BDE-154 | 16 | 0 | 0 | 0 | 20 | 0 | 40 | 0 | 3 | 0 | 27 | 0 | 44 | 0 | 2 | 0 | 6 | 0 | 41 | 0 | 2 | 0 | 8 | 0 | 41 | 0 | 24 | 0 | 6 | 0 | 37 | 0 | 0 | 0 | 12 | 0 |
| PFOA | 37 | 0 | 5 | 0 | **80** | **-5.095** | 53 | 0.534 | 19 | 0 | 17 | 0 | 67 | 0.343 | 6 | 0 | 6 | 0 | 64 | 0.105 | 4 | 0 | 11 | 0 | 60 | -0.004 | 44 | 0 | 19 | 0 | 64 | 0.192 | 6 | 0 | 20 | 0 |
| PFOS | 38 | 0 | 6 | 0 | 19 | 0 | 60 | -1.019 | 8 | 0 | 19 | 0 | 78 | 0.846 | 2 | 0 | 12 | 0 | 56 | -0.041 | 4 | 0 | 39 | 0 | 60 | -0.007 | 41 | 0 | 11 | 0 | 45 | 0 | 8 | 0 | 37 | 0 |

Regression coefficients were estimated from penalized elastic net linear regression models; β (95% CI) represent a 2-standard deviation change in untransformed diversity or absolute SCFA levels (mmol/kg) per 2-standard deviation increase in ln-transformed chemical levels (refer to Tables S5 and S9 for the increments). Models were adjusted for maternal education, breastfeeding duration, C-section, and recent antibiotic use (refer to Figure S4C).

[a] Averaged coefficients from the elastic net models for those exposures which were selected (β≠0) in >50% of the 100 imputed datasets (n=298). We used 10-fold cross validation for model optimization (to determine λ with α=0.8).

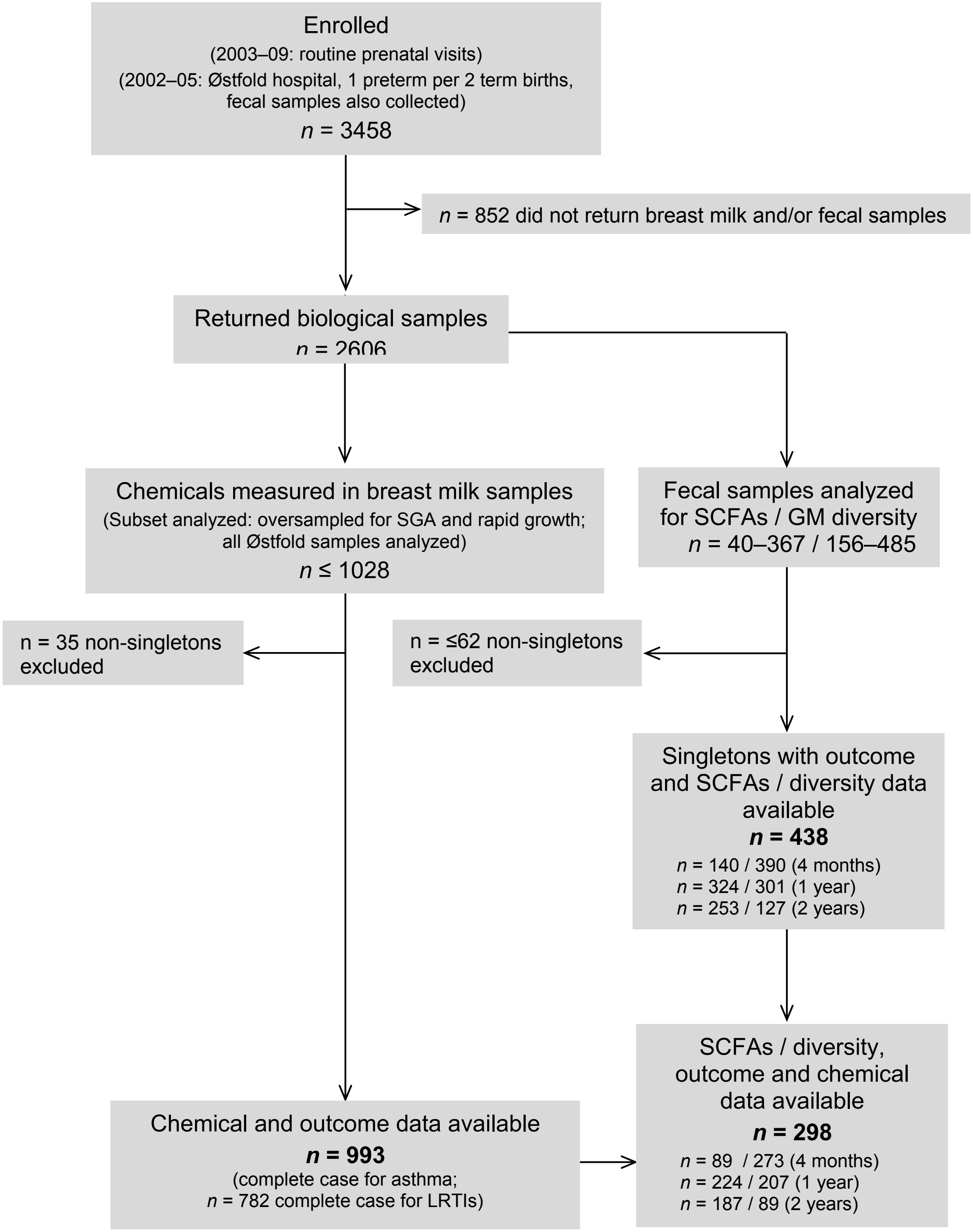

**Figure S1.** Flow chart of study population selection within the HUMIS cohort, and an overview of data used in the present analysis (bolded numbers denote the sample sizes for the primary analyses). Abbreviations: GM, gut microbiota; LRTI, lower respiratory tract infection; SCFAs, short-chain fatty acids; SGA, small-for-gestational-age.

A)

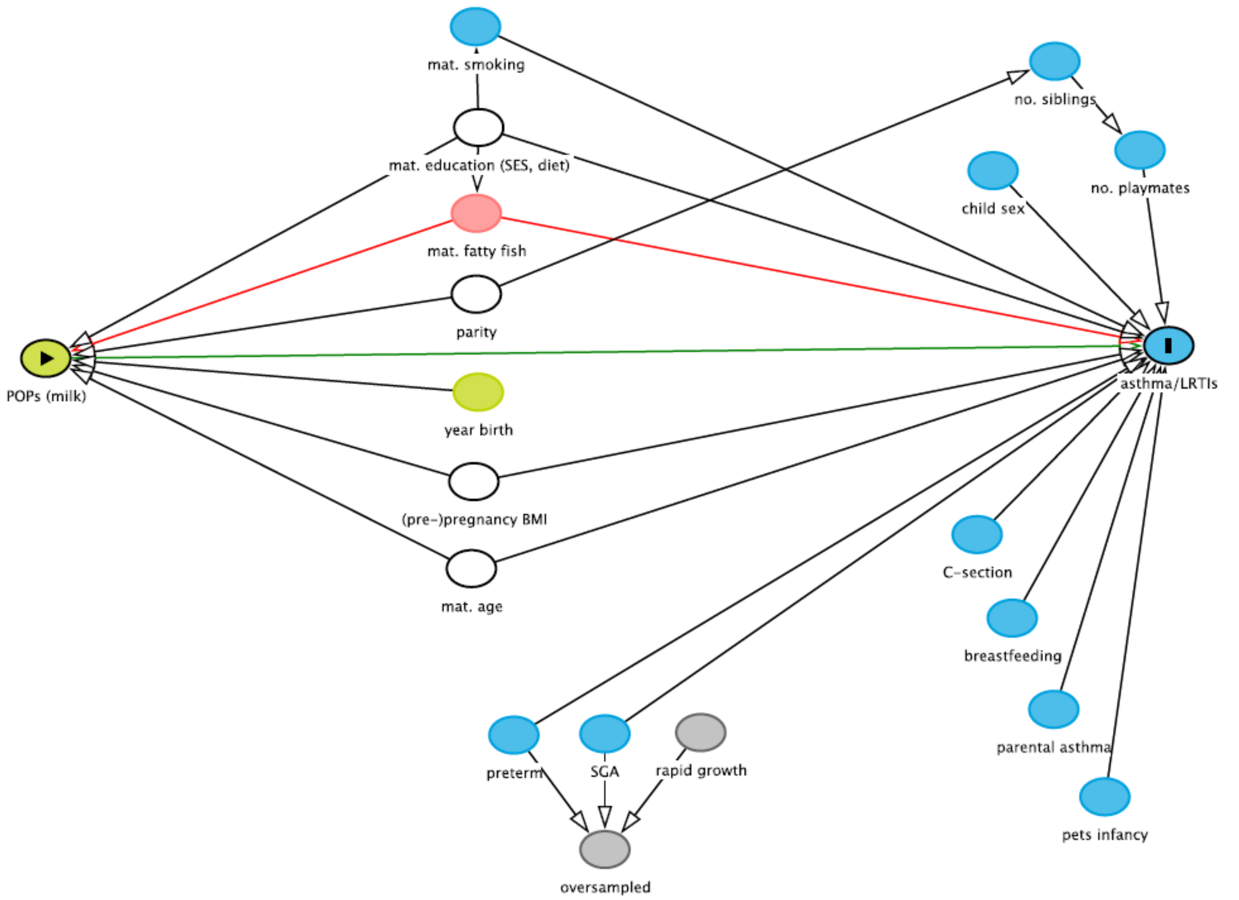

**Figure S2.** Directed acyclic graph (DAG) of the authors' conceptions of the associations between **(A)** POP exposures, covariates, and the respiratory health outcomes (asthma and LRTIs); between **(B)** SCFAs, covariates and the outcomes, and between **(C)** POPs, covariates and the outcomes. DAGs were created with DAGitty (Textor et al. 2011).
For POP–asthma/LRTI models **(A)**, we considered the minimal sufficient adjustment set to include maternal age, parity, pre-pregnancy BMI, and maternal education. We also tested models further adjusted for several covariates for which evidence for an association is weaker or which are plausibly associated with the only the outcome (leading to unnecessary adjustment, possibly decreasing precision in a logistic regression model): child birth year, child sex, delivery mode (C-section), duration of any breastfeeding, parental asthma, maternal marine fish consumption in the year before delivery, and the number of playmates at age 2, as a proxy of daycare attendance (Dogaru et al. 2014; Lignell et al. 2011; Wu et al. 2016). Covariates are defined in the main text. Abbreviations: BMI, body mass index; LRTIs, lower respiratory tract infections; SCFAs, short-chain fatty acids; SES, socioeconomic status; SGA, small for gestational age.

B)

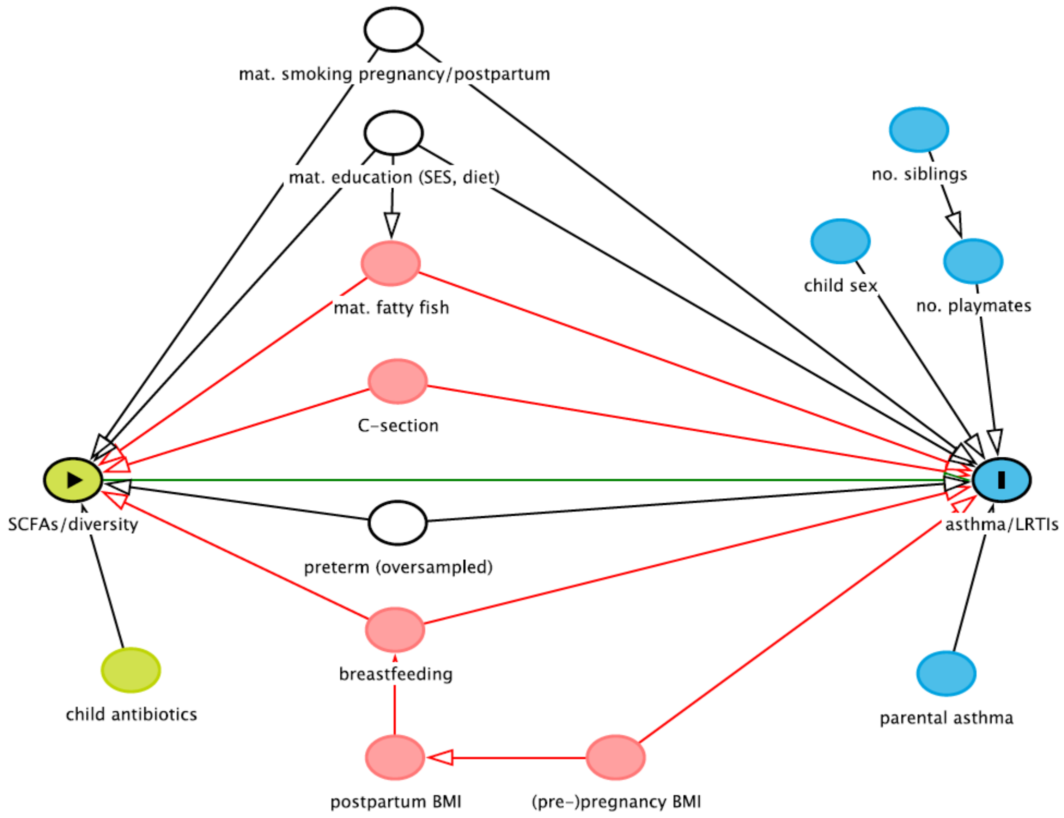

**Figure S2.** *Continued.*
For SCFA/diversity–asthma/LRTI models **(B)**, we considered the minimal sufficient adjustment set to include preterm birth (the subset enrolled in at the Østfold Hospital was oversampled for preterm birth), maternal education (as a proxy of socioeconomic status, diet, and maternal stress), and maternal smoking during pregnancy, and when the child was one year old. In further adjusted models, we also adjusted for covariates for which associations are less established, including maternal fatty fish consumption during pregnancy, breastfeeding, and delivery by C-section.

C)

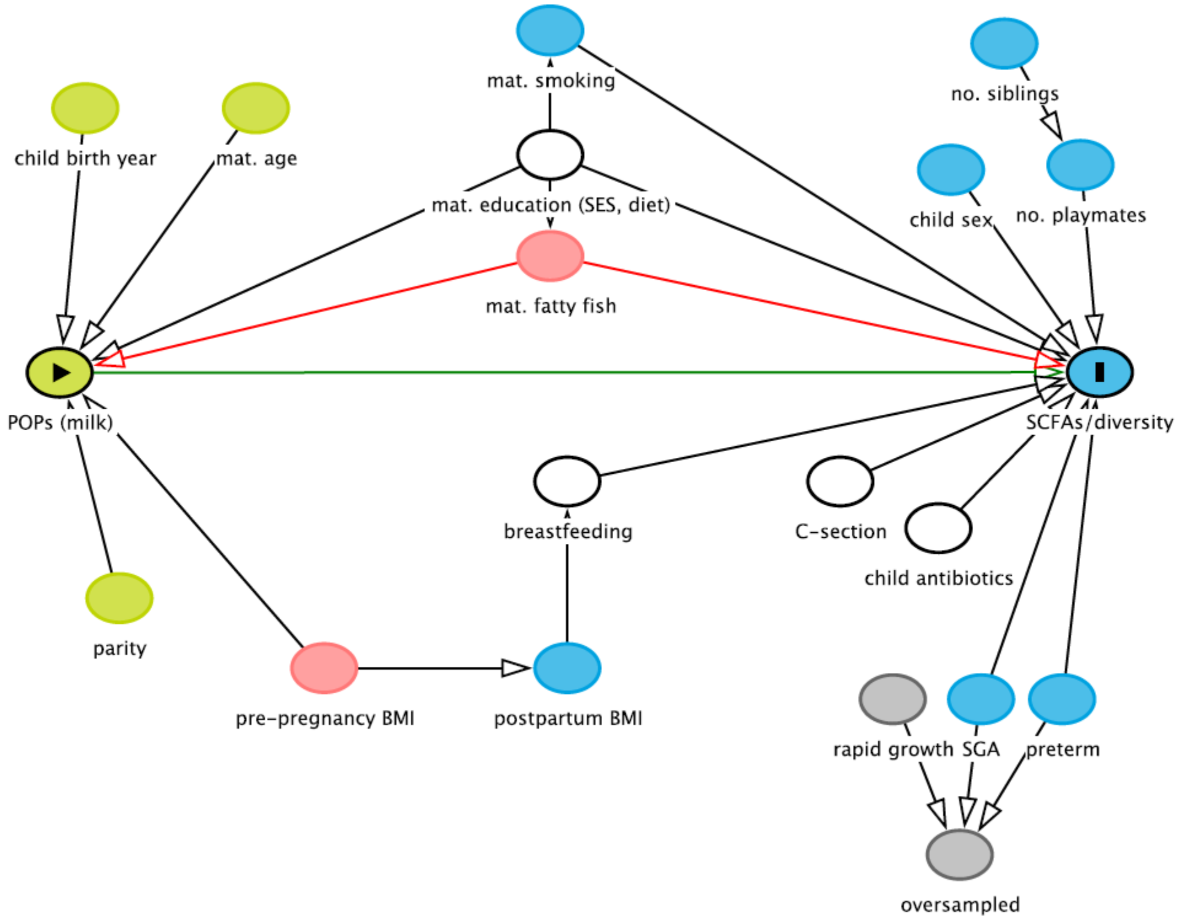

**Figure S2.** *Continued.*

For POP–asthma/LRTI models **(C)**, we considered the minimal sufficient adjustment set to include maternal education (as a proxy of SES and diet), breastfeeding, C-section, and child antibiotics. As these were linear regression models, we adjusted for some covariates likely to be strongly associated with only the outcome (‘control variables’), which can increase precision of estimates.

**Figure S3.** Mediation analysis: **(A)** a simplistic schematic; **(B)** confounding model; and **(C)** results for the two investigated exposure-mediator-outcome pathways.

A)

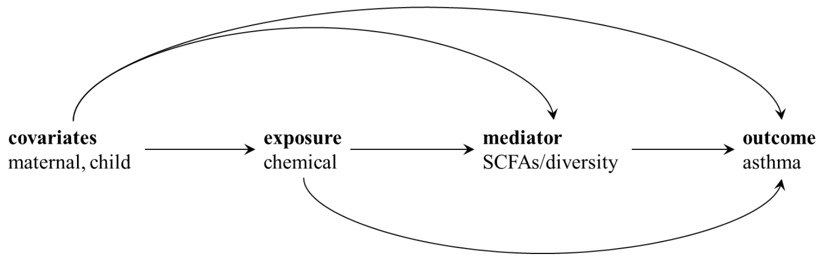

B)

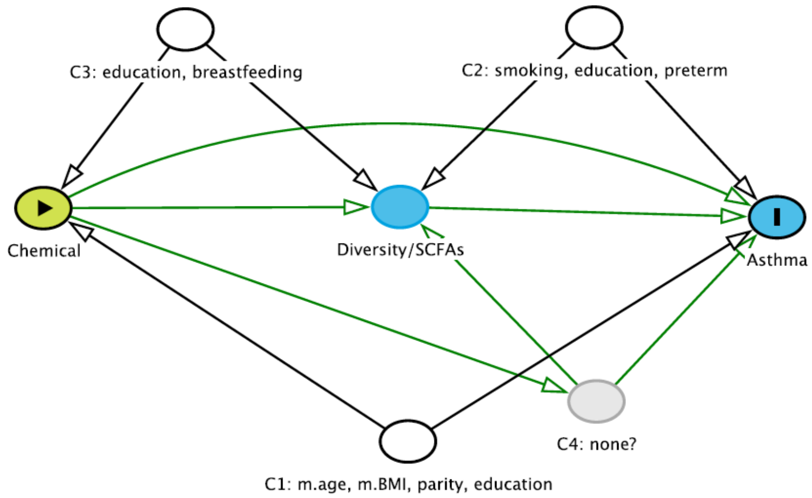

Assumptions: Sufficient control for C1) exposure-outcome, C2) mediator-outcome, C3) exposure-mediator confounding, and that C4) none of the mediator-outcome confounders are themselves affected by the exposure.

C) Mediated (i.e. indirect) natural effects assessed in the mediation analysis [with 95% confidence intervals based on bootstrapping with 1000 resamplings, where the proportion mediated = natural indirect effect/(natural direct effect + natural indirect effect)]

1. β-HCH → total SCFAs at 4 months → asthma at 10 years

| | Estimate | 95% CI | | *p*-value |
|---|---|---|---|---|
| Total Effect | -0.012 | -0.023 | 0.005 | 0.11 |
| Average natural indirect effect | 0.000 | -0.001 | 0.001 | 0.89 |
| Average natural direct effect | -0.012 | -0.023 | 0.005 | 0.11 |
| Proportion mediated | 0.000 | -0.138 | 0.143 | 0.93 |

2. ∑PCBs → Shannon diversity at 4 months → asthma at 10 years

| | Estimate | 95% CI | | *p*-value |
|---|---|---|---|---|
| Total Effect | -0.015 | -0.024 | -0.008 | 0.00 |
| Average natural indirect effect | 0.000 | -0.001 | 0.001 | 0.66 |
| Average natural direct effect | -0.015 | -0.024 | -0.008 | 0.00 |
| Proportion mediated | -0.004 | -0.083 | 0.032 | 0.66 |

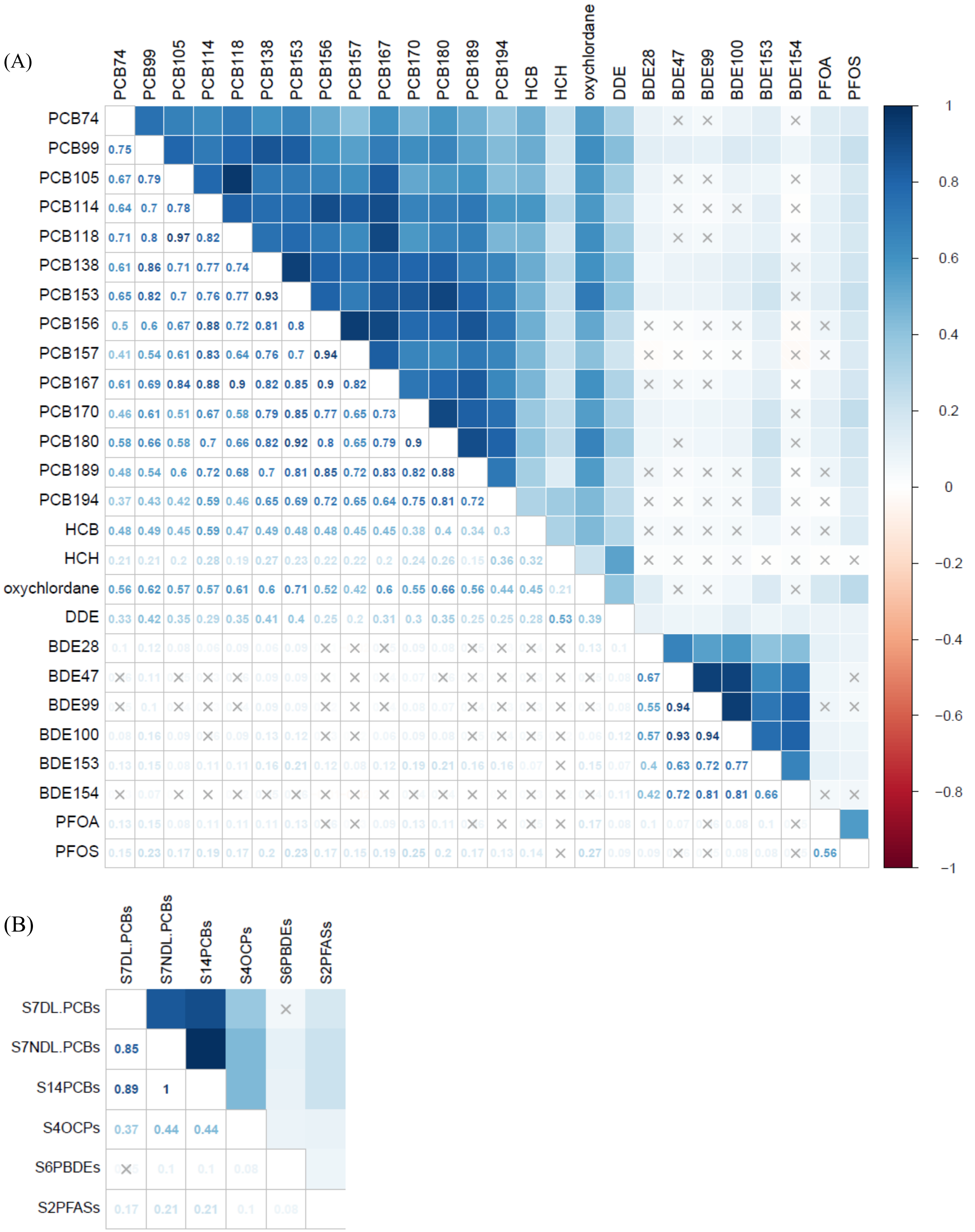

**Figure S4.** Pearson correlation coefficient matrix for the **(A)** 26 individual POP exposure biomarkers and **(B)** summed chemical groups. The intensity of the shaded squares reflects the magnitude of the correlation coefficient, crosses indicate that the correlation was non-significant ($p > 0.05$).

A)

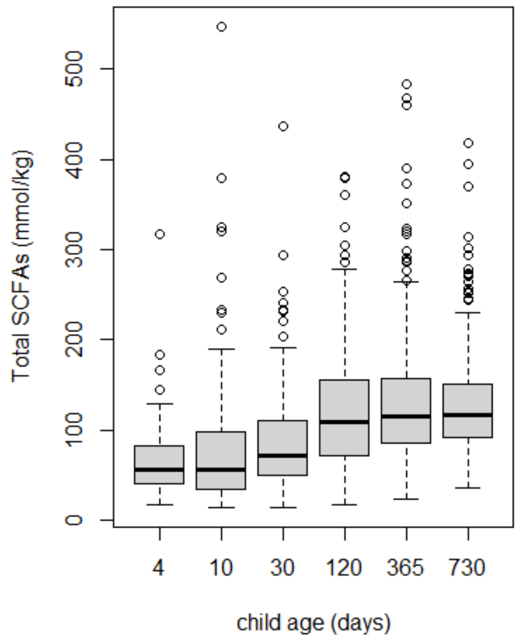

B)

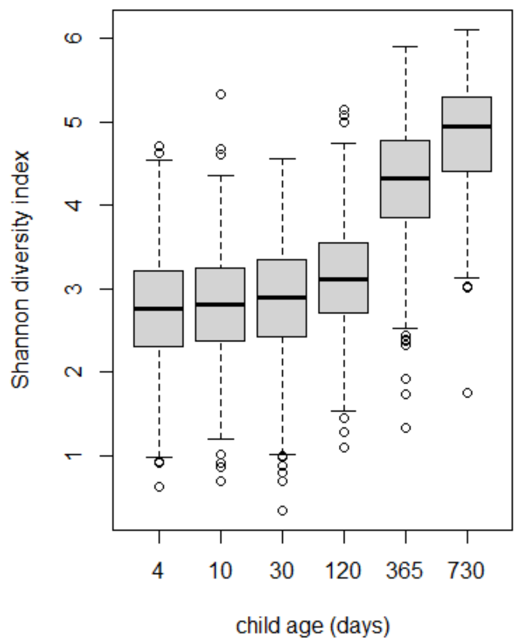

**Figure S5.** The distributions **(A–B)** and Spearman correlations **(C)** for total short-chain fatty acid (SCFA) levels and Shannon diversity at the sample collection time points (when the child was 4, 10, and 30 days, and 4, 12, and 24 months old). In plot C, crosses indicate that the Spearman correlation coefficient was non-significant ($p > 0.05$).

C)

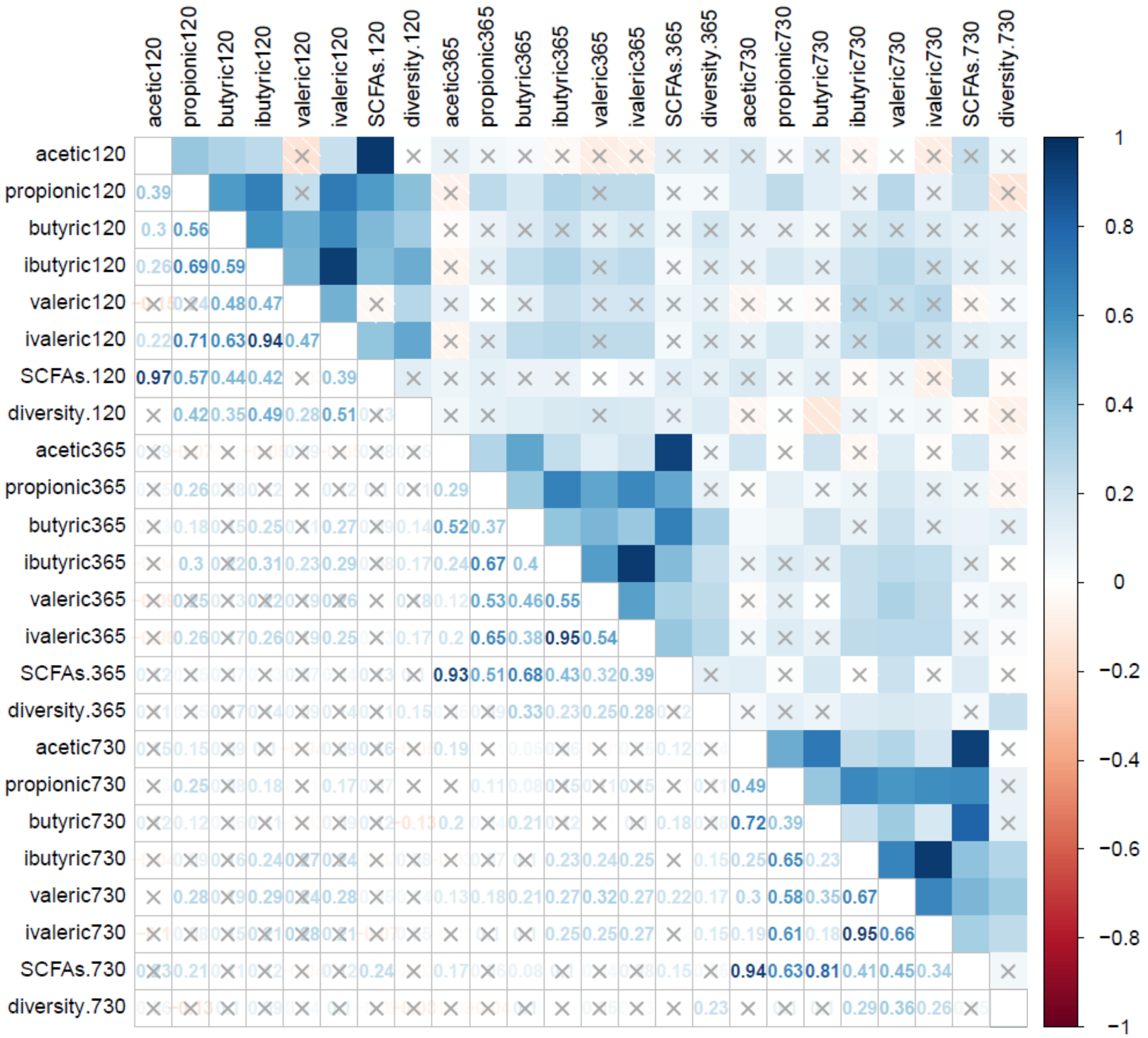

**Figure S5.** *Continued.*
